# dynaPhenoM: Dynamic Phenotype Modeling from Longitudinal Patient Records Using Machine Learning

**DOI:** 10.1101/2021.11.01.21265725

**Authors:** Hao Zhang, Chengxi Zang, Jie Xu, Hansi Zhang, Sajjad Fouladvand, Shreyas Havaldar, Chang Su, Feixiong Cheng, Benjamin S. Glicksberg, Jin Chen, Jiang Bian, Fei Wang

**Affiliations:** Division of Health Informatics, Department of Population Health Sciences, Weill Cornell Medicine, New York, NY, USA; Department of Health Outcomes & Biomedical Informatics, University of Florida, Gainesville, Florida, USA; Institude for Biomedical Informatics (IBI) and Department of Computer Science, University of Kentucky, Lexington, KY, USA; Hasso Plattner Institute for Digital Health at Mount Sinai, Icahn School of Medicine at Mount Sinai, New York, New York, USA; Department of Health Service Administration and Policy (HSAP), College of Public Health, Temple University, PA, USA; Genomic Medicine Institute, Lerner Research Institute, Cleveland Clinic, Cleveland, OH, USA; Department of Molecular Medicine, Cleveland Clinic Lerner College of Medicine, Case Western Reserve University, Cleveland, OH, USA; Case Comprehensive Cancer Center, Case Western Reserve University School of Medicine, Cleveland, OH, USA

## Abstract

Identification of clinically meaningful subphenotypes of disease progression can facilitate better understanding of disease heterogeneity and underlying pathophysiology. We propose a machine learning algorithm, termed dynaPhenoM, to achieve this goal based on longitudinal patient records such as electronic health records (EHR) or insurance claims. Specifically, dynaPhenoM first learns a set of coherent clinical topics from the events across different patient visits within the records along with the topic transition probability matrix, and then employs the time-aware latent class analysis (T-LCA) procedure to characterize each subphenotype as the evolution of these learned topics over time. The patients in the same subphenotype have similar such topic evolution patterns. We demonstrate the effectiveness and robustness of dynaPhenoM on the case of mild cognitive impairment (MCI) to Alzheimer’s disease (AD) progression on three patient cohorts, and five informative subphenotypes were identified which suggest the different clinical trajectories for disease progression from MCI to AD.

## Introduction

Due to the complex and heterogeneous nature of human diseases such as Alzheimer’s disease (AD), patients usually demonstrate diverse clinical manifestations. Identification of clinically meaningful subphenotypes, which are subgroups of patients with coherent clinical characteristics, is critical for improved understanding of the underlying disease mechanisms and inform precision medicine (*1*) (*2*). In recent years, with the increasing adoption of various health information systems such as electronic health records (EHR), comprehensive information about patients have been accumulated, such as demographics, diagnosis, medications, lab tests, etc. (*3*). With these diverse data, there have been existing studies developing data-driven approaches to identify disease subphenotypes (*4-7*), but they typically focused on a set of selected clinical events but did not consider temporal evolutions of these events.

To effectively explore the clinical information within the patient records and identify comprehensive disease subphenotypes, we need to address the following general challenges for analyzing these data: *i*) ***Information Heterogeneity***: These data contain different types of information as mentioned above; *ii*) ***Irregular Visits***: the time intervals between any two successive patient visits are typically irregular; *iii*) ***Missing Values***: there is substantial missing information in patient records (e.g., there will not be any record if a patient did not pay a visit to the clinic, but it does not mean the patient is without the disease); *iv*) ***High-Dimensionality and Sparsity***. Clinical events within patient records are represented as systematic codes with large vocabularies (e.g., there are ∼68,000 distinct diagnosis codes for in ICD-10, which stands for International Classification of Diseases, 10^th^ version (*8*)), and every patient visit only has a few codes (*9*); v) ***Interpretability***. It is critical to make the analysis results interpretable and easy to understand by the clinicians.

With all these considerations, in this paper, we propose a machine learning framework named dynaPhenoM to derive disease progression subphenotypes from longitudinal patient records. Progression subphenotype indicates that patients belonging to the same subphenotype have similar temporal evolution patterns of the clinical events in their records. The overall architecture of dynaPhenoM is shown in Figure 1. After data preprocessing, dynaPhenoM contains two main modules: the dynamic multimodal topic model (DMTM) for deriving new interpretable compressed representations of multimodal clinical events, and the time-aware latent class analysis (T-LCA) for subphenotyping that embeds the time of irregular visits.

**Figure 1.**
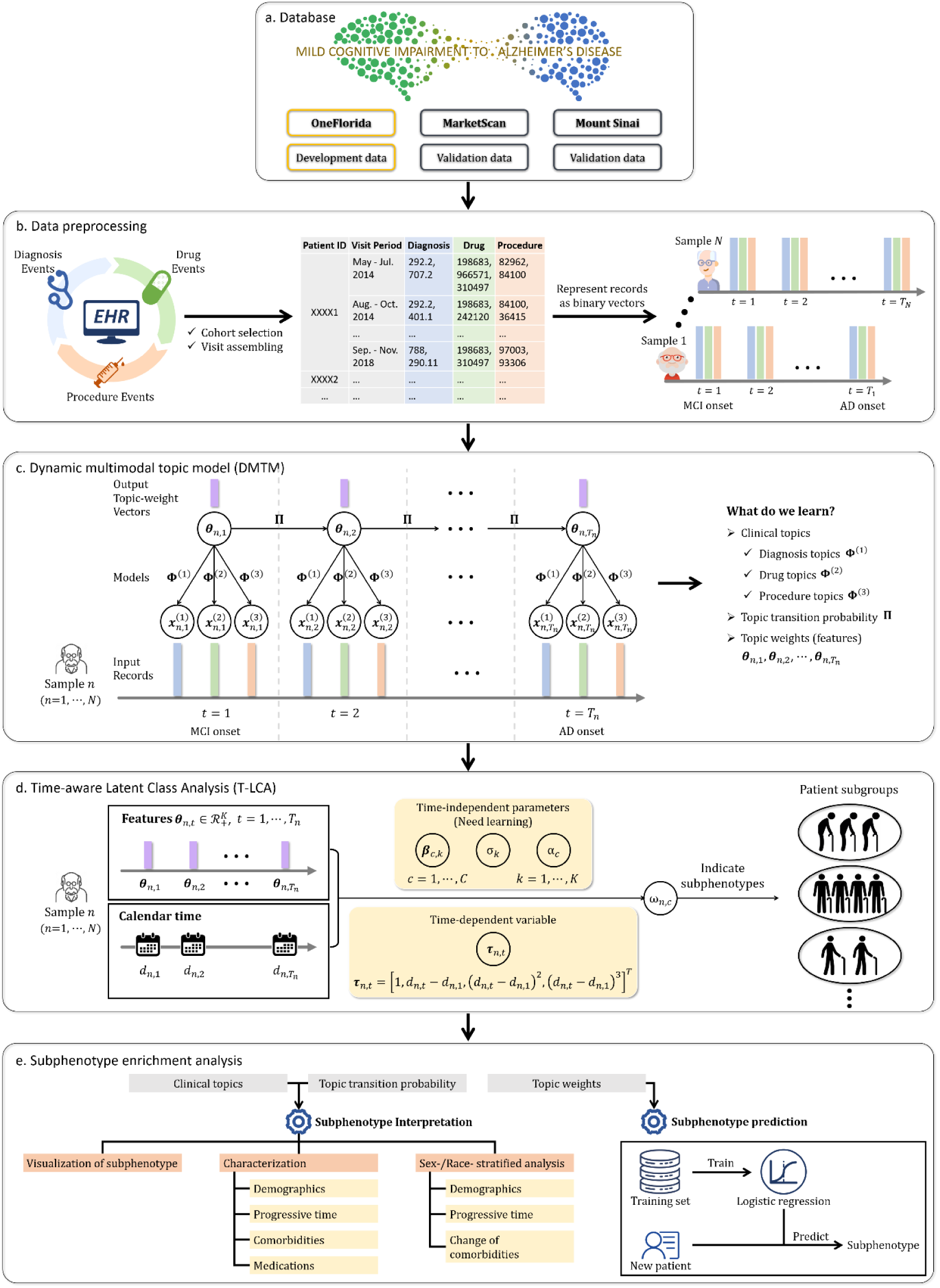
Workflow of dynaPhenoM for deriving longitudinal subphenotypes from longitudinal patient records demonstrated on the case of mild cognitive impairment (MCI) to Alzheimer’s disease (AD) progression. a. Dataset for demonstration of MCI to AD progression. b. Data preprocessing from the original longitudinal patient records for MCI to AD progression, including cohort selection, visit assembling, and representing records as binary vectors. Currently, we aggregate all records within every three months as a single “visit”, while this window size can be tuned according to different cases. For every visit, records from one modality can be represented as a binary vector (1: the visit includes this code, 0: the visit does not include this code) where the length of this binary vector is equal to the total number of unique codes in the modality (different modalities can have different number of unique codes). c. Illustration of DMTM. DMTM regards binary longitudinal vectors as input and output the clinical topics including different modalities, topic transition probability, and topic weights. Clinical topics and topic transition probability are shared by all patients at every visit (global parameters) while topic weights are new features to characterize the patients (patient-specific, local parameters). d. Illustration of T-LCA. T-LCA regards topics weights as input to identify longitudinal subphenotypes, which embeds calendar time of each visit into subphenotyping. e. Utilizing interpretable clinical topics and topic transition probability learned from DMTM, dynaPhenoM performs the subphenotype interpretation, gender and race stratified enrichment analysis, and builds the logistic regression to predict the subphenotype belongings for new patients using early-stage records.

DMTM builds on the concepts of latent topic modeling (LTM) (*10*) which is often used in text-mining tasks. In analogy with test mining, DMTM considers clinical events denoted by codes as words and each visit as a document. DMTM is also related to some studies (*9, 11, 12*) that use methods of matrix factorization or LTM to learn compressed representations from original EHR data. However, existing methods focus on the single visit of each patient while DMTM learns representations from longitudinal information. LCA (*13*) is a widely used subphenotyping method in clinical studies (*14-16*). When LCA is applied for deriving longitudinal subphenotypes (*17*), it does not consider a fact that time intervals between any two successive patient visits are typically irregular. Motivated by this problem, we developed T-LCA in dynaPhenoM.

To demonstrate the effectiveness of dynaPhenoM, we apply it to identify the progression subphenotypes for patients who progressed from mild cognitive impairment (MCI) to Alzheimer’s disease (AD). AD is the most prevalent neurodegenerative disorder that affects millions of people all over the world (*18*) and its prevalence is expected to double in the next 20 years (*19*). The underlying disease mechanism of AD is highly complex and there is no effective treatment for AD yet (*20*). On the other hand, MCI is the stage between the expected cognitive decline of normal aging and dementia, including AD. Understanding the clinical heterogeneity of patients progressing from MCI to AD and identifying the corresponding progression subphenotypes can potentially reveal the different underlying disease pathophysiology and shed light on effective treatments. We leveraged three real world patient record databases, including one national insurance claims database, one EHR database from a state clinical research network, and one EHR database from a regional health system, to achieve our goal. Five clinically-meaningful subphenotypes were identified from the development cohort and validated on the other two cohorts. We performed extensive statistical analysis to interpret these subphenotypes, and we have also built predictive models to investigate whether these subphenotype can be identified early.

## Results

### dynaPhenoM as a framework to identify longitudinal subphenotypes

The overall workflow of dynaPhenoM is illustrated in Figure 1, including two key components: DMTM and T-LCA. DMTM learns new representations of patient visits through a dynamic multi- modal topic modeling process. Considering the irregular patient visits, T-LCA derives the progression subphenotypes from the trajectories of these new representations through an LCA (*13*) type of process.

Specifically, after representing different types of clinical events as respective binary vectors, we first use DMTM to learn a set of multimodal clinical topics from the patient records, where each topic can be viewed as a set of clinical events that are more likely to co-occur within a patient visit. Then for every patient visit, DMTM infers the mixture memberships, also called topic weights, as the new representations of this visit. Higher weight on a particular topic indicates that the corresponding patient visit includes more events from this topic. This topic weight based representation transforms the visit representation from the original high-dimensional binary space to a low-dimensional continuous space, and each dimension in this space is a topic composed of a set of frequently co-occurred clinical events. Thus this representation is highly interpretable. By concatenating the learned representations for all visits of a specific patient according to the timeline, we can get a multi-variate temporal sequence for the patient with irregular intervals. We then derive disease progression subphenotypes by grouping these temporal sequences with T-LCA. Technical details of these two modules are provided in Method and Supplement.

### Cohort Definition

We derived the progression subphenotypes from MCI to AD from the development cohort, and got them validated in two validation cohorts.

#### Development cohort

We leveraged the patient EHR from OneFlorida Clinical Research Consortium (*21*) —a clinical data research network funded by the Patient-Centered Outcomes Research Institute (PCORI) contributing to the national Patient-Centered Clinical Research Network (PCORnet)—to derive the subphenotypes. We used diagnosis codes (detailed in Supplemental Table 1) to identify a total of 5337 patients who experienced the progression from MCI to AD, and among them 2,995 patients whose progression time were longer than one year were included in our development cohort.

#### Validation cohorts

We validated the derived subphenotypes on two independent cohorts. One is the large-scale administrative records in the IBM Health MarketScan Commercial Claims database (*22*) for the years 2009 to 2020. The second one is the patient EHR data from the Mount Sinai Health System which contains five locations in New York City. Similar to the development cohort, we finally obtained 18,805 patients from MarketScan, and 698 patients from Mount Sinai for validating subphenotypes.

Details of these three studied cohorts are summarized in Table 1, where the corresponding codes about key comorbidities and medications are listed in Supplemental Table 2∼4. Detailed descriptions of cohorts are provided in Methods.

**Table 1.**
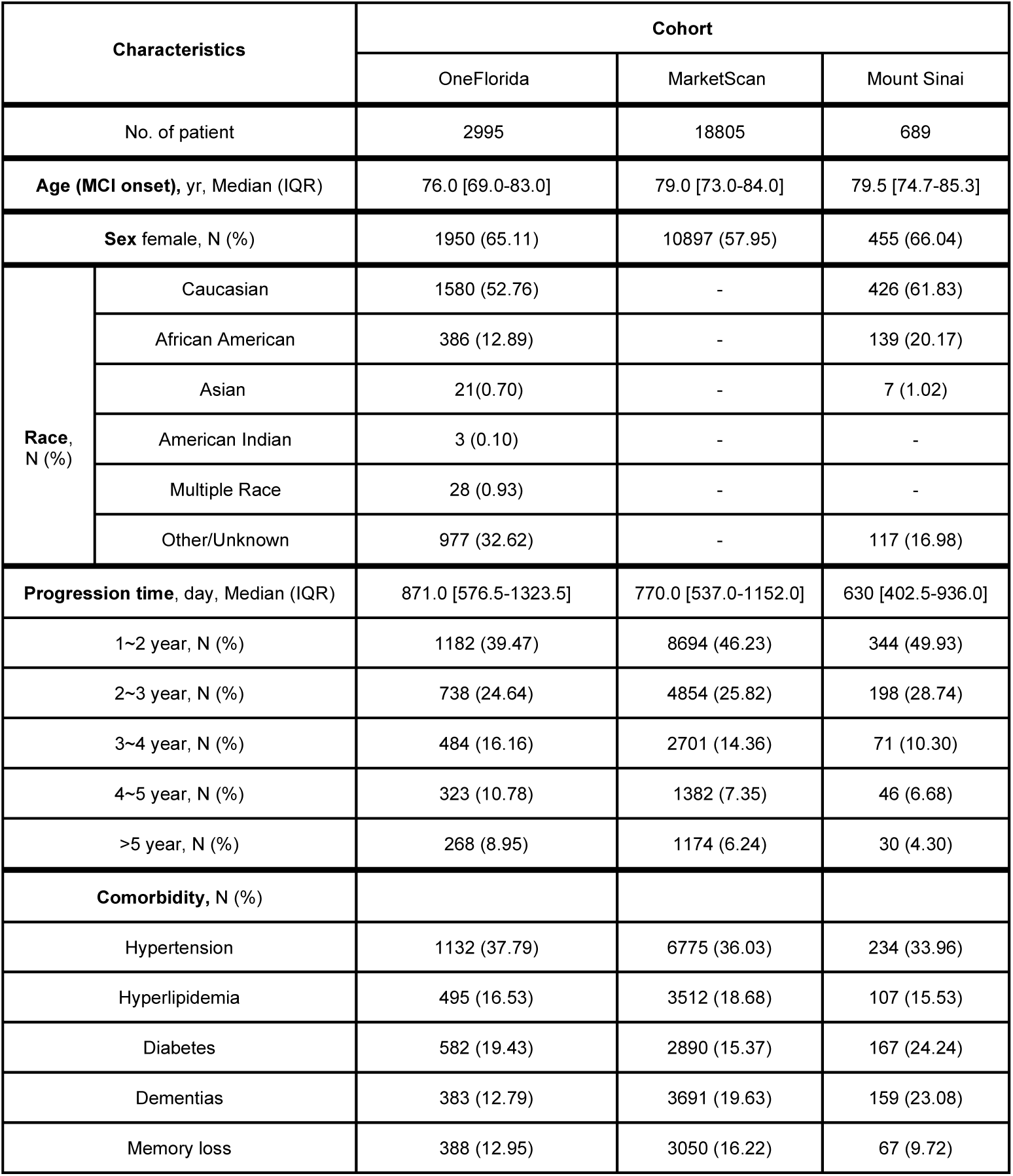
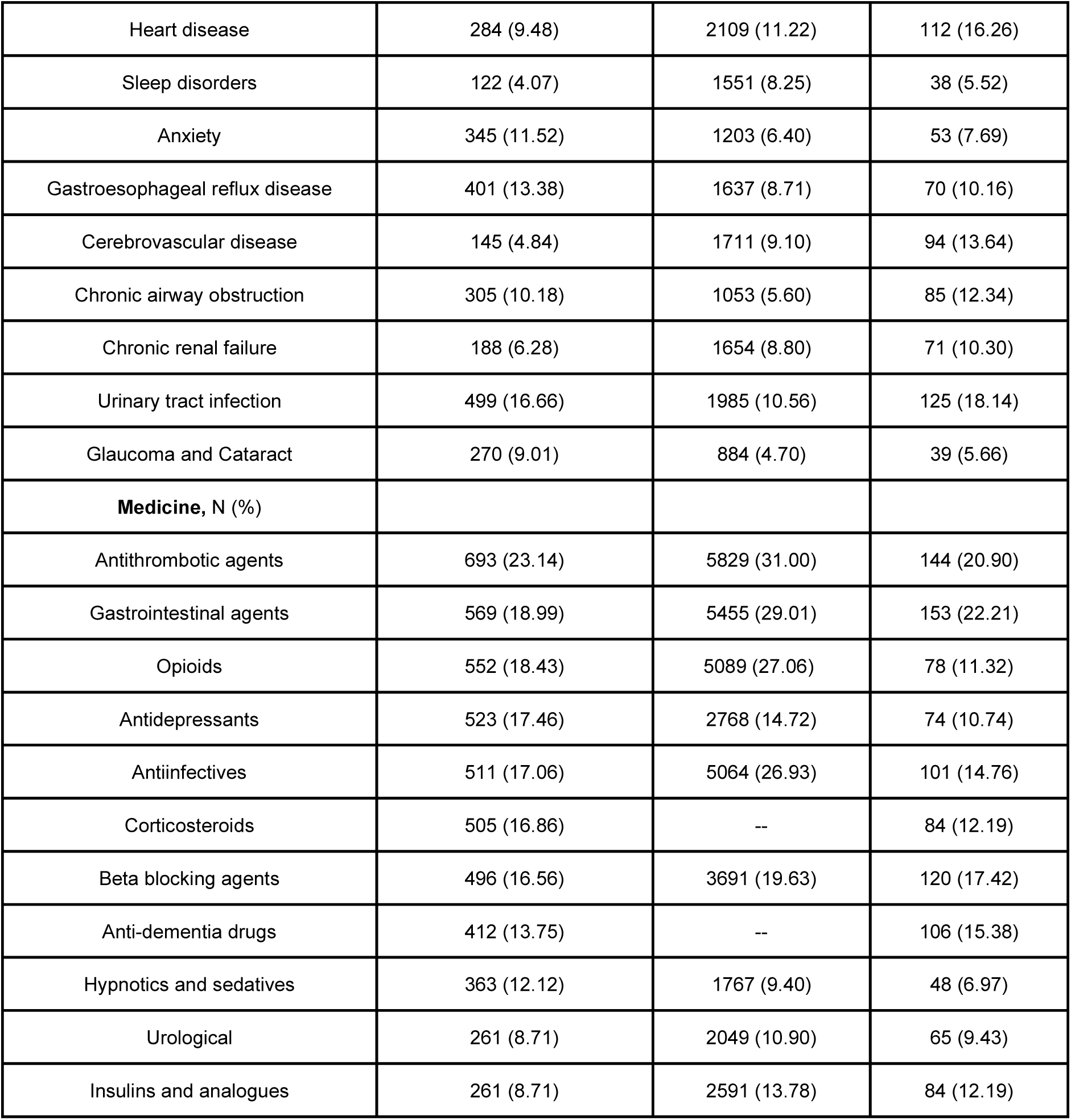
Characteristics of the development (OneFlorida) and the two external validation (MarketScan and Mount Sinai) cohorts.

| Characteristics |  | Cohort |  |  |
| --- | --- | --- | --- | --- |
|  |  | OneFlorida | MarketScan | Mount Sinai |
| No. of patient |  | 2995 | 18805 | 689 |
| Age (MCI onset), yr, Median (IQR) |  | 76.0 [69.0-83.0] | 79.0 [73.0-84.0] | 79.5 [74.7-85.3] |
| Sex female, N (%) |  | 1950 (65.11) | 10897 (57.95) | 455 (66.04) |
| Race, N (%) | Caucasian | 1580 (52.76) | - | 426 (61.83) |
|  | African American | 386 (12.89) | - | 139 (20.17) |
|  | Asian | 21(0.70) | - | 7 (1.02) |
|  | American Indian | 3 (0.10) | - | - |
|  | Multiple Race | 28 (0.93) | - | - |
|  | Other/Unknown | 977 (32.62) | - | 117 (16.98) |
| Progression time, day, Median (IQR) |  | 871.0 [576.5-1323.5] | 770.0 [537.0-1152.0] | 630 [402.5-936.0] |
| 1~2 year, N (%) |  | 1182 (39.47) | 8694 (46.23) | 344 (49.93) |
| 2~3 year, N (%) |  | 738 (24.64) | 4854 (25.82) | 198 (28.74) |
| 3~4 year, N (%) |  | 484 (16.16) | 2701 (14.36) | 71 (10.30) |
| 4~5 year, N (%) |  | 323 (10.78) | 1382 (7.35) | 46 (6.68) |
| >5 year, N (%) |  | 268 (8.95) | 1174 (6.24) | 30 (4.30) |
| Comorbidity, N (%) |  |  |  |  |
| Hypertension |  | 1132 (37.79) | 6775 (36.03) | 234 (33.96) |
| Hyperlipidemia |  | 495 (16.53) | 3512 (18.68) | 107 (15.53) |
| Diabetes |  | 582 (19.43) | 2890 (15.37) | 167 (24.24) |
| Dementias |  | 383 (12.79) | 3691 (19.63) | 159 (23.08) |
| Memory loss |  | 388 (12.95) | 3050 (16.22) | 67 (9.72) |

|  |  |  |  |
| --- | --- | --- | --- |
| Heart disease | 284 (9.48) | 2109 (11.22) | 112 (16.26) |
| Sleep disorders | 122 (4.07) | 1551 (8.25) | 38 (5.52) |
| Anxiety | 345 (11.52) | 1203 (6.40) | 53 (7.69) |
| Gastroesophageal reflux disease | 401 (13.38) | 1637 (8.71) | 70 (10.16) |
| Cerebrovascular disease | 145 (4.84) | 1711 (9.10) | 94 (13.64) |
| Chronic airway obstruction | 305 (10.18) | 1053 (5.60) | 85 (12.34) |
| Chronic renal failure | 188 (6.28) | 1654 (8.80) | 71 (10.30) |
| Urinary tract infection | 499 (16.66) | 1985 (10.56) | 125 (18.14) |
| Glaucoma and Cataract | 270 (9.01) | 884 (4.70) | 39 (5.66) |
| **Medicine, N (%)** |  |  |  |
| Antithrombotic agents | 693 (23.14) | 5829 (31.00) | 144 (20.90) |
| Gastrointestinal agents | 569 (18.99) | 5455 (29.01) | 153 (22.21) |
| Opioids | 552 (18.43) | 5089 (27.06) | 78 (11.32) |
| Antidepressants | 523 (17.46) | 2768 (14.72) | 74 (10.74) |
| Antiinfectives | 511 (17.06) | 5064 (26.93) | 101 (14.76) |
| Corticosteroids | 505 (16.86) | -- | 84 (12.19) |
| Beta blocking agents | 496 (16.56) | 3691 (19.63) | 120 (17.42) |
| Anti-dementia drugs | 412 (13.75) | -- | 106 (15.38) |
| Hypnotics and sedatives | 363 (12.12) | 1767 (9.40) | 48 (6.97) |
| Urological | 261 (8.71) | 2049 (10.90) | 65 (9.43) |
| Insulins and analogues | 261 (8.71) | 2591 (13.78) | 84 (12.19) |

The analysis on OneFlorida data is approved by University of Florida institutional review board under number IRB202000704. The analysis on MarketScan data is approved by University of Kentucky CCTS Enterprise Data Center institutional review board under number 43542. The analysis on Mount Sinai data is approved by institutional review board under number IRB-19- 02369.

### Learning clinical topics with multimodal information

The DMTM module in dynaPhenoM learns multimodal clinical topics from the collection of the records in all patient visits longitudinal records. To choose the optimal number of topics (*k*), on development cohort, we performed five-fold cross-validation to evaluate the data likelihood and topic coherence with different *k* (Figure 4a in Supplement), and finally we set *k* = 30. According to the percentage of mean topic weights (Figure 5 in Supplement; defined in Methods), we selected the 13 prevalent clinical topics (others were shown in Figure 6 in Supplement) learned from the development cohort, which are demonstrated in Figure 2.

**Figure 2.**
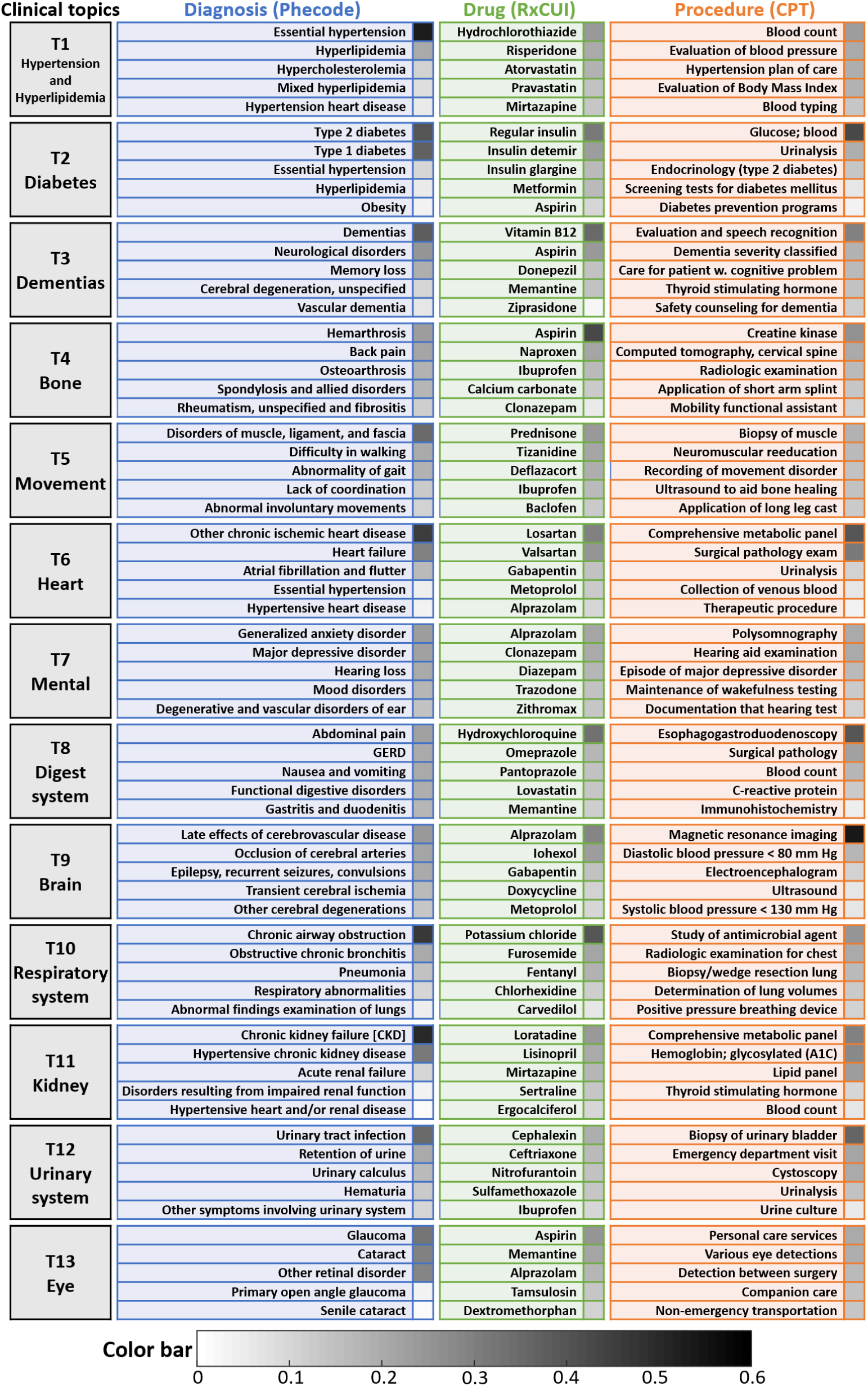
Key (commonly used) clinical topics learned from the development cohort by DMTM, where the color bar indicates the weights of each events in the corresponding topics.

In Figure 2, each clinical topic is represented with three modalities: disease, medication, and procedure, which are then described with the top-5 most related clinical events according to their weights in each topic shown in the color bar. From the figure we can observe that these topics are typically associated with particular disease conditions. For example, topic T11 is related to kidney diseases such as chronic kidney disease (CKD), which may lead to the accumulation of uremic toxins which acts as a high risk factor of cognition impairment and AD (*23*). Topic T1 is related to cardiovascular conditions which have been known to be risk factors of AD (*24-26*). Although the exact mechanism on how cognitive decline and diabetes (in T2) are connected is not clear yet, researchers have shown that that high blood sugar or insulin can harm the brain in several ways (*27, 28*) like high blood sugar causing inflammation that may damage brain cells and cause cognitive impairment. Similarly, there has been research showing that certain mental disorders (in T7) such as anxiety, depression, and hearing loss are commonly observed neuropsychiatric comorbidities of MCI or AD (*29, 30*).

We observe strong coherence across the three modalities for each topic. Such coherence can be reflected on many different aspects including, but not limited to i) medications treating diseases, such as Donepezil/Memantine and dementia (in T3); ii) medications treating disease comorbidities, for example, in T11 (Kidney), major depressive disorder affects one in five patients with CKD, and Sertraline is a potential antidepressant treating for CKD patients with depression (*31*); iii) medications causing disease conditions as side-effects, for example, in T6 (Heart), Gabapentin is a widely used analgesic, anticonvulsant and anxiolytic agent, but authors in (*32*) reported that taking Gabapentin will increase the risk of having heart failure for elderly patients; iv) procedures associated with diseases, such as evaluating blood pressure and body mass index for patients with cardiovascular diseases (T1). Moreover, with the multimodal topics, given one clinical event, we constructed its interactions with other events by calculating their similarities (see Methods), and the detailed results are provided in Supplemental Figure 7.

### Transition probabilities across different clinical topics

Discovering the transition patterns across clinical topics is helpful for understanding the clinical progression of diseases (MCI to AD in our case). Figure 3 shows the transition probabilities across all clinical topics (we summarized the remaining less prevalent 17 topics as others) on the development cohort, where the value of *(i, j)*-th entry represents the transition probability (%) from *i*-th topic to *j*-th topic in two consecutive visits.

**Figure 3.**
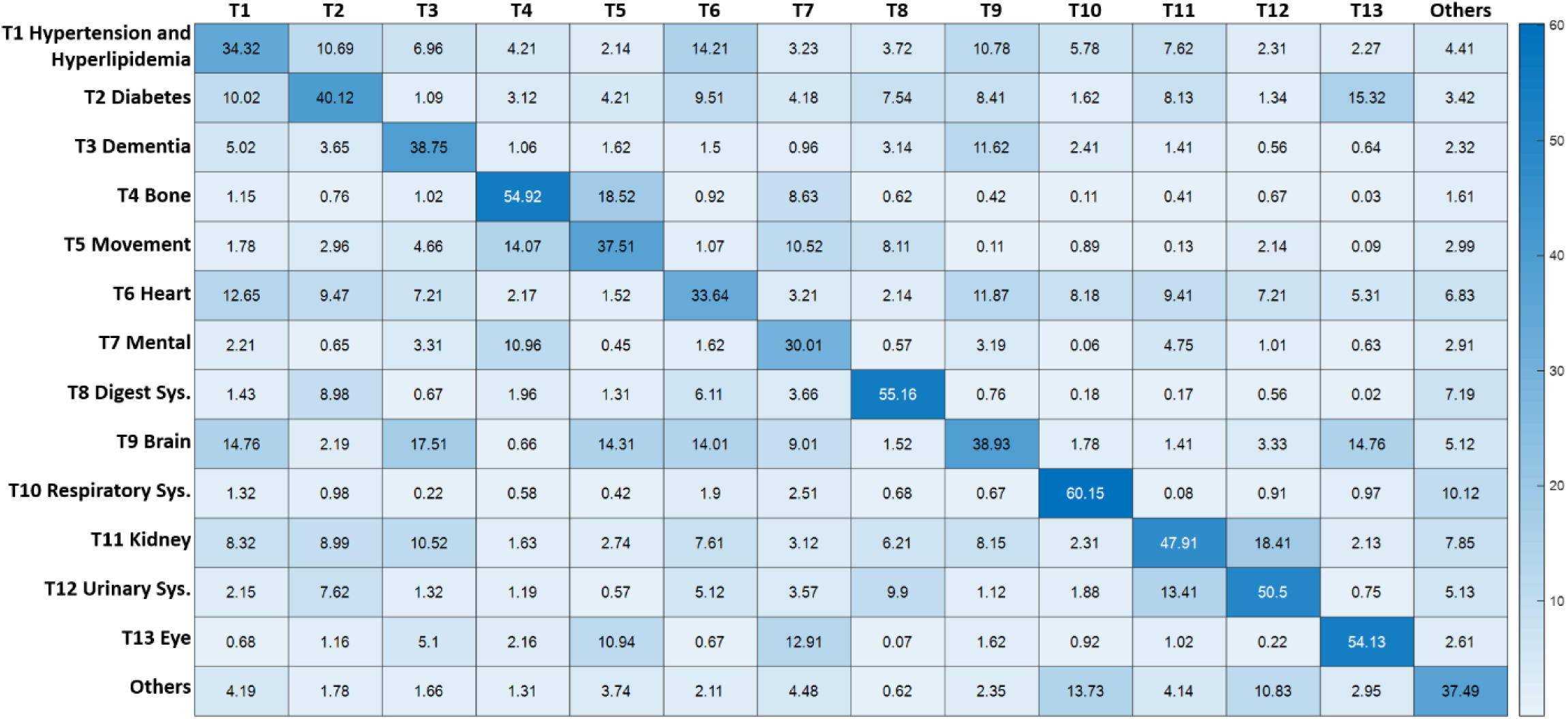
Matrix of topic transition probabilities (%) on the development cohort. Besides the 13 key clinical topics, other 17 topics are integrated into “others” in the matrix. The value in *i*-th row and *j*-th column denotes the transition probability from *ii*-th topic to *j*-th topic.

The figure demonstrates that the diagonal values of the transition matrix are bigger, which suggests that the disease topics for consecutive patient visits tend to stay the same.

In addition, we have also observed other entries with relatively larger values such as transition from cardiovascular disease, including hypertension and hyperlipidemia, to heart disease (T1->T6: 14.21%), brain disease (T1->T9: 10.78%), and diabetes (T1->T2: 10.69%) (*33-37*). From brain disease to eye disease (T9-> T13, 14.76%) and mental problems (T9->T7, 9.01%), as well as from eye disease to mental problems (T13->T7, 12.91%). All these transitions have been demonstrated in prior studies (*38-40*). Other high probability entries include the transitions between T4 (bone) and T5 (movement), T11 (kidney) and T12 (urinary system). All these transitions can also be observed on the derived subphenotypes which are detailed in the next subsection.

These results on clinical topics and their transition probabilities shows that DMTM is able to learn interpretable and clinically meaningful topics. Based on them, DMTM infers topic weights as a new representation for each patient visit in a low-dimensional continuous space, which facilitates the subsequent derivations of progression subphenotypes.

### Progression subphenotypes

With the new representations learned from DMTM, on the development cohort, we used T-LCA to identify five subphenotypes including 2254 (75.26%) patients (see details in Supplemental Method). Figure 4 visualizes these subphenotypes, where the horizontal axis is the calendar time (in month) starting from MCI onset, and vertical axis represents the average (over patients within the corresponding subphenotype) number of diagnosis codes in one topic whose probabilities of occurrence are larger than 0.5. Therefore, larger values on the vertical axis indicates more diagnosis events from the corresponding topic tend to appear (detailed in Method). We demonstrate these subphenotypes according to the change of their topic compositions in Figure 4a, where major topics whose value exceeds 2 on vertical axis at least once during the entire progression course are highlighted in solid lines. Figure 4b illustrates the evolution of each topic within different subphenotypes. Characteristics including demographics, progression time, key comorbidities and medications of these subphenotypes at MCI onset are shown in Table 2 (detailed codes are provided in supplemental Table 2-4). The Kaplan-Meier survival curves with AD onset as outcome event (starting from MCI onset) were shown in Figure 5, which provides a comprehensive picture on the progression speed across the 5 identified subphenotypes. For each subphenotype, we also showed the change in percentage of patients with different comorbidities during the progression (Supplemental Figure 8), and the percentage of patients taking certain medications during the progression (Supplemental Figure 9).

**Table 2.**
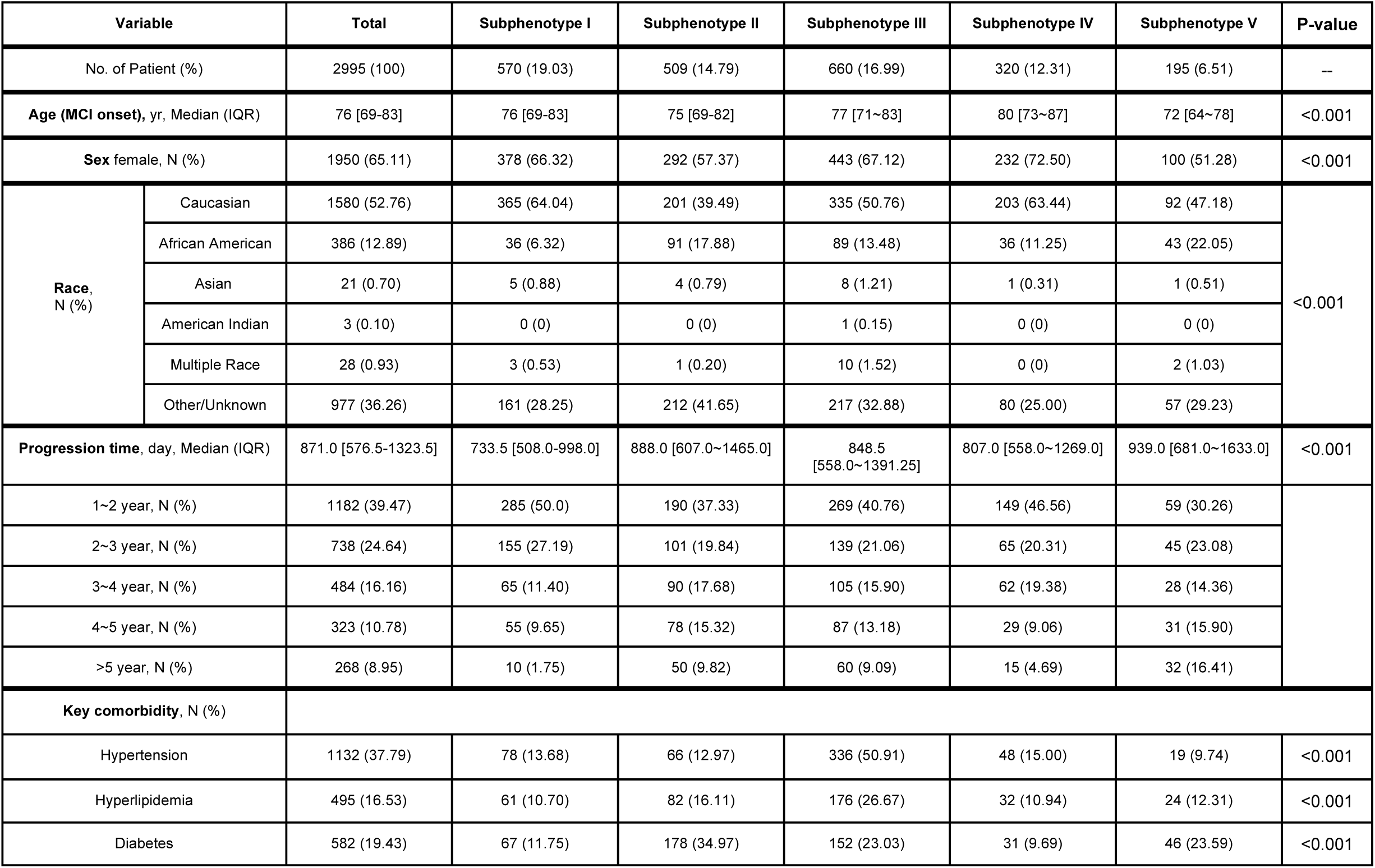
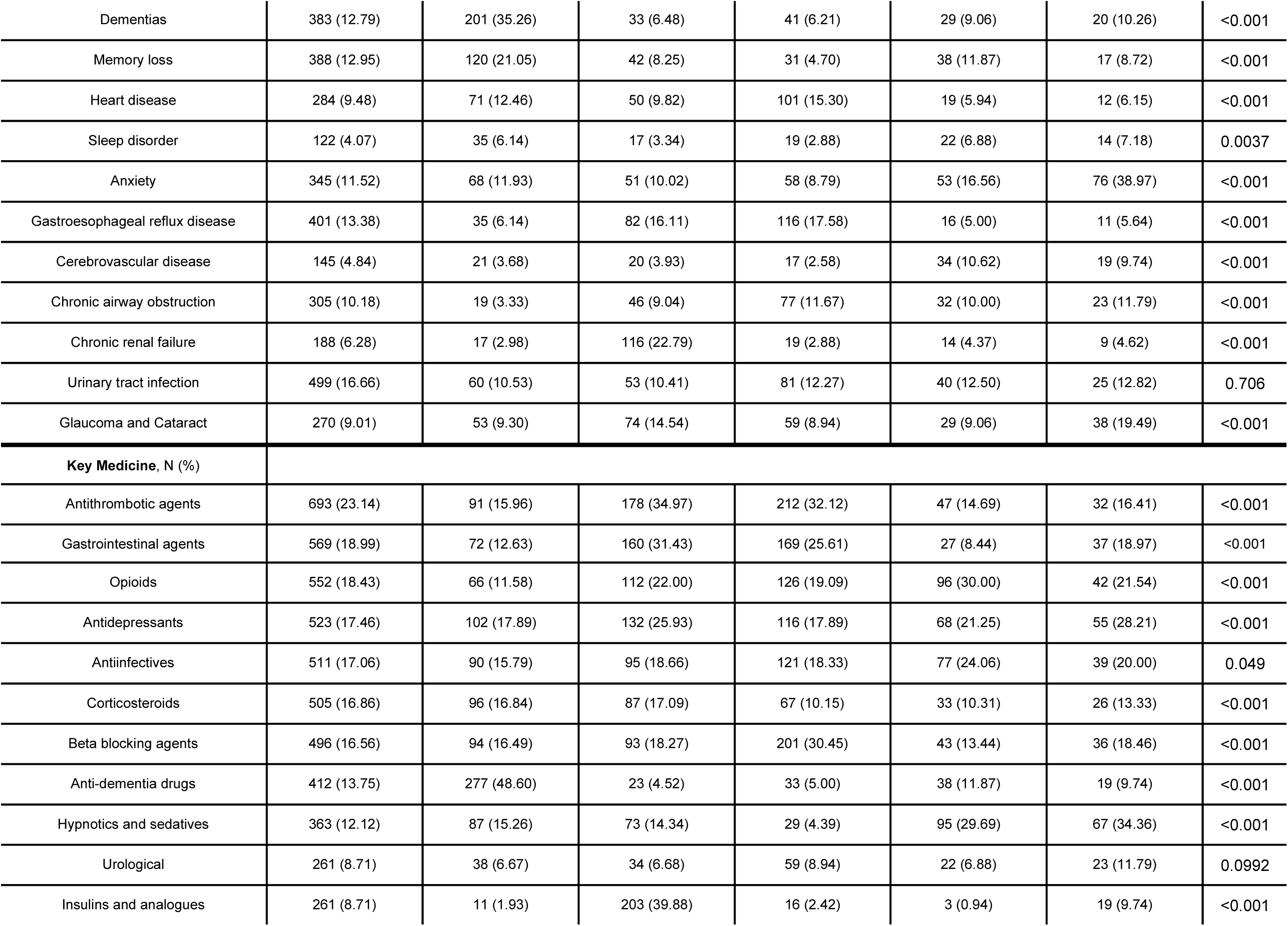
Characteristics (making statistics on MCI onset) of the identified subphenotypes (development cohort). The p-value for sex, race, key comorbidities and medicines are obtained by *χ*^*2*^ test (false discovery rate correction for post-hoc pairwise comparisons in sex and race are in Table 5∼6 in Supplement). The p-value for age and progression time are obtained by Kruskal-Wallis test (with Dunn’s test for post-hoc pairwise comparisons in Table 7∼8 in Supplement).

| Variable |  | Total | Subphenotype I | Subphenotype II | Subphenotype III | Subphenotype IV | Subphenotype V | P-value |
| --- | --- | --- | --- | --- | --- | --- | --- | --- |
| No. of Patient (%) |  | 2995 (100) | 570 (19.03) | 509 (14.79) | 660 (16.99) | 320 (12.31) | 195 (6.51) | -- |
| Age (MCI onset), yr, Median (IQR) |  | 76 [69-83] | 76 [69-83] | 75 [69-82] | 77 [71~83] | 80 [73~87] | 72 [64~78] | <0.001 |
| Sex female, N (%) |  | 1950 (65.11) | 378 (66.32) | 292 (57.37) | 443 (67.12) | 232 (72.50) | 100 (51.28) | <0.001 |
| Race, N (%) | Caucasian | 1580 (52.76) | 365 (64.04) | 201 (39.49) | 335 (50.76) | 203 (63.44) | 92 (47.18) | <0.001 |
|  | African American | 386 (12.89) | 36 (6.32) | 91 (17.88) | 89 (13.48) | 36 (11.25) | 43 (22.05) |  |
|  | Asian | 21 (0.70) | 5 (0.88) | 4 (0.79) | 8 (1.21) | 1 (0.31) | 1 (0.51) |  |
|  | American Indian | 3 (0.10) | 0 (0) | 0 (0) | 1 (0.15) | 0 (0) | 0 (0) |  |
|  | Multiple Race | 28 (0.93) | 3 (0.53) | 1 (0.20) | 10 (1.52) | 0 (0) | 2 (1.03) |  |
|  | Other/Unknown | 977 (36.26) | 161 (28.25) | 212 (41.65) | 217 (32.88) | 80 (25.00) | 57 (29.23) |  |
| Progression time, day, Median (IQR) |  | 871.0 [576.5-1323.5] | 733.5 [508.0-998.0] | 888.0 [607.0~1465.0] | 848.5 [558.0~1391.25] | 807.0 [558.0~1269.0] | 939.0 [681.0~1633.0] | <0.001 |
| 1~2 year, N (%) |  | 1182 (39.47) | 285 (50.0) | 190 (37.33) | 269 (40.76) | 149 (46.56) | 59 (30.26) |  |
| 2~3 year, N (%) |  | 738 (24.64) | 155 (27.19) | 101 (19.84) | 139 (21.06) | 65 (20.31) | 45 (23.08) |  |
| 3~4 year, N (%) |  | 484 (16.16) | 65 (11.40) | 90 (17.68) | 105 (15.90) | 62 (19.38) | 28 (14.36) |  |
| 4~5 year, N (%) |  | 323 (10.78) | 55 (9.65) | 78 (15.32) | 87 (13.18) | 29 (9.06) | 31 (15.90) |  |
| >5 year, N (%) |  | 268 (8.95) | 10 (1.75) | 50 (9.82) | 60 (9.09) | 15 (4.69) | 32 (16.41) |  |
| Key comorbidity, N (%) |  |  |  |  |  |  |  |  |
| Hypertension |  | 1132 (37.79) | 78 (13.68) | 66 (12.97) | 336 (50.91) | 48 (15.00) | 19 (9.74) | <0.001 |
| Hyperlipidemia |  | 495 (16.53) | 61 (10.70) | 82 (16.11) | 176 (26.67) | 32 (10.94) | 24 (12.31) | <0.001 |
| Diabetes |  | 582 (19.43) | 67 (11.75) | 178 (34.97) | 152 (23.03) | 31 (9.69) | 46 (23.59) | <0.001 |

|  |  |  |  |  |  |  |  |
| --- | --- | --- | --- | --- | --- | --- | --- |
| Dementias | 383 (12.79) | 201 (35.26) | 33 (6.48) | 41 (6.21) | 29 (9.06) | 20 (10.26) | <0.001 |
| Memory loss | 388 (12.95) | 120 (21.05) | 42 (8.25) | 31 (4.70) | 38 (11.87) | 17 (8.72) | <0.001 |
| Heart disease | 284 (9.48) | 71 (12.46) | 50 (9.82) | 101 (15.30) | 19 (5.94) | 12 (6.15) | <0.001 |
| Sleep disorder | 122 (4.07) | 35 (6.14) | 17 (3.34) | 19 (2.88) | 22 (6.88) | 14 (7.18) | 0.0037 |
| Anxiety | 345 (11.52) | 68 (11.93) | 51 (10.02) | 58 (8.79) | 53 (16.56) | 76 (38.97) | <0.001 |
| Gastroesophageal reflux disease | 401 (13.38) | 35 (6.14) | 82 (16.11) | 116 (17.58) | 16 (5.00) | 11 (5.64) | <0.001 |
| Cerebrovascular disease | 145 (4.84) | 21 (3.68) | 20 (3.93) | 17 (2.58) | 34 (10.62) | 19 (9.74) | <0.001 |
| Chronic airway obstruction | 305 (10.18) | 19 (3.33) | 46 (9.04) | 77 (11.67) | 32 (10.00) | 23 (11.79) | <0.001 |
| Chronic renal failure | 188 (6.28) | 17 (2.98) | 116 (22.79) | 19 (2.88) | 14 (4.37) | 9 (4.62) | <0.001 |
| Urinary tract infection | 499 (16.66) | 60 (10.53) | 53 (10.41) | 81 (12.27) | 40 (12.50) | 25 (12.82) | 0.706 |
| Glaucoma and Cataract | 270 (9.01) | 53 (9.30) | 74 (14.54) | 59 (8.94) | 29 (9.06) | 38 (19.49) | <0.001 |
| **Key Medicine, N (%)** |  |  |  |  |  |  |  |
| Antithrombotic agents | 693 (23.14) | 91 (15.96) | 178 (34.97) | 212 (32.12) | 47 (14.69) | 32 (16.41) | <0.001 |
| Gastrointestinal agents | 569 (18.99) | 72 (12.63) | 160 (31.43) | 169 (25.61) | 27 (8.44) | 37 (18.97) | <0.001 |
| Opioids | 552 (18.43) | 66 (11.58) | 112 (22.00) | 126 (19.09) | 96 (30.00) | 42 (21.54) | <0.001 |
| Antidepressants | 523 (17.46) | 102 (17.89) | 132 (25.93) | 116 (17.89) | 68 (21.25) | 55 (28.21) | <0.001 |
| Antiinfectives | 511 (17.06) | 90 (15.79) | 95 (18.66) | 121 (18.33) | 77 (24.06) | 39 (20.00) | 0.049 |
| Corticosteroids | 505 (16.86) | 96 (16.84) | 87 (17.09) | 67 (10.15) | 33 (10.31) | 26 (13.33) | <0.001 |
| Beta blocking agents | 496 (16.56) | 94 (16.49) | 93 (18.27) | 201 (30.45) | 43 (13.44) | 36 (18.46) | <0.001 |
| Anti-dementia drugs | 412 (13.75) | 277 (48.60) | 23 (4.52) | 33 (5.00) | 38 (11.87) | 19 (9.74) | <0.001 |
| Hypnotics and sedatives | 363 (12.12) | 87 (15.26) | 73 (14.34) | 29 (4.39) | 95 (29.69) | 67 (34.36) | <0.001 |
| Urological | 261 (8.71) | 38 (6.67) | 34 (6.68) | 59 (8.94) | 22 (6.88) | 23 (11.79) | 0.0992 |
| Insulins and analogues | 261 (8.71) | 11 (1.93) | 203 (39.88) | 16 (2.42) | 3 (0.94) | 19 (9.74) | <0.001 |

**Figure 4.**
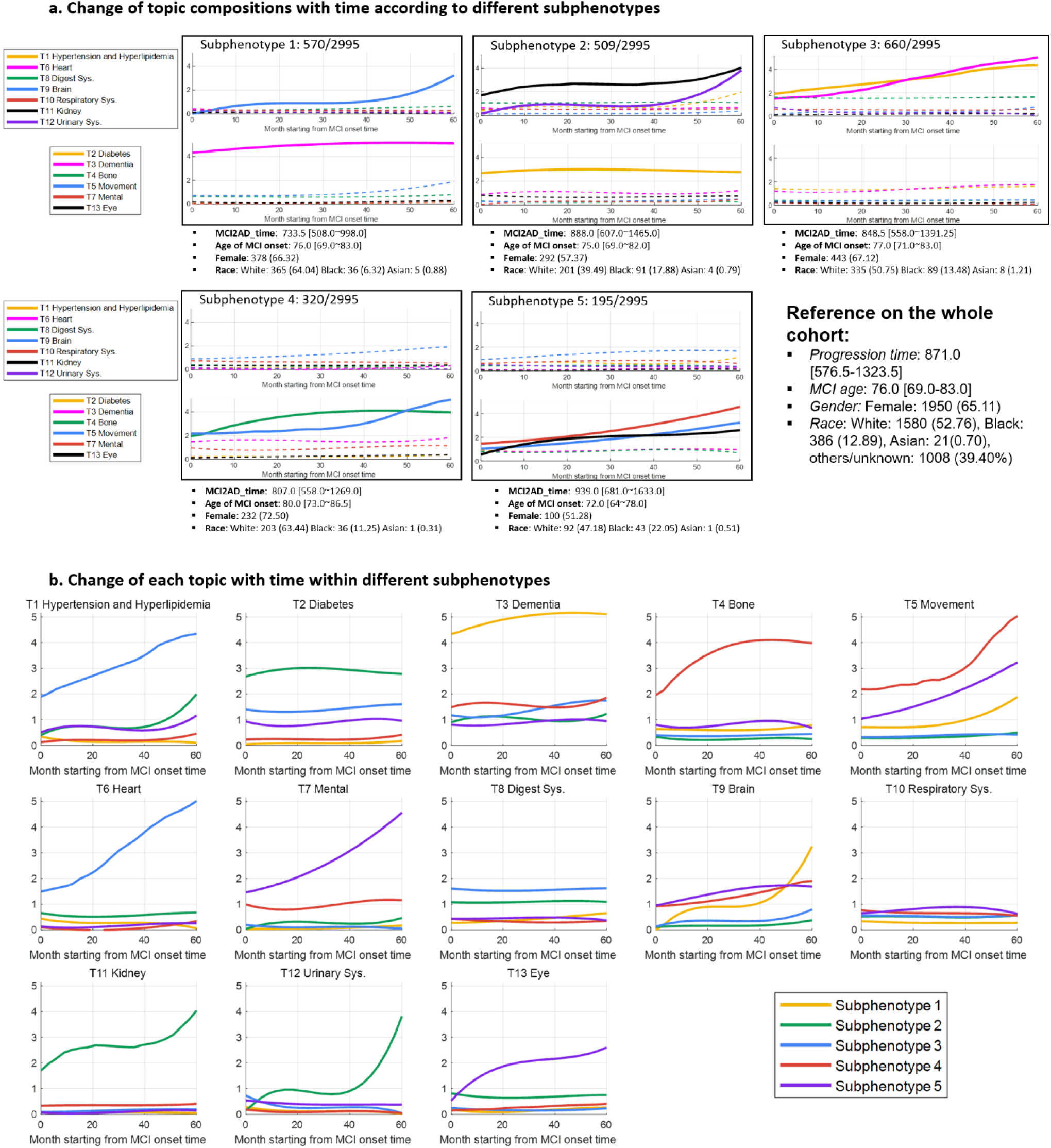
Visualization of longitudinal subphenotypes on the development cohort, which are characterized by the evolution of clinical topic compositions with time. In a, we demonstrate these subphenotypes according to the change of their topic compositions, where major topics whose value exceeds 2 on vertical axis at least once during the entire progression course are showed by solid lines. In b, we illustrate the evolution of each topic within different subphenotypes.

With all these results, in the following we formally characterize each subphenotype.

- **Subphenotype 1** consists of 570 (19.03%) patients with more Caucasian people (64.04%) and has the fastest progressive speed (733.5 [508.0∼998.0] in days; Figure 5). This subphenotype is dominated by T3 (Dementia) and T9 (Brain), where the weight of T3 is stays at a high level during the entire progression, and the value of T9 has clearly increased, especially in the later stage of the progression (Figure 4a). Accordingly, at MCI onset, the percentage of patients having dementia and memory loss is much higher than that in other subphenotypes (Table 2). Meanwhile, during the progression from MCI to AD, more patients would have an increased risk of Parkinson’s disease (PD) and seizures (Supplemental Figure 8), which are closely related with cognitive decline and cerebrovascular problems (*41-43*).
- **Subphenotype 2** consists of 509 (16.99%) patients. Compared to the other subphenotypes, it has more African American (17.88%) and male (42.63%) patients and has the second longest progression time (888.0 [607.0∼1465.0] in days; Figure 5). This subphenotype is dominated by T11 (Kidney), T12 (Urinary system), and T2 (Diabetes), where T2 stays high while T12 and T2 both show increasing trends (Figure 4a). In addition to a high prevalence of CKD and diabetes at MCI onset (Table 2), patients are more likely to have Pneumonia (*44*), Tobacco use disorder (*45*), and anemias (*46*) (Figure 8) during the progression. Moreover, Jain *et al*. (*31*) found that 21% patients with CKD in the U.S. would suffer from a major depressive disorder episode, which could be a potential reason that patients in this subphenotype tend to take antidepressants (Supplemental Figure 9).
- **Subphenotype 3** consists of 660 (22.04%) patients whose demographics and progression time (848.5 [558.0∼1391.25] in days; Figure 5) are close to the cohort level. This subphenotype is characterized by increasing T1 (Hypertension and Hyperlipemia) and T6 (Heart) (Figure 4a), as well as a high level of T8 (Digest system) (Figure 4b). This may cause higher risk of Vitamin-B and Vitamin-D deficiency (Figure 8 in Supplement), which are two common conditions associated with dementia or AD (*47, 48*). Accordingly, the percentage of patients taking Gastrointestinal agents and beta blocking agents is high (Supplemental Figure 9).
- **Subphenotype 4** consists of 320 (10.68%) patients with more female (72.50%) patients and oldest MCI onset age (80.0 [73.0∼86.5] in year) among all subphenotypes. Meanwhile, the progression speed is the second fastest (807.0 [558.0∼1269.0] in days; Figure 5). This subphenotype is characterized by T4 (Bone) and T5 (Movement) whose values stay high during the progression (Figure 4a). This subphenotype has the highest prevalence of Parkinson’s Disease at MCI onset (Supplemental Figure 8), the prevalence of decubitus and hypothyroidism have greatest increase over the progression course, which could be due to movement disorders (*49, 50*). There is also a high rate of opioid prescription in this subphenotype (Supplemental Figure 9) potentially due to the pain caused by problems of bone (T4) and muscle (T5) (Figure 2).
- **Subphenotype 5** consists of 195 (6.51%) patients with more African American (22.05%) patients whose age of MCI onset is the youngest (72.0 [64∼78.0]), and the progression time is the longest (939 [681.0∼1633.0]; Figure 5). This subphenotype is characterized by increasing T7 (Mental), T6 (Movement), and T13 (Eye). Correspondingly, compared with other subphenotypes, we observed the largest increased percentage of patients who suffer from schizophrenia, obesity, bipolar disorder, and fatigue (Supplemental Figure 8), most of which are associated with mental disorders.

### Sex- and race- stratified analysis

We have also conducted sex- and race-stratified analysis for the entire patient cohort and with respect to different subphenotypes. We first checked the difference of MCI onset ages and lengths of progression time breaking down by different race and sex subgroups, and the results are shown in Figure 6. On the entire patient cohort, the age distributions between different sex (Figure 6a) or race (Figure 6b) groups are significantly different but there is no significant difference on the progression time (Figure 6f, 6g). Furthermore, the distributions of age and progression time have significant differences (Figure 6c, 6g) across different subphenotypes (detailed pairwise comparisons are provided in Supplemental Table 5∼8). With further analysis across all subphenotypes, we found that the female patients are typically older than male patients at MCI onset (Figure 6d), and the progression time between patients with different genders have no significant difference (Figure 6f). We have also examined these indices with respect to different races across different subphenotypes (Figure 6e and 6j). Some differences are observed. For example, the age of Caucasian patients is higher than that of African American patients on Subphenotype 1 (p-value<0.001) and Subphenotype 2 (p-value<0.001); the MCI-to-AD progression time of Caucasian patients is longer than that of the African American patients in Subphenotype 2 (p-value<0.001), while shorter than that of African American patients in Subphenotype 3 (p-value=0.031).

**Figure 6.**
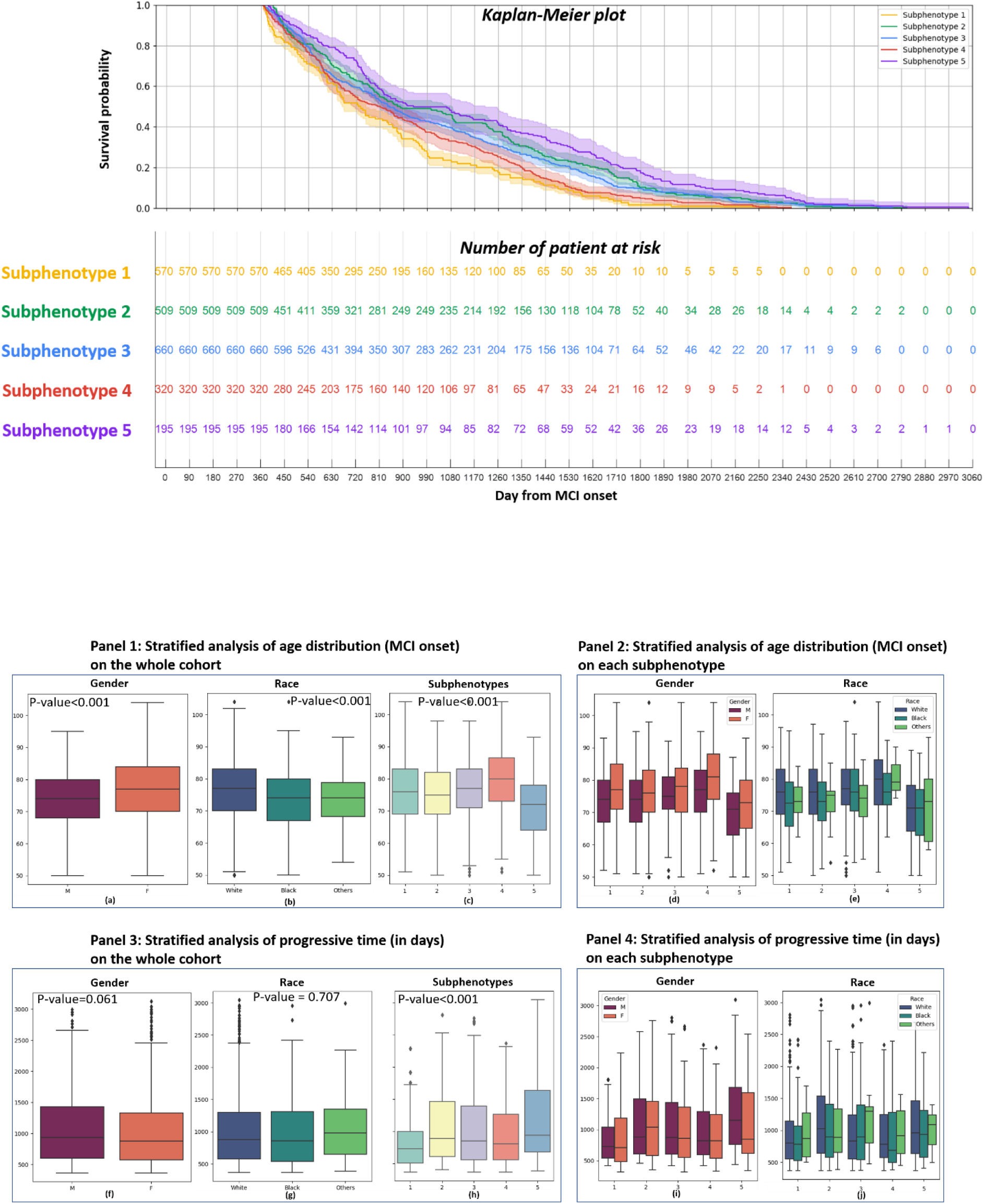
Distributions of age on MCI onset (top, Panel 1 and 2) and progressive time (bottom, Panel 3 and 4) on the development cohort. The Panel 1 and 3 are visualized by different genders ((a) and (f)), races((b) and (g)), and subphenotypes ((c) and (h)) on the whole cohort, while the panel 2 and 4 are visualized by different genders ((d) and (i)) and races ((e) and (j)) on each subphenotype.

There have been prior studies showing gender and race can affect the manifestation and pathophysiology of dementia or AD (*51-54*), thus we did both sex-stratified (Figure 7) and race-stratified analysis (Figure 10 in Supplement) for key clinical components along with their corresponding top-5 diagnosis events (Figure 2). To demonstrate the heterogeneity of disease progression, we show differences of these diagnoses at both MCI and AD onsets for different stratified groups in each subphenotype. One immediate observation is that for each specific topic or disease, there is no consistent observations across all subphenotypes, indicating the complexity of disease progression pattern across different sex- or race- stratified subgroups (*55*). On the other hand, we do have some consistent observations in at least three subphenotypes. For example, at AD onset topics T1 (Hypertension and Hyperlipidemia), T3 (Dementia), T9 (Brain), T11 (Kidney), and T12 (Urinary system) have significant differences between male and female. More concretely, the corresponding diseases of these topics have demonstrated different prevalence between female and male, such as hypertension and hyperlipidemia in T1, cardiovascular diseases in T9, and urinary tract infection in T12 are more prevalent in women, while neurological disorder in T3 and hearing loss in T7 are more prevalent in men, which have also been mentioned in prior studies (*53*). We further noticed that their health condition changes during MCI-to-AD progression are different for different subphenotypes. For example, subphenotype 3 is characterized by increased risk of T1 (Hypertension and Hyperlipidemia) and T6 (Heart disease) (Figure 4a), and the values of these two topics change slowly in other subphenotypes (Figure 4b). These two topics have also demonstrated sex-stratified difference on the progression from MCI to AD in subphenotype 3. Specifically, at MCI onset, only heart failure (belonging to these two topics) prevalence is significantly different between male and female (more prevalent in female), while at AD onset more comorbidities from these two topics stand out. For example, essential hypertension, hyperlipidemia, other chronic ischemic heart disease, and heart failure are all more prevalent in female patients. Similar observations are found in *i)* subphenotype 1 for T9 (Brain) where cerebrovascular diseases and occlusion of cerebral arteries are significantly more prevalent in female patients at AD onset but not at MCI onset, while Epilepsy, recurrent seizure, and convulsions are significantly more prevalent among male patients at MCI onset but not at AD onset; *ii)* subphenotype 2 for T12 (Kidney) where urinary tract infection and retention of urine are significantly more prevalent in female patients at AD onset but not at MCI onset; *iii)* subphenotype 5 for T13 (Eye) where cataract and senile cataract are significantly more prevalent among male patients at AD onset but not at MCI onset. These observations can help us better understand the progression heterogeneity from MCI to AD (*54, 56-59*).

**Figure 7.**
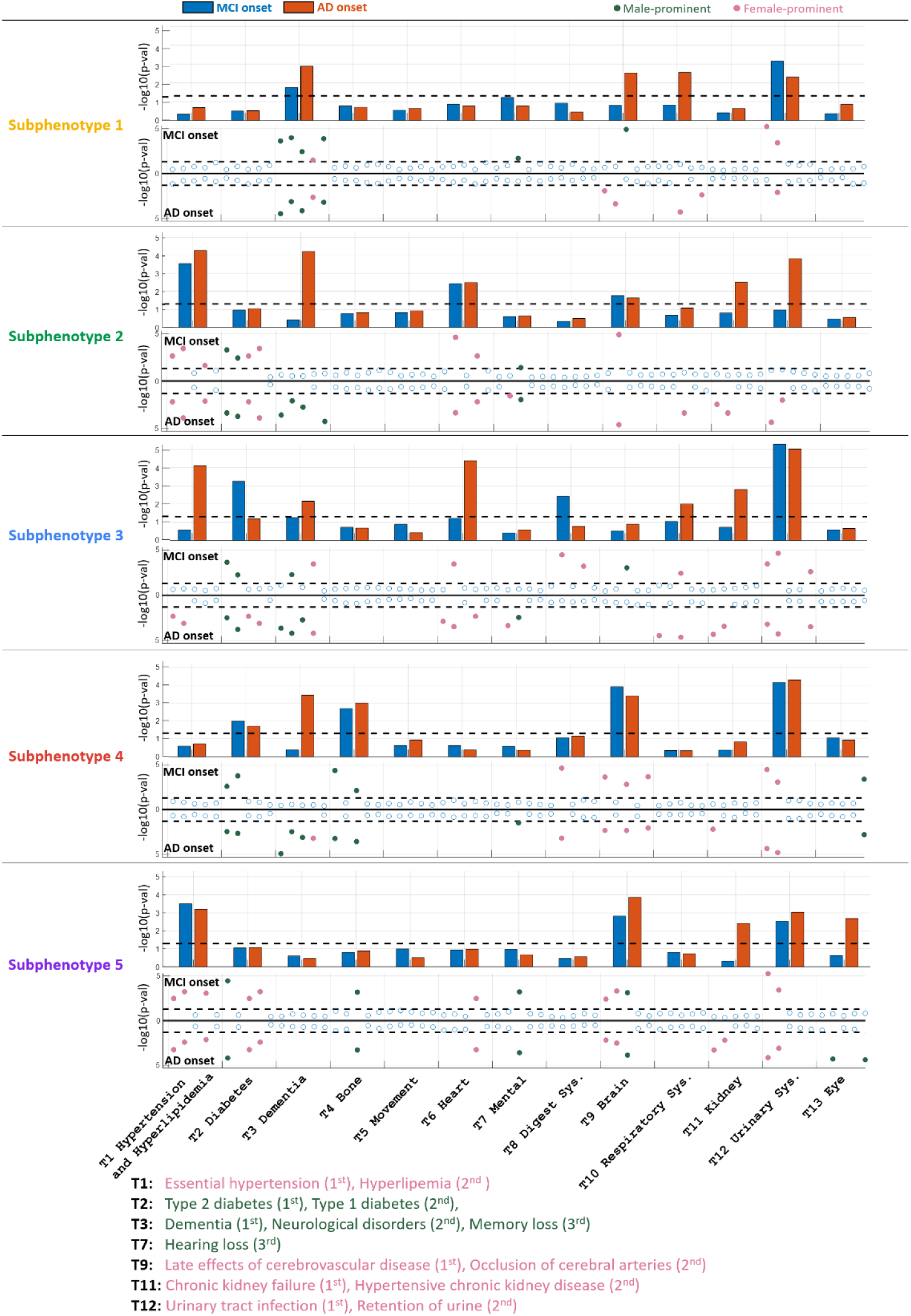
Sex-stratified enrichment analysis of comorbidities on the development cohort. For each subphenotype, the top Bar plot shows the p-value of topic weights on MCI onset (blue) and AD onset (orange) computed using Kruskal-Wallies test. The bottom Miami plot shows the p-value of the top-5 (large weights) diagnosis events in each topic computed by Fisher Exact test on both MCI and AD onset, where some diseases are colored if they are significant in female (pink) or male (green), evaluated by log odds ratio. The black dotted lines in Bar and Miami plots denote p-value=0.05, Below five subphenotypes, names of some key diseases in the topic are listed, where the rank in the bracket denotes the rank of diseases in each topic according to the weights in Figure 2.

### Subphenotype reproducibility

To demonstrate the robustness of these derived progression subphenotypes, we have also reproduced these subphenotypes on the MarketScan and Mount Sinai data, with more details summarized in Table 1.

Using the same procedures, we were able to derive a set of progression subphenotypes whose baseline characteristics at MCI onset are provided in Supplemental Table 9∼13. The top-13 most prevalent clinical topics, topic transition matrix, topic composition and evolutions of different subphenotypes, outcome analysis in terms of encountering AD onset, distributions of age and progressive time, percentage of patients with different comorbidities during progression, and sex-stratified comorbidity analysis on MarketScan are provided in Supplemental Figure 11∼17. Since there is no race information in MarketScan, we performed region-stratified analysis instead and the results are shown in Supplemental Figure 15, from which we can observe that patients in the South region have younger MCI onset age and longer progressive time (compared to the overall statistics on the entire MarketScan data set), which is consistent with the results listed in Table 2 collected from OneFlorida data. The related results obtained from the Mount Sinai dataset are shown in Supplemental Figure 18∼22 and Table 14 with detailed descriptions in Supplemental results. On these two validation cohorts, we identified the same subphenotypes with similar demographics and comorbidity characteristics, illustrating the robustness of our methods.

### Early prediction of the progression subphenotype

Since the identified subphenotypes capture patients’ health condition progression patterns within the full course of MCI-to-AD conversion, early prediction of patients’ subphenotype memberships may largely enhance their clinical implications. To evaluate such predictability of the derived subphenotypes, we conducted two sets of experiments, i.e., internal and external predictions. Internal prediction refers to the procedure of developing and evaluating the predictive model on the same cohort (OneFlorida or MarketScan) through 5-fold cross validation. External prediction is the paradigm of training the predictive model on one cohort (e.g., OneFlorida or MarketScan) and evaluate it on the other cohort (e.g., MarketScan or OneFlorida), which evaluates the ability of model transportability. For both experiments, we trained a logistic regression model based on average topic weights representations learned from DMTM for all visits before the MCI onset (we also tried to add 3-month or 6-month data after MCI onset) to predict the subphenotype assignments (Workflow is in Figure 8a with details in Method). The prediction results measured by accuracy and area under the receiver operator characteristic curve (AUC) are shown in Figure 8b, where we used diagnosis, drug, and procedure events collected from different periods as the input: i) before MCI onset (baseline); ii) until three months after MCI onset; iii) until six months after MCI onset. We observed that with baseline data on development cohort (OneFlorida dataset), the subphenotypes can be predicted with accuracy 63.84%, Micro AUC 78.69%, Macro AUC 77.78%, and the performance can be further improved to accuracy 79.93%, Micro AUC 85.03%, Macro AUC 83.58% with additional data from three months after baseline, and to accuracy 86.04%, Micro AUC 92.72%, Macro AUC 92.39% with additional data from six months after baseline. Similar tendencies were also overserved on the other experimental settings. Moreover, the model performance did not change much (the mean accuracy decreased by 0.79% from MarketScan to OneFlorida while by 5.90% from MarketScan to OneFlorida) when applying to an independent external data set, which demonstrates the robustness of these identified subphenotypes and the transportability of the predictive model.

**Figure 8.**
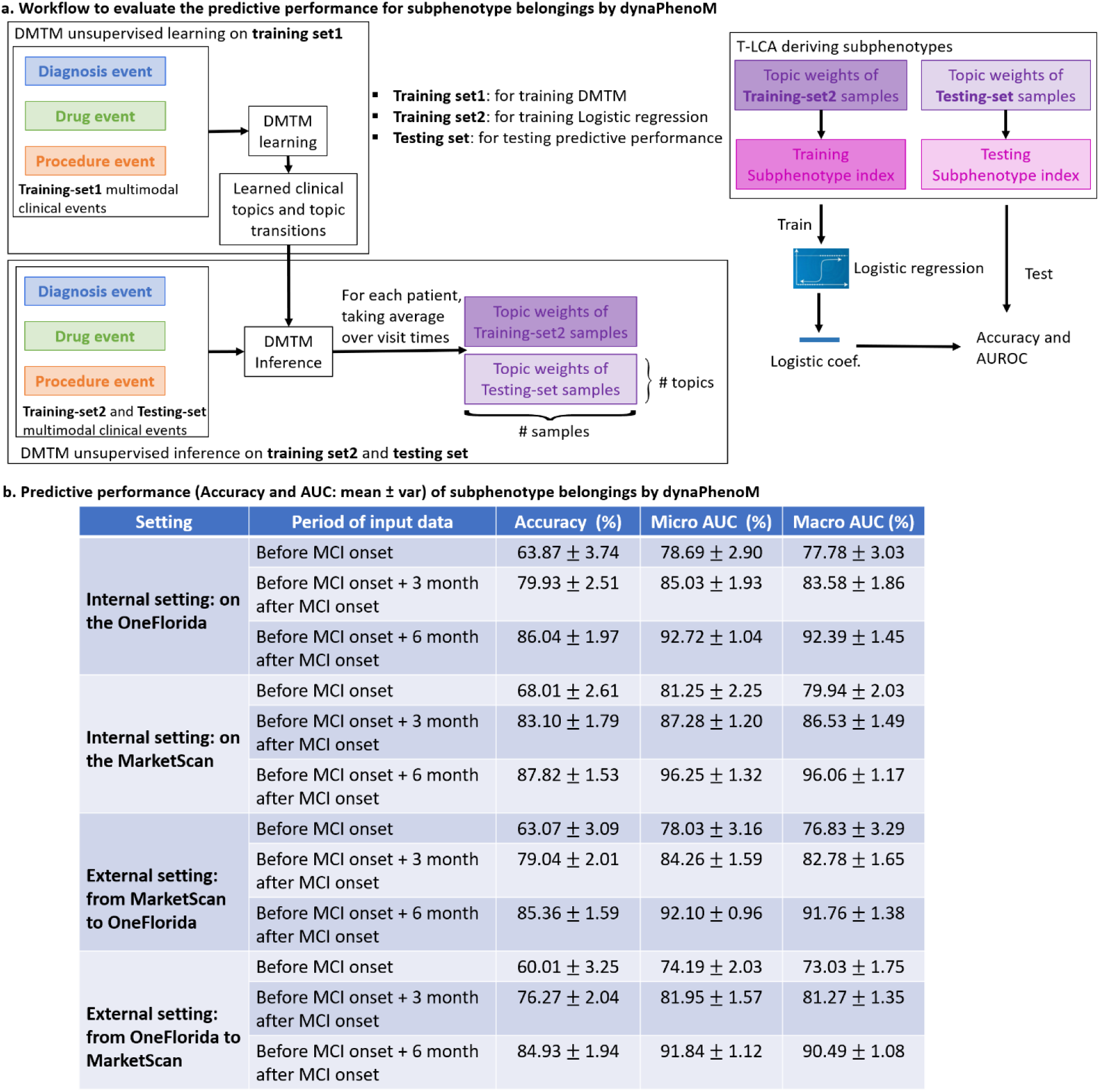
a: Workflow to evaluate the predictive performance for subphenotype belongings. We conducted two sets of experiments: internal and external predictions. Internal prediction refers to the procedure of developing and evaluating the predictive model on a same cohort (OneFlorida or MarketScan) through 5-fold cross validation. External prediction is the paradigm of training the predictive model on one cohort (e.g., OneFlorida or MarketScan) and evaluate it on the other cohort (e.g., MarketScan or OneFlorida). b: Results on different experimental settings.

## Discussion

Identification of clinically-meaningful disease progression subphenotypes can provide invaluable information regarding disease heterogeneity and underlying pathophysiology. In this paper, we developed the dynaPhenoM to achieve this goal using longitudinal patient records. These patient records involve EHR from two independent health systems and a national insurance database. Technically, dynaPhenoM includes two key components, DMTM for extracting interpretable multimodal clinical topics from patient visit vectors and building continuous valued low-dimensional visit representations, and T-LCA to derive progression subphenotypes based on the newly built representations.

To evaluate the effectiveness and robustness of dynaPhenoM, we performed comprehensive analysis on the case of progression from MCI to AD based with the OneFlorida database as development cohort (including 2,995 patients), and the MarketScan database (including 18,805 patients) and the Mount Sinai database (including 689 patients) as validation cohorts. As seen in existing research (*18, 60-62*), AD is highly heterogenous, thus categorizing patients into different clinically coherent subgroups is important for understanding the mechanism of AD and develop stratified medicine. Different from existing works that focus on identifying AD subphenotypes according to specific clinical data (e.g., cognitive assessment score) at AD onset, our study identified progression subphenotypes with a diverse set of clinical events during the progression from MCI to AD. Therefore, we expect our analysis can provide additional insights on the dynamic evolution of the disease.

With dynaPhenoM, we were able to identify 13 important and clinically-meaningful topics and five progression subphenotypes characterized by distinct patient demographics, progression duration, and associated comorbidities. Specifically, **Subphenotype 1** is dominated by topics of brain diseases, includes more Caucasian people, and has the shortest MCI-to-AD progression duration (among the 5 subphenotypes). During the progression from MCI to AD, the patients are with increased risks of PD and seizure. **Subphenotype 2** is composed of more male and African American patients and dominated by the topics of diseases of kidney, urinary system, and diabetes. Patients within this subphenotype have the second longest progression duration and second youngest MCI onset age. More patients would suffer from pneumonia (*44*) and anemias. **Subphenotype 3** is described by the increased risk of topics related to hypertension, hyperlipemia, and heart diseases, which may also be associated with a higher risk of Vitamin-B and Vitamin-D deficiency. **Subphenotype 4** is characterized by high risk of topics about diseases of bone and disorder of movement, with more female. The patients in this subphenotype have the oldest MCI onset age and second shortest progression speed. **Subphenotype 5** includes more African American patients and is dominated by topics of mental, movement, and eye problems. More patients would suffer from schizophrenia, obesity, bipolar disorder, and fatigue, most of which are associated with mental disorders.

We have also performed sex- and race- stratified analysis for each subphenotype on MCI-onset age and progression duration. We found that more females than males with MCI will progress to AD but males tend to have younger MCI or AD onset ages, and the progression durations from MCI to AD are similar for males and females. These trends are observed on both the entire cohort and each of the identified subphenotypes. In addition, we also observed that African American patients tend to have younger MCI onset ages than Caucasian patients (*63*) and have similar progression duration with Caucasian patients. The race-stratified analysis shows different patterns among different subphenotypes. For instance, the difference of MCI onset between Caucasian and African American patients are significant (p-value<0.001) on Subphenotype 1 and 2, but not significant on other three subphenotypes. African American patients have longer progression duration on subphenotype 2 (p-value<0.001) but shorter one on subphenotype 3 (p-value = 0.015). Chen et al. (*64*) pointed out that we need pay more attention about the disparities in dementia prevalence across racial or ethnic groups from the understanding of mechanism of dementia to the drug development.

As suggested by previous clinical studies (*54, 56-59*),studying the differences on the changes of related comorbidities before AD onset can potentially improve our understanding of the underlying disease mechanism and offer informative guide for follow-up treatments. To achieve this goal, we performed further sex- and race- stratified analysis of comorbidities in terms of key clinical topics along with their associated top-5 diagnoses. To better explore the changes during the progression, we did such analysis on both MCI and AD onsets, where the observations on AD onset are similar with those in Tang et.al. (*53*). For example, female AD patients have greater association with hypertension (T1), hyperlipidemia (T1), cardiovascular risk factors (T9), and urinary tract infection (T12) while male AD patients have higher risk in hearing loss (T7) and neurological disorders (T3).

To validate the robustness and reproducibility of the results obtained from dynaPhenoM, we validated our method on another large cohort, where we obtained consistent results as derived from the development cohort. We have also demonstrated that these subphenotypes are predictable at early stage (within 6 months after MCI onset), which further enhances their potential clinical utilities.

There are limitations on the proposed approach. Technically, there are two main modules in dynaPhenoM, DMTM and T-LCA. For DMTM, currently it only considers discrete clinical events including diagnoses, medications and procedures. Actually, equipped with techniques in (*65*), we can further extend DMTM to consider continuous valued events such as lab tests. For T- LCA, it is currently an independent procedure building on top of the representations derived from DMTM. In other words, there is no guarantee that the learned representations can lead to coherent subgroups identified using T-LCA. In the future, we will investigate approaches that can link DMTM and T-LCA in a unified framework so that the topic-based representation and progression subphenotype can be jointly derived. In the study, only structured information in EHR or claims has been explored. For AD, important information is encoded in unstructured data sources, such as neuroimage, clinical notes, and genetic data. We will explore strategies to incorporate these data in future studies as well. Even though, not limited in the case of disease progression from MCI to AD, dynaPhenoM is an efficient data-driven framework to identify progression subphenotypes from longitudinal multimodal clinical data.

## Methods

### Detailed descriptions of cohort definition

#### Development cohorts

We leveraged the patient EHR from OneFlorida Clinical Research Consortium (*21*) to derive the subphenotypes. Detailed inclusion/exclusion cascade is demonstrated in Supplemental Figure 1. All events in each patient’s records, including diagnoses (ICD-9 and ICD-10 codes), drugs (RxCUI and NDC codes) and procedures (CPT codes) from MCI onset to AD onset, were collected in our modeling process. The diagnosis codes were then mapped to 1,643 unique PheCode (*66*) (groups ICD codes into clinically relevant phenotypes). For drugs, the NDC codes were then mapped to RxCUI (ingredient level), and the total number of unique RxCUI codes appeared was 5905. The total number of unique CPT codes appeared was 5129. In our investigation, we have aggregated the patient visits within every 3 months from the MCI onset to AD onset to form the record sequence for each patient.

#### Validation cohorts

We validated the derived subphenotypes on two independent cohorts. The first one is IBM Health MarketScan Commercial Claims database (*22*) for the years 2009 to 2020. This dataset contains about 164 millions of enrollees annually across the US, and these enrollees are nationally representative of the US population with respect to gender, regional distribution, and age, supporting well-powered subgroup analysis. The second one is the patient EHR data from the Mount Sinai Health System which contains five locations in New York City. Similar to the development cohort, we applied a set of inclusion/exclusion criteria (detailed in Figure 2 and 3 in Supplement) on these two datasets, and we finally obtained 18,805 patients from MarketScan, and 698 patients from Mount Sinai for validating subphenotypes. For the MarketScan dataset, the patient diagnosis codes were recorded as ICD-9 and ICD-10, medications were encoded by Generic Product Identifier (GPI) codes, and procedures were encoded with PROCCD codes (mixture of CPT and HCPCS). In the patient cohort we extracted, the diagnosis codes were mapped to 1,750 unique PheCode, while the total unique GPI and PROCCD codes were 4,023 and 8,252. For the Mount Sinai dataset, the diagnosis, medication, and procedure events were encoded with 723 unique PheCode, 3497 unique RxCUI, and 1069 unique CPT codes.

### Dynamic multimodal topic model (DMTM)

We represented the diagnosis, drug, and procedure events as three binary feature vectors. Due to the large vocabulary size (total unique codes) of three modalities, the original feature vectors are high-dimensional and sparse (*9*), which makes efficient clustering difficult. Therefore, we proposed a novel probabilistic model, dynamic multimodal topic model (DMTM), to extract low-dimensional continuous features from original high-dimensional binary vectors. The new features extracted from DMTM are not only beneficial for the following derivation of subphenotypes, but also explainable for the exploration of subphenotypes.

As shown in Figure 1, DMTM models longitudinal multimodal clinical events from longitudinal patient events as a latent generative process, from the first visit to the last, whose specific notations are provided in Supplemental Table 15. After collecting all patient records, the *m*-th modality of *n*-th patient at *t*-th visit can be represented as a binary vector 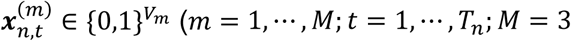 in our current case including diagnosis, medication, and procedure events), where *V*_*m*_ represents the total unique clinical events (vocabulary size) in *m*-th modality, and *T*_*n*_ is the total number of visits for the *n*-th patient. Suppose that there are *k* latent clinical topics and each contains *M* different types of topics corresponding to different modalities denoted as 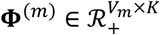, in which the *k*-th column, 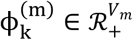, represents *k*-th topic, a distribution over all events (unique codes) in *m*-th modality. DMTM assumes that 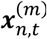 is composed of *k* topics with 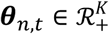 being the topic weight vector (mixture composition) shared by all modalities. Therefore, *k*-th topic in different modalities 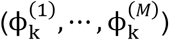 are highly correlated, forming as *k*-th clinical topics shown in Figure 2. To model the transition pattern of topic weights between two successive visits, DMTM introduces a transition matrix 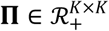, where each element, *Π*_*ij*_, represents the probability of transition from *i*-th topic to the *j*-th topic. Formally, the generative process of DMTM can be written as:

- Topic weights: *θ*_*n*,1_ ∼ *Gamma* (r, 1), *θ*_*n, t*_ ∼ *Gamma* (*τ*_0_Π*θ*_*n,t*−1_, *τ*_0_), *t* = 2, …, *T*_*n*_
- Latent clinical topics and transition matrix: 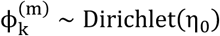, *Π*_*kk*_ ∼ *Dirichlett*(*ν*_1_*ν*_*k*_, …, ξv_*k*_, …, v_K_v_*k*_), *k* = 1, …, *k*
- Intermediate variables 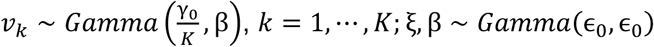
- EHR clinical events which are represented as:

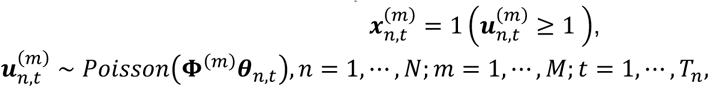

where, *Gamma, Diri*c*hlet*, and *Poi*sso*n* denote the Gamma, Dirichlet, and Poisson distribution, respectively; **1**(·) is an indicator function representing that 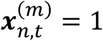 if 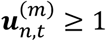, and 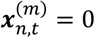 if 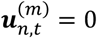. This function is called Bernoulli-Poisson link (*67*), whose mathematical motivation is that after transforming a binary-modeling problem (clinical event happens or does not happen in this visit) into a count modeling one, one is readily equipped with a rich set of statistical tools developed for count data analysis using the Poisson and negative binomial distributions.

There are four positive hyperparameters to be set by users: *τ*_0_, γ_0_, η_0_, ϵ_0_. In our setting, we set them as *τ*_0_ = 1, γ_0_ = 100, η_0_ = 0.01, ϵ_0_ = 0.1. We developed the Gibbs sampling to estimate the posterior of all variables (in Supplement). Here, we only showed the posterior of topic weights ***θ***_*n, t*_ to explain why DMTM can alleviate the problem of missing events in longitudinal patient records.

### Robustness of DMTM for missing events

The posterior of *θ*_*t*_ (without loss of generality, we ignore the patient index *n*) at *t*-th visit is a Gamma distribution as

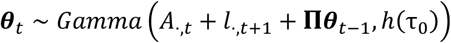

where, from a mathematical view, Π*θ*_*t*−1_ transforms the information from the prior visit (*t* − 1), *A*·,_*t*_ represents the information of current visit (*t*), and *l*·,_*t*+1_ transforms the information from the next visit (*t* + 1). In other words, when inferring the topic weight vector of *t*-th visit (*θ*_*t*_), DMTM not only uses the clinical events from the current visit, but also looks forward and backward to use the information from neighboring visits. As a result, even if some events are missed at current event, DMTM may recall them by relating events from neighboring visits.

### Measuring similarity of multimodal clinical events on the topic space

As discussed before, DMTM learns the topic matrix of multimodal clinical events, represented as 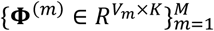. Illustrated in the workflow in Supplemental Figure 7a, we can regard each row from Φ^(*m*^) as a projection of clinical events to the inferred shared topic embeddings space, which enables the discover of associations among events (*9*). For example, we obtained the embeddings of two events as ***e***_1_ and ***e***_2_ from topic matrices. We calculated the cosine similarity between them as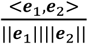, where < ·,· > denotes the inner product of two embeddings and || · || denotes the norm of vector. Thus, in Supplemental Figure 7b, given a query clinical event, we showed the top-10 related (most similar) diagnosis events, top-5 related medication and procedure events.

### Identify key topics

We found that using all topics to learn subphenotype is still inefficient, and the interpretation is not intuitive. To solve this problem, before leaning subphenotypes, on each dataset, we firstly identified key topics on each dataset from all topics. Since topic weight vector *θ*_*n,t*_ can describe the importance of each topic in describing the observed data, in the following, we introduced how to identify key topics according to the topic weight vector.

Specifically, for *k*-th topic, we use 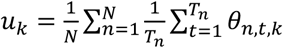 to represent the mean topic usage since we take average over all patients (index by *n*) and all visits (index by *t*). After that, for the *k*-th topic, we define the percentage of mean topic usage as 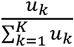. If 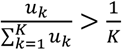, we consider this topic as a key topic since threshold 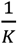 (*k* is the total number of topics) is the mean usage over all topics. The results of identifying key topics on three cohorts are provided in Supplementary Figure 5.

### Time-aware latent class analysis (T-LCA)

Most existing works in clinical research about deriving longitudinal subphenotypes were implemented using latent class analysis (group based trajectory modeling) (*68*) or dynamic time warping (*69*), which often regarded visit times rather than calendar time as time stamps. However, such methods ignored the fact that the time interval between two visits may be irregular, varying from days to months, which is important for clinical study since it embeds the progressive speed of diseases. To this end, in this paper, we introduced time-aware latent class analysis (T-LCA).

Specifically, the new features extracted by DMTM are topic weight vectors denoted as 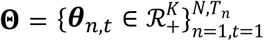 (note that here we use *k* to represent the number of key topics), T-LCA models the data likelihood of Θ by a mixture of Gaussian distribution as:

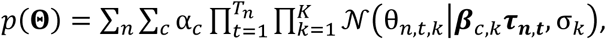

where, 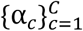 are the mixture coefficients with *C* being the number of subphenotypes, and the mean ***β***_*c,k*_*τ*_***n***,***t***_ is defined as:

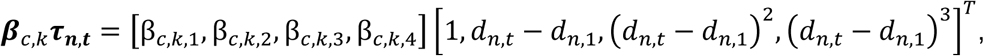

where, *d*_*n*,1_ is the calendar time of starting point (such as MCI onset in our case) for *n*-th patient; *d*_*n,t*_ is the calendar time of *t*-th visit for *n*-th patient. In other words, *d*_*n,t*_ − *d*_*n*,1_ models the calendar time interval from the starting point to every visit, which embeds the natural time progression. This is the reason why we call our proposed new type of LCA as time-aware LCA. To learn the parameters 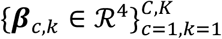 and 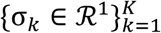, and infer the subphenotype belonging for each patient, we use the Expectation–Maximization (EM) algorithm (64) whose details are in Supplement.

As shown in Figure 4, we used *y*-axis to represent the mean (over patients in corresponding subphenotype) number of diagnosis events (for one topic) whose probabilities of occurrence are larger than 0.5. Here we provided more details to illustrate it.

As shown in the generative process of DMTM for multimodal longitudinal patient events, we used the Bernoulli-Poisson link to transform the binary-modeling problem (clinical event happens or does not happen in this visit) into a count modeling, which enables us to readily employ a rich set of statistical tools developed for count data to do data mining. If we marginalized out the auxiliary variable 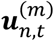, we obtained a Bernoulli random variable as

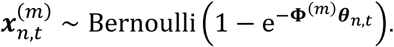

According to the property of Bernoulli distribution, the mean is 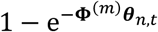, where 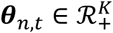 represents the topic weight vector (the total number of topics is *k*) of *n*-th patient at visit of time *t*.

Assume that one subphenotype has *N*′ patients. In the visualization of subphenotypes (Figure 4), for *k*-th topic at calendar time *t*, we firstly calculate the mean of corresponding topic weight as 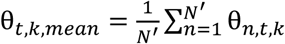, and then obtain the 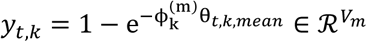 (note that *y*_*t, k*_ ∈[0,1] since 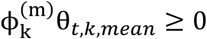). Each value in *y*_*t, k*_ represents the mean probability (decided by *k*-th topic) of each clinical event appearing in calendar time *t*. We count the number of clinical events whose appearing probability is larger than 0.5 as the value of *y* axis in Figure 4. In other words, the larger the value of *y*-axis is, the more diagnosis events from the corresponding topic will occur, the higher risk of having these diseases.

### Prediction of subphenotype assignment

For dynaPhenoM, we proposed two experimental settings to evaluate its performance on prediction of subphenotype assignments, which can further illustrate the robustness and generalizability of our method. One of the settings is to train and evaluate the performance in one dataset (internal) by five-fold cross validation. The other setting is to evaluate using two datasets (external) by training models on one dataset and then testing the trained model on another dataset. Specifically, as shown in Figure 8a, for both settings, we firstly split training set as training set1 (60%) and training set2 (40%). We collected all longitudinal patient events and then trained the DMTM on training set1. After training, we obtained the clinical topics 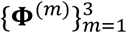 and topic transition matrix ∏. Given well-learned 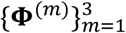 and ∏, regarding training set2 and testing set as input, we used DMTM to infer their topic weight vectors represented as 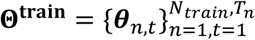 and 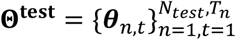, respectively. Having obtained the topic weight vector of both training-set2 and testing samples, we used T-LCA to derive the subphenotype belongings of all samples (*N*_*train*_ + *N*_*test*_). Regarding the mean over time of topic weights as the features for each patient that means 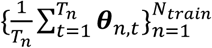 and 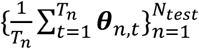, we trained a logistic regression model on training set2 and then tested the performance on testing set. From Table 1 and Supplementary table 16, we observed distribution shift of basic characteristics and the data between cohorts of MarketScan and OneFlorida. From Figure 8, we found such distribution shift does not affect performance too much, especially when trained on MarketScan (larger dataset) and tested on OneFlorida. Since there are total five subphenotypes (multiple classes), for AUC results, we provide both micro-AUC and Macro-AUC.

## Supporting information

All supplements for dynaPhenoM including figures and tables

## Data Availability

All data produced in the present study are available upon reasonable request to the authors

## Data availability

The real-world data analyzed in this article were provided by OneFlorida Clinical Research Consortium (OneFlorida dataset), IBM MarketScan Research Databases (MarketScan dataset), and the Mount Sinai Health System (Mount Sinai dataset). These data are not publicly accessible due to restricted user agreement. Requests for access to OneFlorida dataset should be submitted to and approved by OneFlorida Clinical Research Consortium (https://www.ctsi.ufl.edu/ctsa-consortium-projects/oneflorida/); access to MarketScan dataset can be obtained by contacting IBM (https://www.ibm.com/products/marketscan-research-databases/databases); access to Mount Sinai dataset can be sent to Benjamin. However, we have provided a toy data incorporated in the open-source tool we released for understanding the method.

## Code availability

The implementation of the proposed DMTM and T-LCA in Python and MATLAB are publicly available at https://github.com/haozhangWCM/dynaPhenoM.

## Acknowledgements

FW acknowledges the support from NIH RF1AG072449 and NSF 1750326. JC is supported by National Center for Advancing Translational Sciences (NCATS) Grant (UL1TR001998). JB is supported by National Institutes of Health (NIH) Grants (R21 AG068717, R01CA246418-02S1, R56AG069880). FC is supported by NIH U01AG073323, R01AG066707 and R56AG074001.

