## Supplementary material for "dynaPhenoM: Dynamic Phenotype Modeling from Longitudinal Patient Records Using Machine Learning": All supplements for dynaPhenoM including figures and tables

#### Abstract

Identification of clinically meaningful subphenotypes of disease progression can facilitate better understanding of disease heterogeneity and underlying pathophysiology. We propose a machine learning algorithm, termed dynaPhenoM, to achieve this goal based on longitudinal patient records such as electronic health records (EHR) or insurance claims. Specifically, dynaPhenoM first learns a set of coherent clinical topics from the events across different patient visits within the records along with the topic transition probability matrix, and then employs the time-aware latent class analysis (T-LCA) procedure to characterize each subphenotype as the evolution of these learned topics over time. The patients in the same subphenotype have similar such topic evolution patterns. We demonstrate the effectiveness and robustness of dynaPhenoM on the case of mild cognitive impairment (MCI) to Alzheimer's disease (AD) progression on three patient cohorts, and five informative subphenotypes were identified which suggest the different clinical trajectories for disease progression from MCI to AD.

### Methods

#### Monte Carlo Markov Chain inference via the Gibbs sampling for DMTM

Our proposed DMTM is one of important modules in dynaPhenoM. DMTM learns the clinical topics and extracts the new low-dimensional continuous feature based on these clinical topics. Such new features are not only clinically interpretable, but also efficient for deriving longitudinal subphenotypes. To train the DMTM, we introduce Monte Carlo Markov Chain (MCMC) inference by Gibbs sampling. During the inference, we need the following probability properties.

**Property 1** (**P1**) If $y_{\cdot} = \sum_{n=1}^{N} y_n$ (in the following, we use · to represent the summation over one dimension), where $y_n \sim Poisson(\psi)$ are independent Poisson-distributed random variables. Then, we have:

$$(y_1, \cdots, y_n) \sim Multi\left(y_{\cdot}, \frac{\psi_1}{\sum_{n=1}^{N} \psi_n}, \cdots, \frac{\psi_N}{\sum_{n=1}^{N} \psi_n}\right),\ y_{\cdot} \sim Poisson(\textstyle\sum_{n=1}^{N} \psi_n) \quad (1)$$

where, $Multi$ represents the multinomial distribution.

**Property 2** (**P2**) $y \sim Poisson(c\psi)$, where $c$ is a constant, and $\psi \sim Gamma(a, b)$ then $y \sim NB\left(a, \frac{c}{c+b}\right)$ is a negative binomial-distributed random variable. We can equivalently parameterize it as $y \sim NB\big(a, g(\rho)\big)$, where $g(\rho) = 1 - exp(-\rho)$ is the Bernoulli-Poisson link and $\rho = log\left(1 + \frac{c}{b}\right)$.

**Property 3** (**P3**) If $y \sim NB\big(a, g(\rho)\big)$, and $l \sim CRT(y, a)$ is a Chinese restaurant table distributed random variable, then $y$ and $l$ are equivalently jointly distributed as $y \sim SumLog\big(l, g(\rho)\big)$ and $l \sim Poisson(a\rho)$.

Given these three properties, next, we introduce the MCMC inference via Gibbs sampling for DMTM. Without loss of generalizability, in the next analysis, we ignore the sample index $n$.

Firstly, according to the Bernoulli-Poisson link in the generative process of clinical event, we can sample the auxiliary latent counts $\boldsymbol{u}_{n,t}^{(m)}$ by its conditional posterior:

$$\boldsymbol{u}_t^{(m)} | - \sim \boldsymbol{x}_t^{(m)} Poisson_{+}(\boldsymbol{\Phi}^{(m)} \boldsymbol{\theta}_t), \quad (2)$$

where, $Poisson_{+}$ represents the truncated Poisson distribution.

Then we can augment each latent count observations of three modalities $\{\boldsymbol{u}_t^{(m)}\}_{m=1}^{3}$ into summation of $K$ (the number of topics) latent counts as

$$u_{v,t}^{(1)} = \textstyle\sum_{k=1}^{K} A_{vkt}^{(m)},\ A_{vkt}^{(m)} \sim Poisson\left(\Phi_{vk}^{(m)} \theta_{kt}\right), \quad (3)$$

where $v$ is the index of the clinical event. Applying P1, for different $v$ and $t$, we can sample $\{A_{vkt}^{(m)}\}_{k=1}^{K}$ by multinomial distribution as

$$\left(A_{v1t}^{(m)}, \cdots, A_{vKt}^{(m)}\right) \sim Multi\left(u_{v,t}^{(m)}, \frac{\Phi_{v1}^{(m)} \theta_{1t}}{\sum_{k=1}^{K} \Phi_{vk}^{(m)} \theta_{kt}}, \cdots, \frac{\Phi_{vK}^{(m)} \theta_{Kt}}{\sum_{k=1}^{K} \Phi_{vk}^{(m)} \theta_{kt}}\right). \quad (4)$$

We define $A_{\cdot kt}^{(m)} = \sum_{v=1}^{V_m} A_{vkt}^{(m)}$, according to (3) and $\sum_{v=1}^{V_m} \Phi_{vk}^{(m)} = 1$ (each topic follows Dirichlet distribution), then we have

$$A_{\cdot kt}^{(m)} \sim Poisson\left(\sum_{v=1}^{V_1} \Phi_{vk}^{(m)} \theta_{kt}\right) = Poisson(\theta_{kt}) \ .$$

We start with $\theta_T$ at the last time point $T$. As none of the other time-step factors depend on it in their priors, we can compute the sum of three latent counts $\{A_{\cdot kT}^{(m)}\}_{m=1}^{3}$ as

$$A_{\cdot kt}^{(1+2+3)} = A_{\cdot kt}^{(1)} + A_{\cdot kt}^{(2)} + A_{\cdot kt}^{(3)} \sim Poisson(3\theta_{kT}). \tag{5}$$

According to the generative process of DMTM, since the prior of $\boldsymbol{\theta}_T$ is $\boldsymbol{\theta}_T \sim Gamma\left(\tau_0 \Pi \boldsymbol{\theta}_{n,t-1}, \tau_0\right)$, applying P2 with Equation (5), we can marginalize out $\theta_{kT}$ to obtain

$$A_{\cdot kt}^{(1+2+3)} \sim NB\left(\sum_{k_i=1}^{K} \pi_{kk_i} \theta_{k(T-1)}, g(\zeta)\right), \ g(\zeta) = log\left(1 + \frac{3}{3+1}\right). \tag{6}$$

In order to marginalize out $\boldsymbol{\theta}_{T-1}$, we introduce an auxiliary variable following the Chines restaurant table (CRT) distribution as

$$e_{kT} \sim CRT\left(A_{\cdot kt}^{(1+2+3)}, \sum_{k_i=1}^{K} \pi_{kk_i} \theta_{k(T-1)}\right). \tag{7}$$

According to P3, we re-express the joint distribution over $A_{\cdot kt}^{(1+2+3)}$ in (6) and $e_{kT}$ in (7) as

$$A_{\cdot kt}^{(1+2+3)} \sim SumLog\left(e_{kT}, g(\zeta)\right), \ e_{kT} \sim Poisson\left(\zeta \times \sum_{k_i=1}^{K} \pi_{kk_i} \theta_{k(T-1)}\right).$$

Therefore, $e_{kT}$ can be seen as the count propagating from $T$ to $T-1$. Repeating the process all the way back to $t = 1$, we are able to marginalize out all gamma latent variables $\{\boldsymbol{\theta}_t\}_{t=1}^{T}$, and for each iteration, we provide closed-form conditional posteriors for all learnable parameters as follows.

- Sample latent count observations:

$$\boldsymbol{u}_t^{(m)} | - \sim \boldsymbol{x}_t^{(m)} Poisson_{+}(\boldsymbol{\Phi}^{(m)} \boldsymbol{\theta}_t)$$

- Sample decomposed latent counts:

$$\left(A_{v1t}^{(m)}, \cdots, A_{vKt}^{(m)}\right) \sim Multi(u_{v,t}^{(m)}, \frac{\Phi_{v1}^{(m)} \theta_{1t}}{\sum_{k=1}^{K} \Phi_{vk}^{(m)} \theta_{kt}}, \cdots, \frac{\Phi_{vK}^{(m)} \theta_{Kt}}{\sum_{k=1}^{K} \Phi_{vk}^{(m)} \theta_{kt}}),$$

- Sample clinical topics:

$$\phi_k^{(m)} | - \sim \text{Dirichlet}(\eta + \sum_{n=1,t=1}^{N,T_n} A_{1kt,n}^{(m)}, \cdots, \eta + A_{V^m kt,n}^{(m)}),$$

- Sample the propagation latent counts from $t = T$ to $t = 1$:

$$l_{\cdot kT} \sim Poisson(\tau_0 \theta_{kT})$$

$$l_{kt\cdot} \sim CRT\left(A_{\cdot kt}^{(1+2+3)} + l_{\cdot k(t+1)}, \tau_0 \sum_{k_i=1}^{K} \pi_{kk_i} \theta_{k_i t}\right),$$

$$(l_{kt1}, \cdots, l_{ktK}) \sim Multi\left(u_{v,t}^{(m)}, \frac{\pi_{k1} \theta_{1(t-1)}}{\sum_{k_i=1}^{K} \pi_{kk_i} \theta_{k_i(t-1)}}, \cdots, \frac{\pi_{kK} \theta_{K(t-1)}}{\sum_{k_i=1}^{K} \pi_{kk_i} \theta_{k_i(t-1)}}\right),$$

- Sample transformation matrix:

$$\pi_k \sim \text{Dirichelt}\left(v_1 v_k + \sum_{n=1}^{N} \sum_{t=1}^{T_n} l_{1kt,n}, \cdots, \xi v_k + \sum_{n=1}^{N} \sum_{t=1}^{T_n} l_{kkt,n}, \cdots, v_K v_k + \sum_{n=1}^{N} \sum_{t=1}^{T_n} l_{Kkt,n}\right)$$

- Sample topic weight vector:

$$\theta_{k,1} \sim Gamma\left(A_{\cdot k1}^{(1+2+3)} + l_{\cdot k2} + \tau_0 v_k, 2\tau_0 + 1\right),$$

$$\theta_{k,t} \sim Gamma\left(A_{\cdot kt}^{(1+2+3)} + l_{\cdot k,t+1} + \tau_0 \sum_{k_2=1}^{K} \pi_{kk_2}\theta_{k_2,t-1}, 2\tau_0 + 1\right).$$

The Python or MATLAB scripts are provided in our open-source tool to perform the DMTM sampling procedure.

**Expectation Maximization for T-LCA**

After extracting the topic weight vector as the new features for each visit, we used this vector to derive the subphenotypes by our proposed T-LCA. Similar to the original LCA, T-LCA can be learned via expectation maximization described as follows.

Specifically, the new features extracted by DMTM are topic weights denoted as $\boldsymbol{\Theta} = \{\boldsymbol{\theta}_{n,t}\}_{n=1,t=1}^{N,T_n}$, T-LCA models the data likelihood by a mixture of Gaussian distribution as:

$$p(\boldsymbol{\Theta}) = \sum_n \sum_c \alpha_c \prod_{t=1}^{T_n} \prod_{k=1}^{K} \mathcal{N}\left(\theta_{n,t,k} \middle| \boldsymbol{\beta}_{c,k}\boldsymbol{\tau}_{\boldsymbol{n},\boldsymbol{t}}, \sigma_k\right).$$

**E-step.** In E-step, we infer the probability of each sample belonging to each subphenotype:

$$\omega_{n,c} = \frac{\alpha_c \prod_{t=1}^{T_n} \prod_{k=1}^{K} \frac{1}{\sigma_k\sqrt{(2\pi)}} exp\left(-0.5\left(\frac{\theta_{n,t,k} - \boldsymbol{\beta}_{c,k}\boldsymbol{\tau}_{\boldsymbol{n},\boldsymbol{t}}}{\sigma_k}\right)^2\right)}{\sum_{c=1}^{C} \alpha_c \prod_{t=1}^{T_n} \prod_{k=1}^{K} \frac{1}{\sigma_k\sqrt{(2\pi)}} exp\left(-0.5\left(\frac{\theta_{n,t,k} - \boldsymbol{\beta}_{c,k}\boldsymbol{\tau}_{\boldsymbol{n},\boldsymbol{t}}}{\sigma_k}\right)^2\right)}$$

Then, calculating the summations of probabilities at every local cite as:

$$\eta_c = \sum_{n=1}^{N} \omega_{n,c}.$$

**M-step.** The overall objective including all samples from all local cites in M step can be written as

$$Q = \sum_{n=1}^{N} \sum_{c=1}^{C} \omega_{n,c} \, log \frac{\alpha_c p\left(\boldsymbol{\theta}_{\boldsymbol{n}} \middle| \{\boldsymbol{\beta}_{c,k}, \sigma_k\}_{k=1}^{K}\right)}{\omega_{n,c}}.$$

We need to maximize the $Q$ with respect to all parameters. Firstly, we derive how to update the $\{\boldsymbol{\beta}_{c,k} \in R^4\}_{c=1,k=1}^{C,K}$, where we take one of the elements in $\boldsymbol{\beta}_{c,k}$, $\{\beta_{c,k,i}\}_{i=1}^{4}$, as an example to illustrate. Let $\frac{\partial Q}{\partial \beta_{c,k,i}} = 0$, we have:

$$\beta_{c,k,i} = \frac{\sum_{n=1}^{N} \sum_{t=1}^{Tn} \omega_{n,c}\tau_{n,t,i}\left(\theta_{n,t,k} - \boldsymbol{\beta}_{c,k}\boldsymbol{\tau}_{\boldsymbol{n},\boldsymbol{t}} + \beta_{c,k,i}\tau_{n,t,i}\right)}{\sum_{n=1}^{N_l} \sum_{t=1}^{Tn} \omega_{n,c}\left(\tau_{n,t,i}\right)^2},$$

where, $\tau_{n,t,i} = \left(d_{n,t} - d_{n,1}\right)^{i-1}$. Calculating the following variables:

$$A_{c,k,i} = \sum_{n=1}^{N} \sum_{t=1}^{T_n} \omega_{n,c}\tau_{n,t,i}\left(\theta_{n,t,k} - \boldsymbol{\beta}_{c,k}\boldsymbol{\tau}_{\boldsymbol{n},\boldsymbol{t}} + \beta_{c,k,i}\tau_{n,t,i}\right),$$

$$B_{c,k,i} = \sum_{n=1}^{N} \sum_{t=1}^{T_n} \omega_{n,c} \left(\tau_{n,t,i}\right)^2.$$

And then, updating $\beta_{c,k,i}$ on computational center as:

$$\beta_{c,k,i} = \frac{A_{c,k,i}}{B_{c,k,i}}.$$

Next, we introduce how to update the variance. First step is calculating the aggregated variables as:

$$D_k = \sum_{n=1}^{N} \sum_{t=1}^{T_n} \omega_{n,c} \left(\theta_{n,t,k} - \boldsymbol{\beta}_{c,k} \boldsymbol{\tau}_{\boldsymbol{n},\boldsymbol{t}}\right)^2,$$

$$E_k = \sum_{n=1}^{N} \sum_{t=1}^{T_n} \omega_{n,c} T_n.$$

And then, updating $\sigma_k$ on computational center as:

$$\sigma_k = \frac{\sum_{l=1}^{L} D_k}{\sum_{l=1}^{L} E_k}.$$

Note, the proportion $\{\alpha_c\}_{c=1}^{C}$ on the whole dataset is calculated as:

$$\alpha_c = \frac{\eta_c}{\mathrm{N}}, c = 1, \cdots, C.$$

**Deciding the subphenotypes for each patient**

After running T-LCA, we infer the probability of $n$-th patient belonging to the $c$-th subphenotypes as

$$\omega_{n,c} = \frac{\alpha_c \prod_{t=1}^{T_n} \prod_{k=1}^{K} \frac{1}{\sigma_k \sqrt{(2\pi)}} exp\left(-0.5 \left(\frac{\theta_{n,t,k} - \boldsymbol{\beta}_{c,k} \boldsymbol{\tau}_{\boldsymbol{n},\boldsymbol{t}}}{\sigma_k}\right)^2\right)}{\sum_{c=1}^{C} \alpha_c \prod_{t=1}^{T_n} \prod_{k=1}^{K} \frac{1}{\sigma_k \sqrt{(2\pi)}} exp\left(-0.5 \left(\frac{\theta_{n,t,k} - \boldsymbol{\beta}_{c,k} \boldsymbol{\tau}_{\boldsymbol{n},\boldsymbol{t}}}{\sigma_k}\right)^2\right)}$$

For most of the patients, we found that their probabilities of belonging to one specific subphenotype are clear which means one of $\{\omega_{n,c}\}_{c=1}^{C}$ is large enough (>0.9). However, for some patients, their probabilities of belonging to one specific subphenotype are not clear because none of the $\{\omega_{n,c}\}_{c=1}^{C}$ is larger than 0.5. After observing some typical examples from the data (two examples are shown in Figure 23 in Supplement), we found that these patients are characterized by more clinical topics. Here, we mainly identify the subphenotypes with patients who have certain belongings.

**The reason of fewer key topics was identified on the Mount Sinai dataset**

When validating our method on the Mount Sinai dataset, we found that the optimal number of topics is 20, which is smaller than 30 on other datasets (Supplemental Figure 4). Accordingly, we identified 8 key topics from these 20 topics (Supplemental Figure 5). To further explore this, on three datasets, we compare the percentage of occurrence of top diagnosis records (see Figure 2 and Supplemental Figure 11) in these missing topics, and the results are shown in

Supplemental Table 16. Specifically, for the diagnosis modality of $n$-th patient, the original feature of each visit is a binary vector denoted as $\{\boldsymbol{x}_{n,t}^{(1)}\}_{t=1}^{T_n}$. Assuming that the index of interested diagnosis event is $i$, then we use $\frac{1}{N}\sum_{n=1}^{N}\frac{1}{T_n}\sum_{t=1}^{T_n} x_{n,t,i}^{(1)}$ to represent percentage of occurrence of this event. From the results. we found that these diagnosis events occur less frequently on the Mount Sinai cohort than it does on the other two cohorts. DMTM is developed to capture the common event co-occurrence pattern. This is the reason why fewer key topics was identified on the Mount Sinai dataset compared to the other two datasets used in this work.

### Results

##### Interactions of multimodal clinical events on the Development cohort

The clinical topics describe correlations of multimodal clinical events that belong to one topic. Besides, it would be informative if a method uncover associations between multimodal clinical events even if they belong to different topics, such as understanding the comorbidity interactions, or interaction between disease and medications. For this purpose, we need to evaluate the similarities of two clinical events. A naive method is to use their distributions over patients from raw observational data. However, due to the sparsity and high-dimensionality problems, the measurements are too small to evaluate the similarity accurately as discussed in MixEHR. Instead, as illustrated in Supplemental Figure 7a, we regard the distributions over topics (one row of a topic) as the event feature to calculate their cosine similarities. Based on the similarity, given a specific clinical event as an interesting one, centering on it, DMTM discovered its reasonable interactions with other clinical events. In Figure 7b, we select two typical diseases and one medication in OneFlorida dataset, where i) dementia is a popular disease for the patients who progressed from MCI to AD; and ii) Parkinson's disease (PD) is common in elderly people, to visualize their top (top 10 for diagnosis events while top 5 for drug and procedure events) related events.

##### Subphenotype reproduced results on the Mount Sinai dataset

The size of the Mount Sinai cohort is smaller compared to the other two cohorts, which is one of the reasons why the results of data likelihood and topic coherence (Supplemental Figure 4c) show that 30 topics are not optimal for the Mount Sinai dataset. Therefore, we set the total number of topics on Mount Sinai cohort as 20 and identify 8 key clinical topics (Supplemental Figure 18). We found that these 8 key clinical topics are the subset of 13 topics identified from the other two cohorts, missing topics about Bone, movement, Mental, Digest system, Respiratory system, and Eye. After comparing the number of top diagnosis records (Supplemental Table 16) in missing topics, we observed that the percentage of these diseases in Mount Sinai dataset are much lower than those in the other two datasets (more discussions are provided in Supplemental Methods). Since DMTM focuses on capturing the common code co-occurrent pattern, it may ignore these infrequent patterns. Through our developed T-LCA, due to the lack of these key topics which mainly characterize the subphenotype 4 for 5 in OneFlorida and MarketScan cohort, we identified three subphenotypes on the Mount Sinai dataset, which correspond to the first three subphenotypes in the other two cohorts. These results have validated the reproducibility of the identified subphenotypes.

### Figures and Tables for Supplement

#### Illustration of cohort selection

**Cohort selection on OneFlorida dataset**

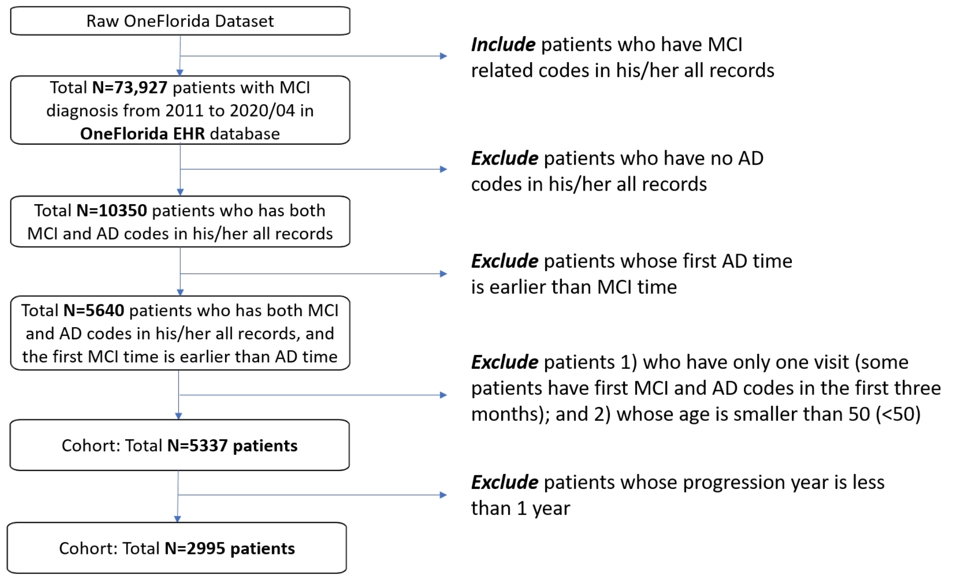

**Cohort selection on MarketScan dataset**

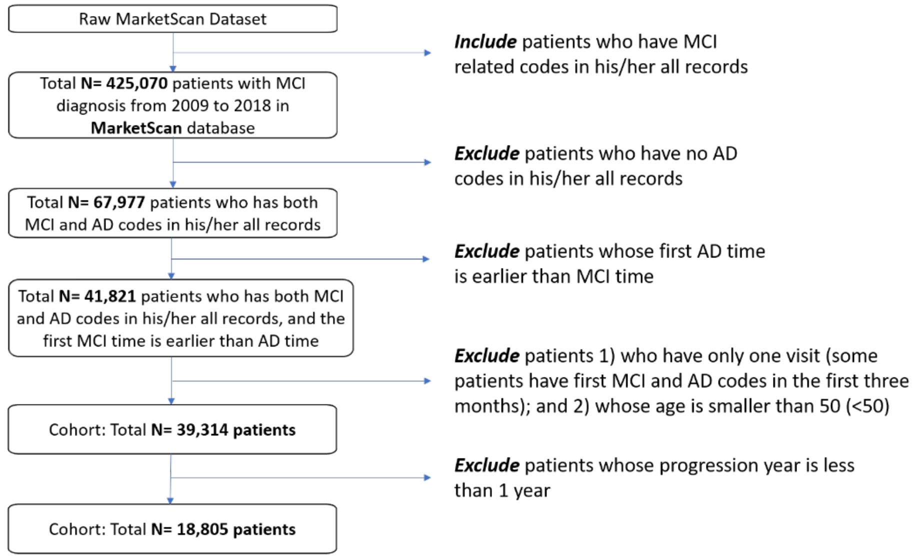

#### Cohort selection on Mount Sinai dataset

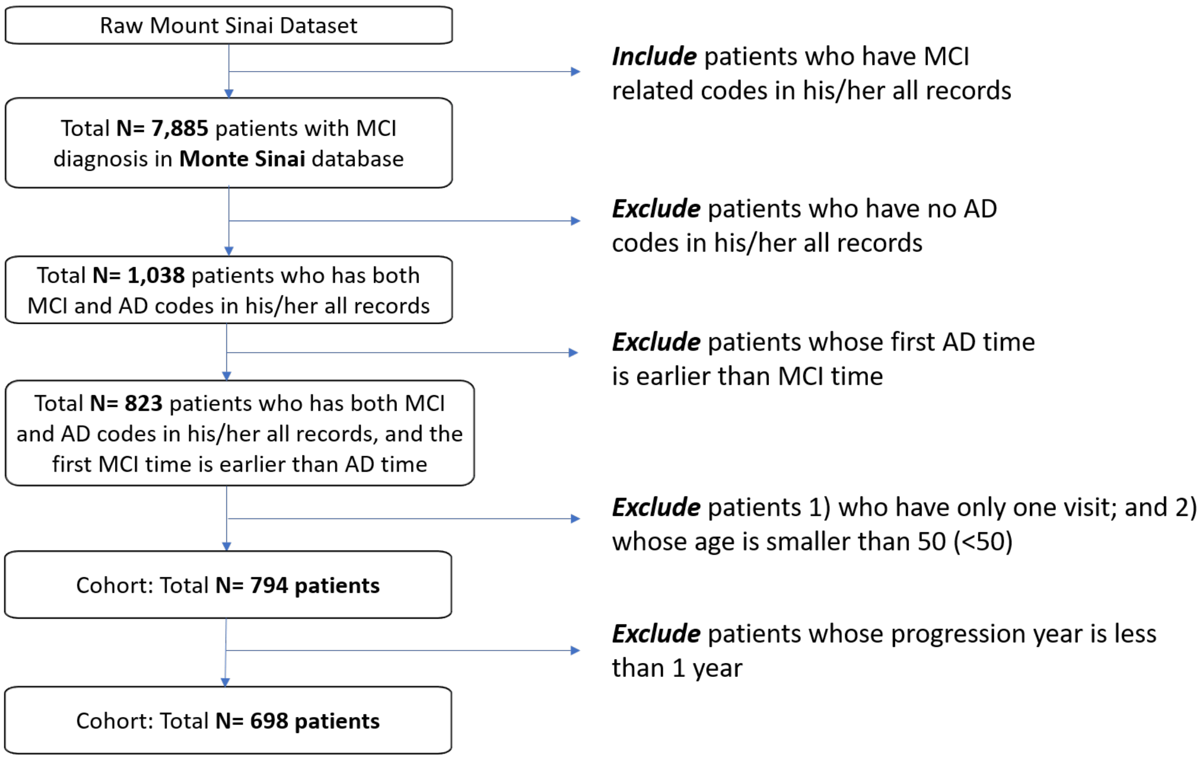

#### Illustration of model selection (number of topics, key topics)

**a. On the OneFlorida dataset (development cohort)**

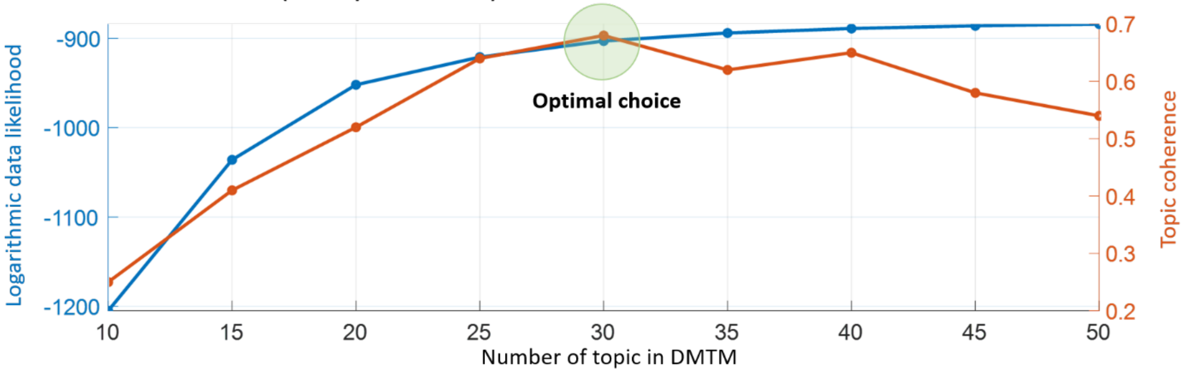

**b. On the MarketScan dataset (validation cohort)**

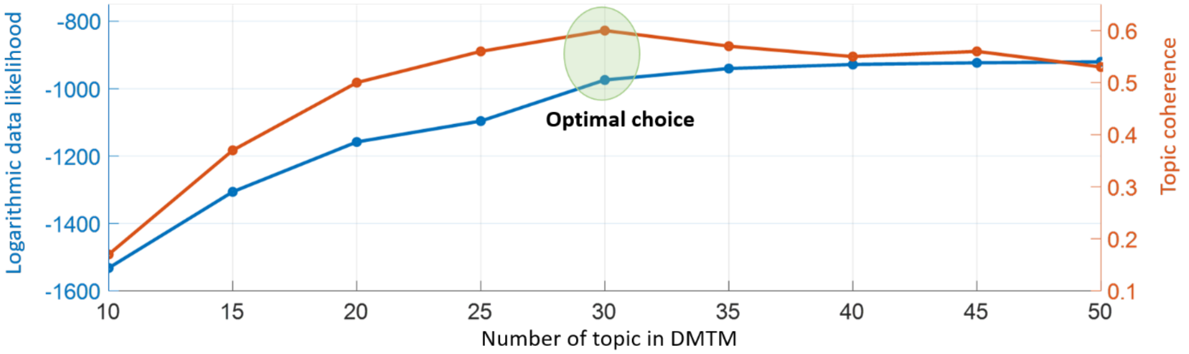

**c. On the Mount Sinai dataset (validation cohort)**

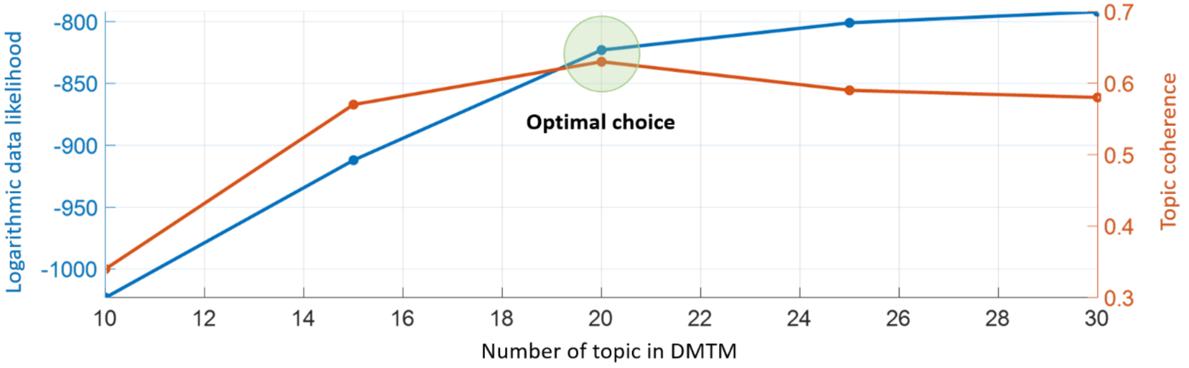

**a. On the OneFlorida dataset (development cohort)**

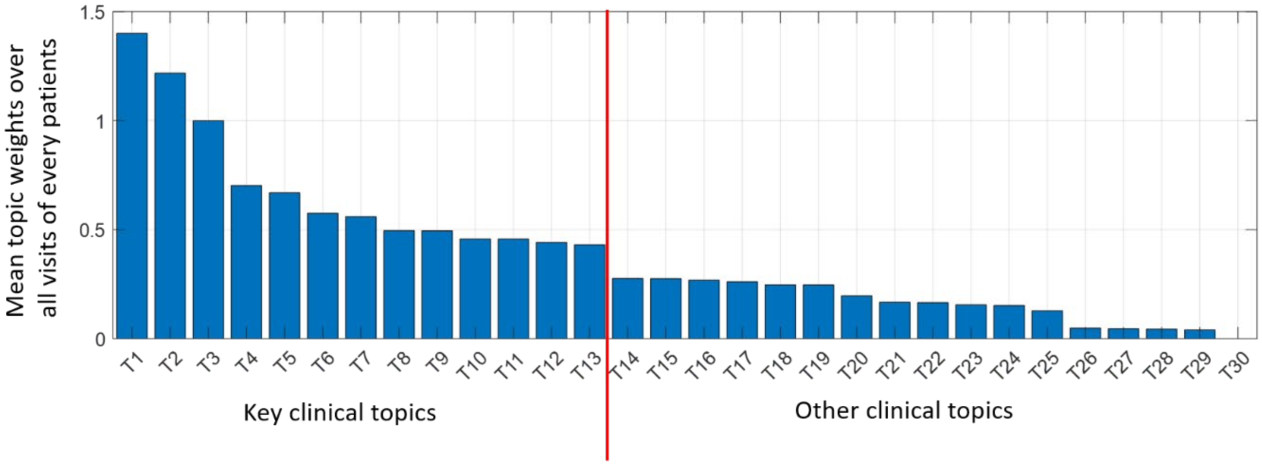

**b. On the MarketScan dataset (validation cohort)**

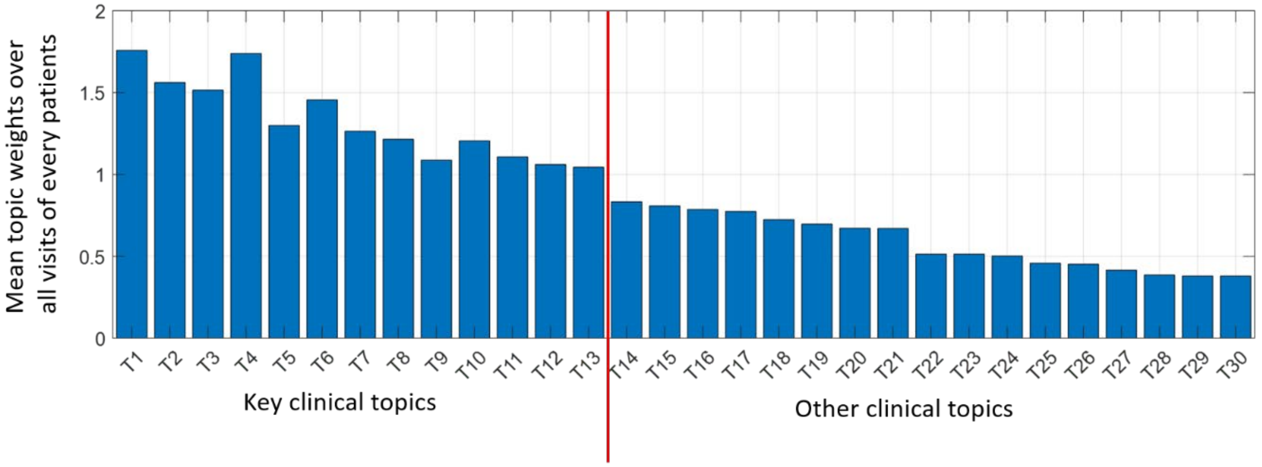

**c. On the Mount Sinai dataset (validation cohort)**

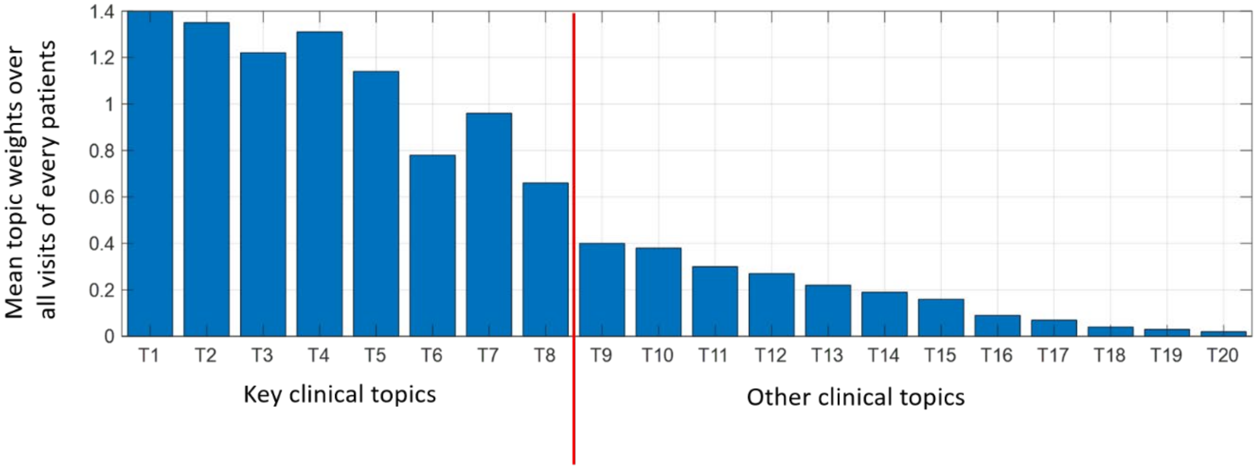

#### Results on the development dataset (OneFlorida dataset).

| Clinical topics | Diagnosis (Phecode) | Drug (RxCUI) | Procedure (CPT) |
|---|---|---|---|
| T14 Nail | Dermatophytosis of nail | Itraconazole | Debridement of nail(s) |
| | Ingrowing nail | Trihexyphenidyl | Subsequent nursing facility care |
| | Diseases of nail | Terbinafine | Paring or cutting of benign hyperkeratotic lesion |
| | Corns and callosities | Ergocalciferol | Trimming of nondystrophic nails |
| | Acquired deformities of finger | Fluoxetine | Avulsion of nail plate |
| T15 Parkinson's disease | Parkinson's disease | Trihexyphenidyl | Exercises to develop range of motion |
| | Muscle weakness | Carbidopa | Activities to improve functional performance |
| | Delirium dementia | Levodopa | Neuromuscular reeducation of movement |
| | Abnormal movement | Amantadine | Self-care/home management training |
| | Dysphagia | Entacapone | Gait training |
| T16 Anemias | Other anemias | Sevelamer | Blood count |
| | Iron deficiency anemias | Leucovorin | Iron binding capacity |
| | Chronic renal failure [CKD] | Folic acid | Collection of venous blood by venipuncture |
| | Anemia in neoplastic disease | Acetaminophen | Blood typing |
| | Renal failure | Ferrous sulfate | Iron |
| T17 Headache | Migraine | Ibuprofen | Outpatient visit for the evaluation |
| | Sleep disorders | Acetaminophen | Subsequent nursing facility care |
| | Other headache syndromes | Naproxen | Glucose; quantitative, blood |
| | Malaise and fatigue | Bupropion | Chloride; blood |
| | Syncope and collapse | Citalopram | Creatinine; blood |
| T18 Back pain | Back pain | Gabapentin | Radiologic examination |
| | Spondylosis and allied disorder | Tizanidine | Neuromuscular reeducation |
| | Spinal stenosis | Duloxetine | Face-to-face talk with the patient and/or family |
| | Cervicalgia | Ibuprofen | Computed tomography |
| | Sleep disorders | Baclofen | Outpatient visit for the evaluation |
| T19 Disease of prostate | Hyperplasia of prostate | Tamsulosin | Measurement of post-voiding residual urine |
| | Cancer of prostate | Finasteride | Prostate specific antigen |
| | Retention of urine | Tamsulosin | Urinalysis |
| | Hematuria | Omeprazole | Outpatient visit for the evaluation |
| | Urinary tract infection | Alfuzosin | Therapeutic procedure |
| T20 Skin | Actinic keratosis | Fluorouracil | Level IV - Surgical pathology |
| | Seborrheic keratosis | Imiquimod | Destruction |
| | Neoplasm of uncertain behavior of skin | Diclofenac | Collection of venous blood |
| | Other non-epithelial cancer of skin | Ingenol | Outpatient visit for the evaluation |
| | Chronic dermatitis due to solar radiation | Aminolevulinic acid | Comprehensive metabolic panel |
| T21 Abdomen | Abdominal pain | Ondansetron | Computed tomography, abdomen and pelvis |
| | Nausea and vomiting | Dicyclomine | Emergency department visit |
| | Other symptoms involving abdomen and pelvis | Tramadol | Lipase |
| | Diverticulosis | Fluticasone | Therapeutic, prophylactic, or diagnostic injection |
| | Abdominal hernia | Promethazine | Comprehensive metabolic panel |
| T22 Disease of liver | Cirrhosis of liver without mention of alcohol | Propylthiouracil | Blood count |
| | Other disorders of liver damage | Oxandrolone | Comprehensive metabolic panel |
| | Malignant neoplasm of liver | Diazepam | Unlisted procedure, liver |
| | Other anemias | Lactulose | Liver imaging; with vascular flow |
| | Thrombocytopenia | Acetaminophen | Management of liver hemorrhage |
| T23 Disease of lung | Abnormal findings examination of lungs | Ciprofloxacin | Biopsy/wedge resection lung |
| | Nonspecific chest pain | Sulfamethoxazole | Destruction Procedures on the lung |
| | Bacterial pneumonia | Avelumab | Resection of apical lung tumor |
| | Bacterial infection | Cephalexin | Manipulation chest wall |
| | Pneumonia | Acetaminophen | Computed tomography |
| T24 Mixed | Hypertension | Aspirin | Prothrombin time |
| | Cardiac dysrhythmias | Metoprolol | Blood count |
| | Ischemic Heart Disease | Amlodipine | Electrocardiogram |
| | Essential hypertension | Tramadol | Outpatient visit for the evaluation |
| | Disorders of lipoid metabolism | Furosemide | Comprehensive metabolic panel |
| T25 Mixed | Essential hypertension | Donepezil | Medication list documented in medical record |
| | Cerebral degeneration | Memantine | Eecent diastolic blood pressure |
| | Mixed hyperlipidemia | Aspirin | Review of all medications |
| | Memory loss | Atorvastatin | Recent systolic blood pressure |
| | Dementias | Diclofenac | Body Mass Index (BMI) |
| T26 Mixed | Acute renal failure | Sulfamethoxazole | Hospital care, per day |
| | Urinary tract infection | Ceftriaxone | Electrocardiogram |
| | Type 2 diabetes | Atorvastatin | Radiologic examination |
| | Chronic renal failure [CKD] | Insulin | Urinalysis |
| | Essential hypertension | Pantoprazole | Glucose, blood |
| T27 Mixed | Disorders of muscle, ligament, and fascia | Folic acid | Therapeutic procedure |
| | Difficulty in walking | Tizanidine | Therapeutic activities |
| | Back pain | Gadobenate | Self-care/home management training |
| | Hemarthrosis | Ibuprofen | Physical therapy evaluation |
| | Decubitus | Deflazacort | Biopsy of muscle |
| T28 Mixed | Cerebral degeneration | Donepezil | Magnetic resonance |
| | Dementias | Memantine | Outpatient visit for the evaluation |
| | Memory loss | Alprazolam | Ultrasound |
| | Neurological disorders | Gabapentin | Dementia severity classified |
| | Alzheimer's disease | Vitamin B12 | Care for patient w. cognitive problem |
| T29 Mixed | Glaucoma | Temazepam | Personal care services |
| | Mood disorders | Citalopram | Episode of major depressive disorder |
| | Major depressive disorder | Metipranolol | Various eye detections |
| | Other retinal disorders | Methazolamide | Companion care |
| | Hearing loss | Aspirin | Hearing aid examination |
| T30 Mixed | Malignant neoplasm | Sodium chloride | Basic metabolic panel |
| | Essential hypertension | Acetaminophen | Blood count |
| | Other chronic ischemic heart disease | Heparin | Magnesium |
| | Other diseases of respiratory system | Iohexol | Troponin, quantitative |
| | Hyperlipidemia | Fentanyl | Phosphorus inorganic (phosphate) |

Color bar: 0 0.1 0.2 0.3 0.4 0.5 0.6

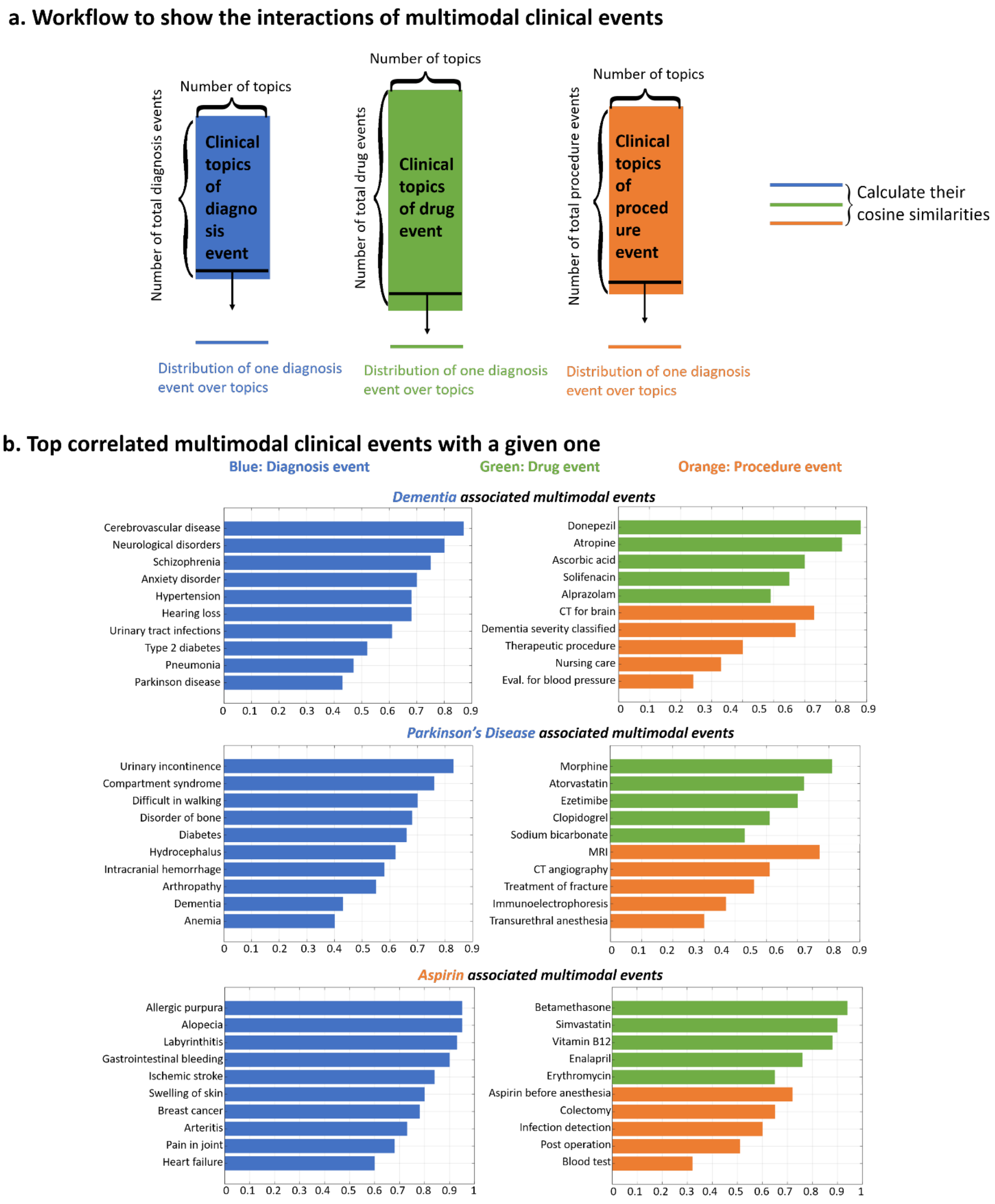
a. Workflow to show the interactions of multimodal clinical events
Number of topics
Number of total diagnosis events
Clinical topics of diagnosis event
Number of total drug events
Clinical topics of drug event
Number of total procedure events
Clinical topics of procedure event
Calculate their cosine similarities
Distribution of one diagnosis event over topics
Distribution of one diagnosis event over topics
Distribution of one diagnosis event over topics
b. Top correlated multimodal clinical events with a given one
Blue: Diagnosis event
Green: Drug event
Orange: Procedure event
Dementia associated multimodal events
Cerebrovascular disease
Neurological disorders
Schizophrenia
Anxiety disorder
Hypertension
Hearing loss
Urinary tract infections
Type 2 diabetes
Pneumonia
Parkinson disease
Donepezil
Atropine
Ascorbic acid
Solifenacin
Alprazolam
CT for brain
Dementia severity classified
Therapeutic procedure
Nursing care
Eval. for blood pressure
Parkinson's Disease associated multimodal events
Urinary incontinence
Compartment syndrome
Difficult in walking
Disorder of bone
Diabetes
Hydrocephalus
Intracranial hemorrhage
Arthropathy
Dementia
Anemia
Morphine
Atorvastatin
Ezetimibe
Clopidogrel
Sodium bicarbonate
MRI
CT angiography
Treatment of fracture
Immunoelectrophoresis
Transurethral anesthesia
Aspirin associated multimodal events
Allergic purpura
Alopecia
Labyrinthitis
Gastrointestinal bleeding
Ischemic stroke
Swelling of skin
Breast cancer
Arteritis
Pain in joint
Heart failure
Betamethasone
Simvastatin
Vitamin B12
Enalapril
Erythromycin
Aspirin before anesthesia
Colectomy
Infection detection
Post operation
Blood test
0 0.1 0.2 0.3 0.4 0.5 0.6 0.7 0.8 0.9 1

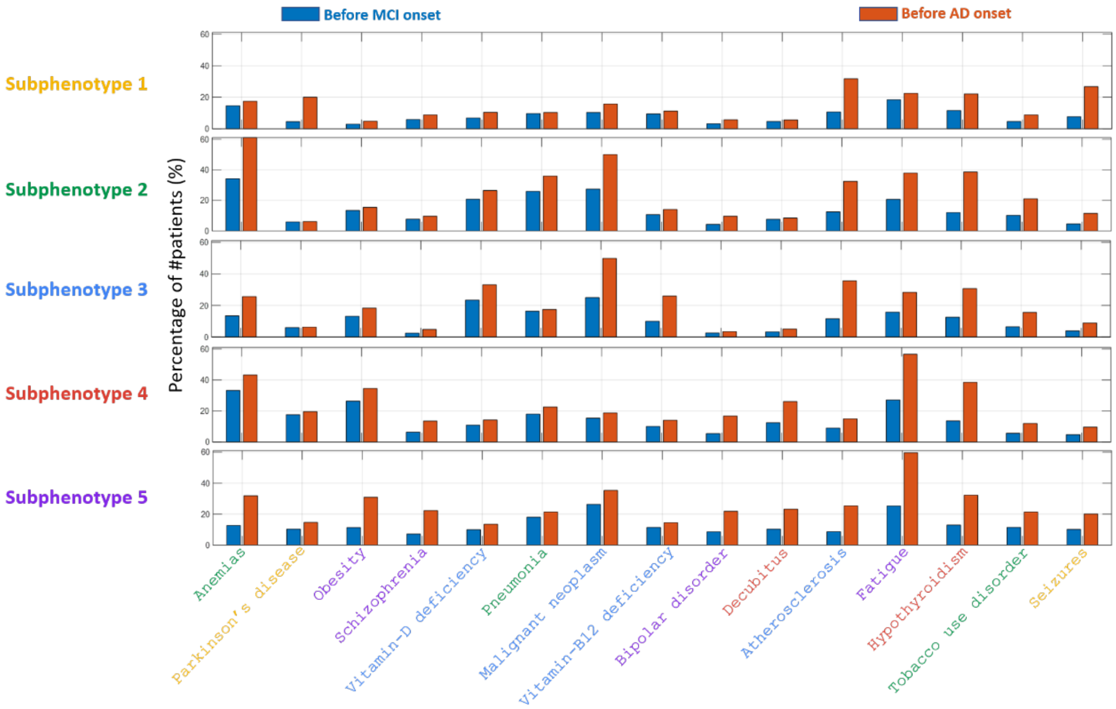

Before MCI onset
Before AD onset
Subphenotype 1
Subphenotype 2
Subphenotype 3
Subphenotype 4
Subphenotype 5
Percentage of #patients (%)
Anemias
Parkinson's disease
Obesity
Schizophrenia
Vitamin-D deficiency
Pneumonia
Malignant neoplasm
Vitamin-B12 deficiency
Bipolar disorder
Decubitus
Atherosclerosis
Fatigue
Hypothyroidism
Tobacco use disorder
Seizures

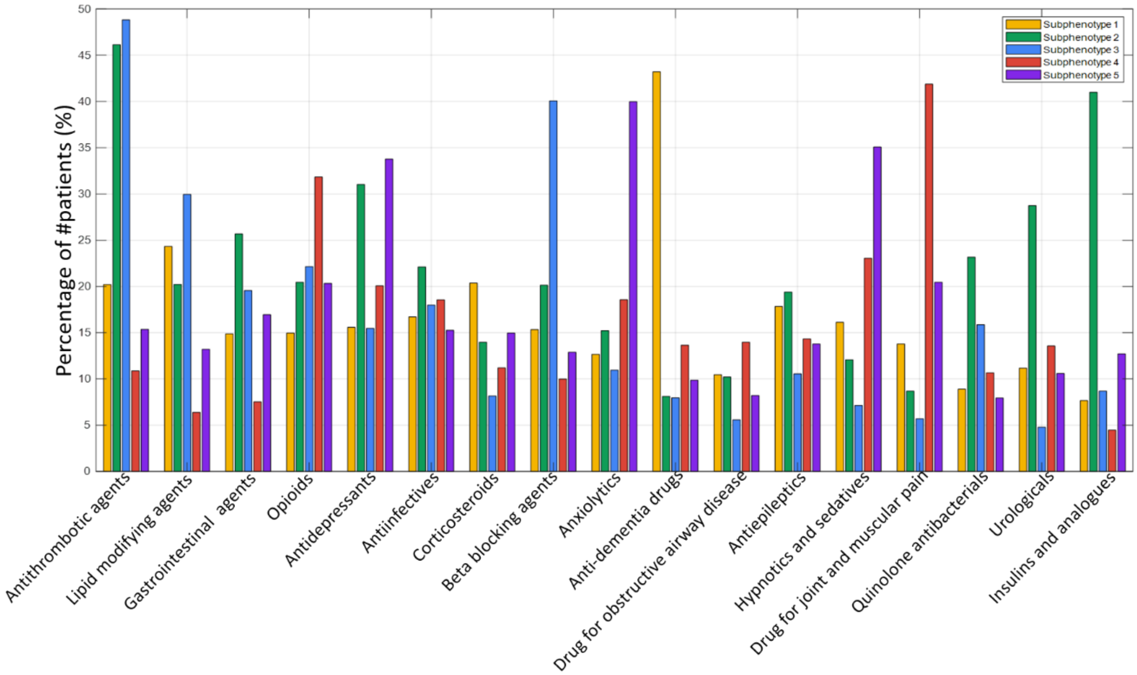

Percentage of #patients (%)
0
5
10
15
20
25
30
35
40
45
50
Subphenotype 1
Subphenotype 2
Subphenotype 3
Subphenotype 4
Subphenotype 5
Antithrombotic agents
Lipid modifying agents
Gastrointestinal agents
Opioids
Antidepressants
Antiinfectives
Corticosteroids
Beta blocking agents
Anxiolytics
Anti-dementia drugs
Drug for obstructive airway disease
Antiepileptics
Hypnotics and sedatives
Drug for joint and muscular pain
Quinolone antibacterials
Urologicals
Insulins and analogues

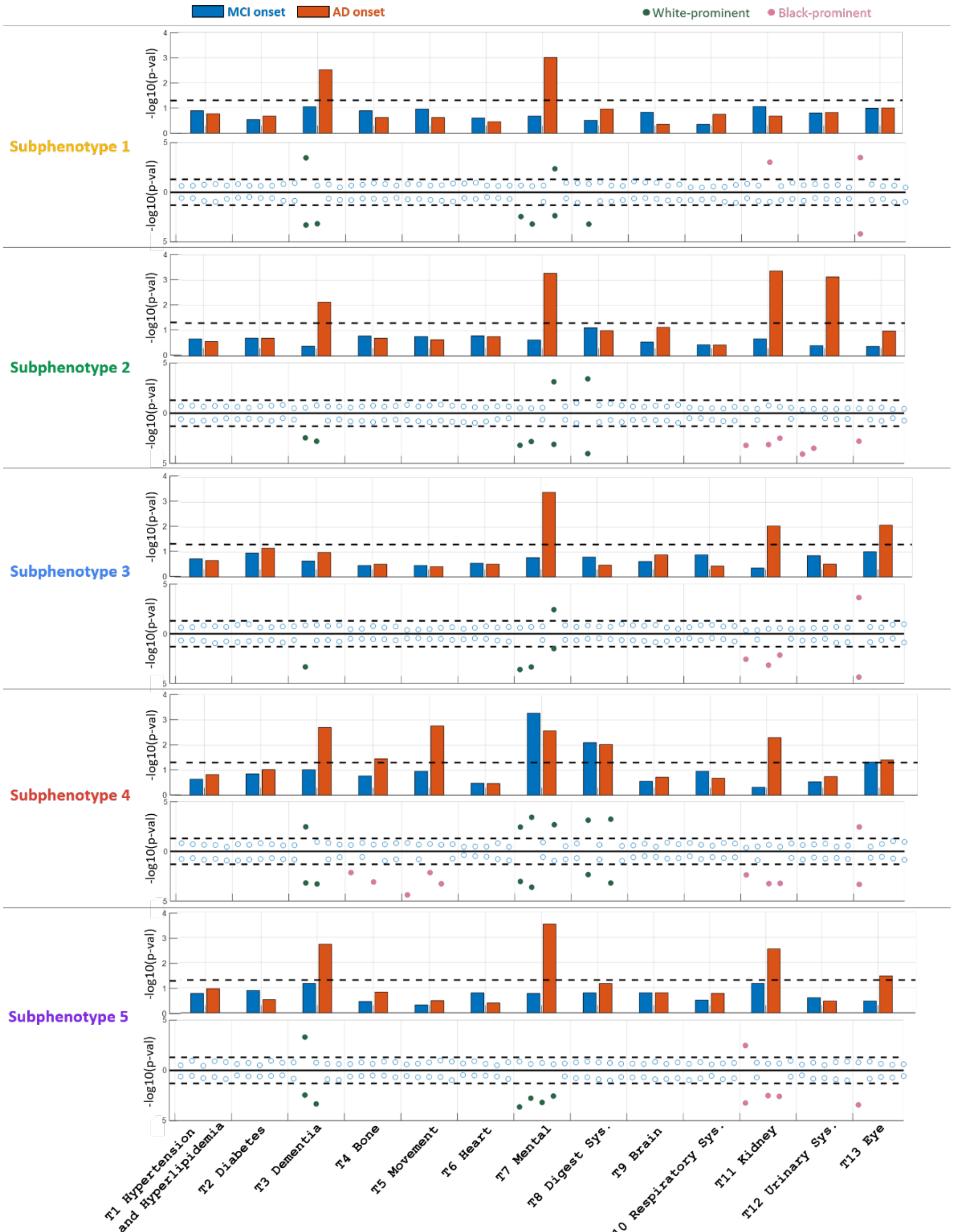

**T3:** Neurological disorders (2nd), Memory loss (3rd)
**T7:** Generalized anxiety disorder (1st), Major depressive disorder (2nd)
**T11:** Chronic kidney failure (1st), Acute renal failure (3rd)
**T13:** Glaucoma (1st)

#### Results on the MarketScan dataset (validation cohort).

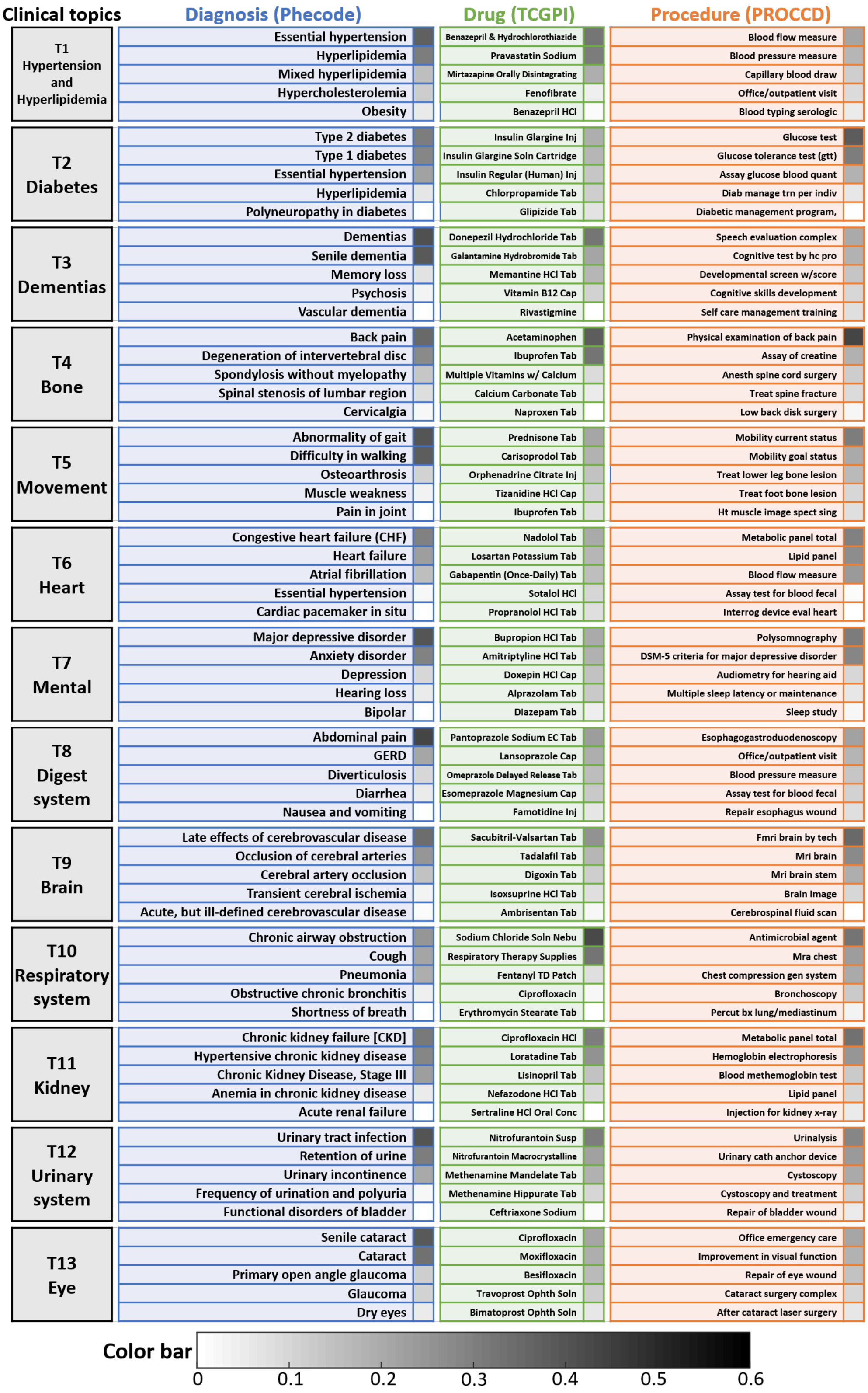

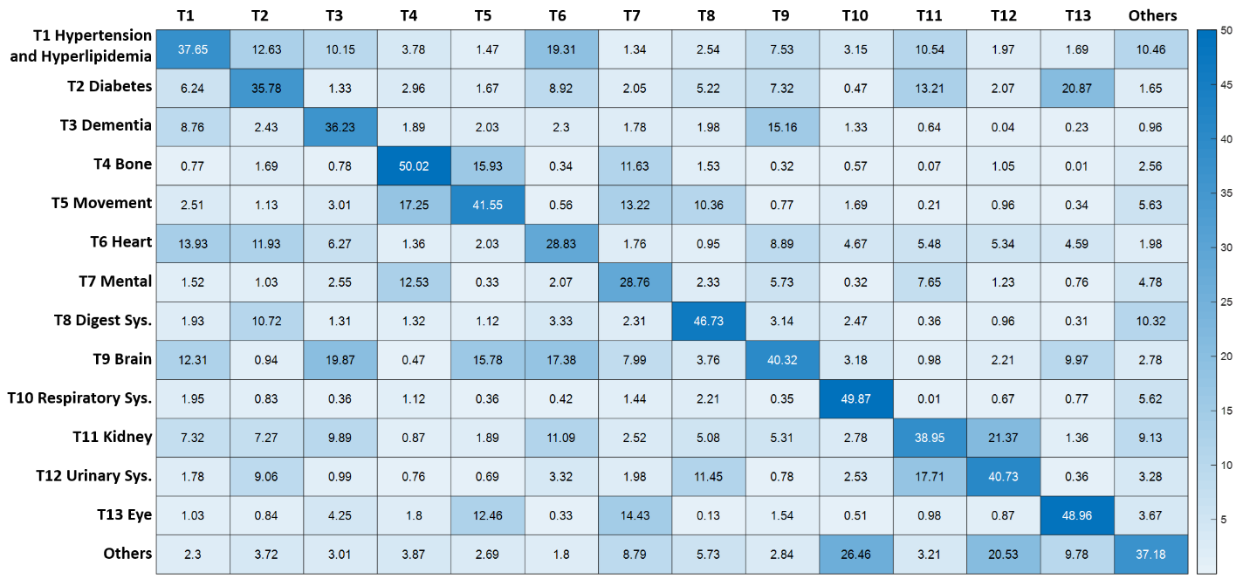
T1 T2 T3 T4 T5 T6 T7 T8 T9 T10 T11 T12 T13 Others
T1 Hypertension and Hyperlipidemia 37.65 12.63 10.15 3.78 1.47 19.31 1.34 2.54 7.53 3.15 10.54 1.97 1.69 10.46
T2 Diabetes 6.24 35.78 1.33 2.96 1.67 8.92 2.05 5.22 7.32 0.47 13.21 2.07 20.87 1.65
T3 Dementia 8.76 2.43 36.23 1.89 2.03 2.3 1.78 1.98 15.16 1.33 0.64 0.04 0.23 0.96
T4 Bone 0.77 1.69 0.78 50.02 15.93 0.34 11.63 1.53 0.32 0.57 0.07 1.05 0.01 2.56
T5 Movement 2.51 1.13 3.01 17.25 41.55 0.56 13.22 10.36 0.77 1.69 0.21 0.96 0.34 5.63
T6 Heart 13.93 11.93 6.27 1.36 2.03 28.83 1.76 0.95 8.89 4.67 5.48 5.34 4.59 1.98
T7 Mental 1.52 1.03 2.55 12.53 0.33 2.07 28.76 2.33 5.73 0.32 7.65 1.23 0.76 4.78
T8 Digest Sys. 1.93 10.72 1.31 1.32 1.12 3.33 2.31 46.73 3.14 2.47 0.36 0.96 0.31 10.32
T9 Brain 12.31 0.94 19.87 0.47 15.78 17.38 7.99 3.76 40.32 3.18 0.98 2.21 9.97 2.78
T10 Respiratory Sys. 1.95 0.83 0.36 1.12 0.36 0.42 1.44 2.21 0.35 49.87 0.01 0.67 0.77 5.62
T11 Kidney 7.32 7.27 9.89 0.87 1.89 11.09 2.52 5.08 5.31 2.78 38.95 21.37 1.36 9.13
T12 Urinary Sys. 1.78 9.06 0.99 0.76 0.69 3.32 1.98 11.45 0.78 2.53 17.71 40.73 0.36 3.28
T13 Eye 1.03 0.84 4.25 1.8 12.46 0.33 14.43 0.13 1.54 0.51 0.98 0.87 48.96 3.67
Others 2.3 3.72 3.01 3.87 2.69 1.8 8.79 5.73 2.84 26.46 3.21 20.53 9.78 37.18
50
45
40
35
30
25
20
15
10
5

**a. Change of topic compositions with time according to different subphenotypes**

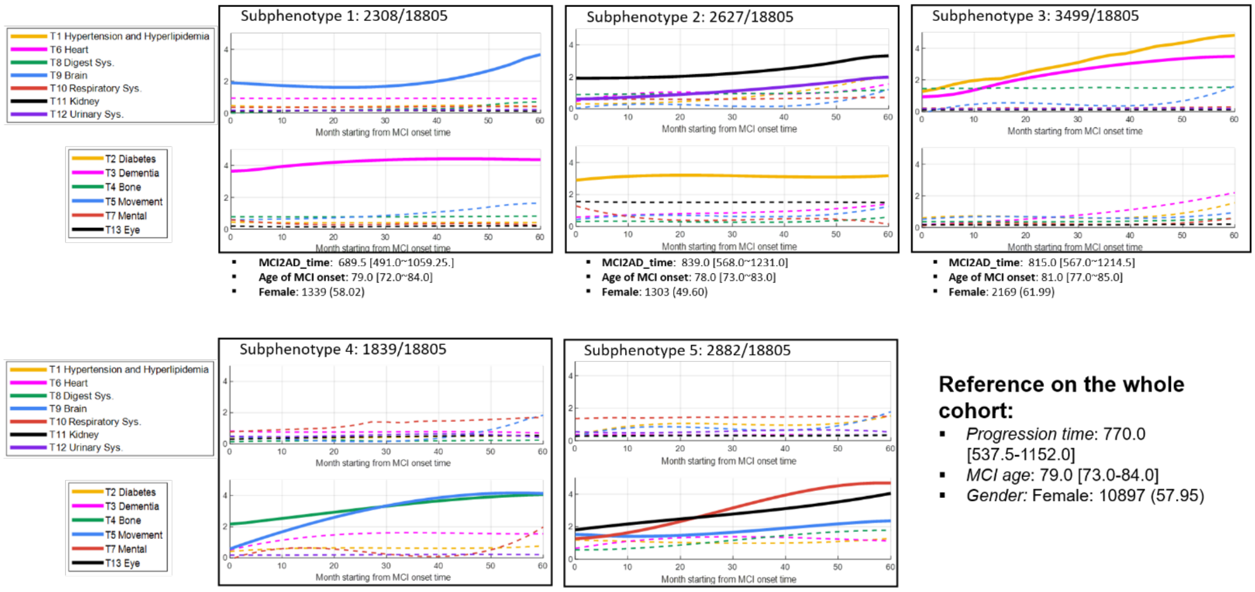

Subphenotype 1:
- **MCI2AD_time**: 689.5 [491.0~1059.25.]
- **Age of MCI onset**: 79.0 [72.0~84.0]
- **Female**: 1339 (58.02)

Subphenotype 2:
- **MCI2AD_time**: 839.0 [568.0~1231.0]
- **Age of MCI onset**: 78.0 [73.0~83.0]
- **Female**: 1303 (49.60)

Subphenotype 3:
- **MCI2AD_time**: 815.0 [567.0~1214.5]
- **Age of MCI onset**: 81.0 [77.0~85.0]
- **Female**: 2169 (61.99)

Subphenotype 4:
- **MCI2AD_time**: 745.0 [530.0~1108.5]
- **Age of MCI onset**: 83.0 [78.0~87.0]
- **Female**: 1394 (75.80)

Subphenotype 5:
- **MCI2AD_time**: 897.5 [688.0~1274.75]
- **Age of MCI onset**: 74.0 [66.0~80.0]
- **Female**: 1895 (65.76)

**Reference on the whole cohort:**
- *Progression time*: 770.0 [537.5-1152.0]
- *MCI age*: 79.0 [73.0-84.0]
- *Gender:* Female: 10897 (57.95)

**b. Change of each topic with time within different subphenotypes**

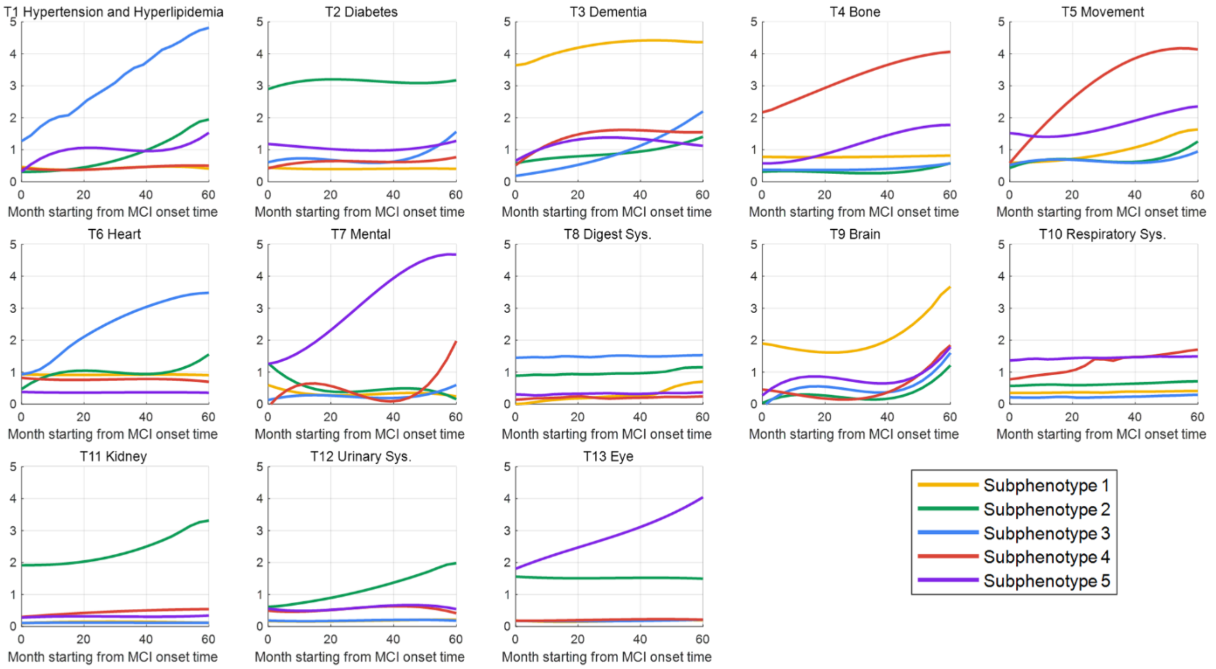

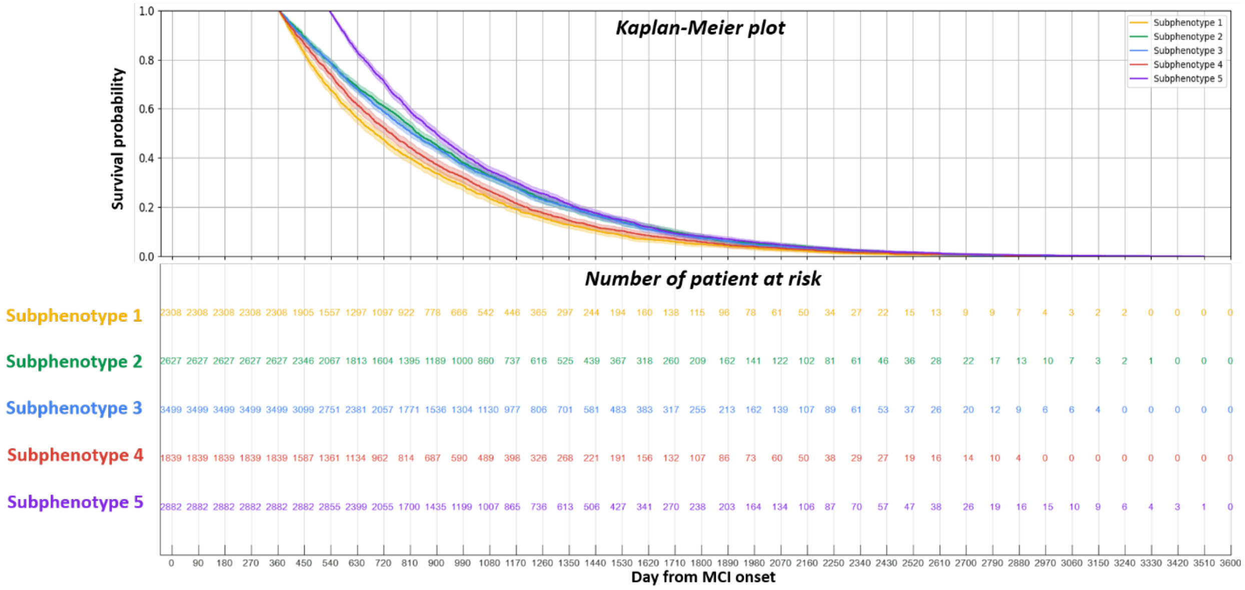

Kaplan-Meier plot
Survival probability
Subphenotype 1
Subphenotype 2
Subphenotype 3
Subphenotype 4
Subphenotype 5
Number of patient at risk
Subphenotype 1 2308 2308 2308 2308 2308 1905 1557 1297 1097 922 778 666 542 446 365 297 244 194 160 138 115 96 78 61 50 34 27 22 15 13 9 9 7 4 3 2 2 0 0 0 0
Subphenotype 2 2627 2627 2627 2627 2627 2346 2067 1813 1604 1395 1189 1000 860 737 616 525 439 367 318 260 209 162 141 122 102 81 61 46 36 28 22 17 13 10 7 3 2 1 0 0 0
Subphenotype 3 3499 3499 3499 3499 3499 3099 2751 2381 2057 1771 1536 1304 1130 977 806 701 581 483 383 317 255 213 162 139 107 89 61 53 37 26 20 12 9 6 6 4 0 0 0 0 0
Subphenotype 4 1839 1839 1839 1839 1839 1587 1361 1134 962 814 687 590 489 398 326 268 221 191 156 132 107 86 73 60 50 38 29 27 19 16 14 10 4 0 0 0 0 0 0 0 0
Subphenotype 5 2882 2882 2882 2882 2882 2882 2855 2399 2055 1700 1435 1199 1007 865 736 613 506 427 341 270 238 203 164 134 106 87 70 57 47 38 26 19 16 15 10 9 6 4 3 1 0
0 90 180 270 360 450 540 630 720 810 900 990 1080 1170 1260 1350 1440 1530 1620 1710 1800 1890 1980 2070 2160 2250 2340 2430 2520 2610 2700 2790 2880 2970 3060 3150 3240 3330 3420 3510 3600
Day from MCI onset

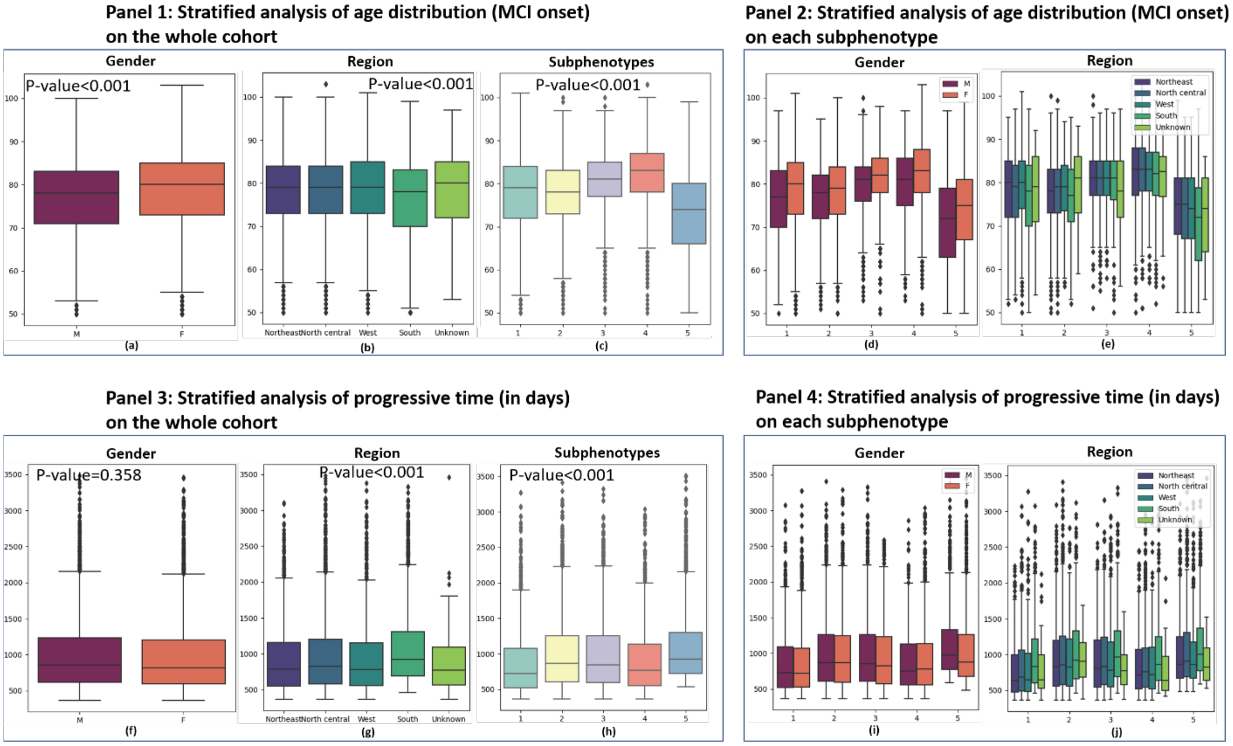

Panel 1: Stratified analysis of age distribution (MCI onset) on the whole cohort
Gender
P-value<0.001
M
F
(a)
Region
P-value<0.001
Northeast North central West South Unknown
(b)
Subphenotypes
P-value<0.001
(c)
Panel 2: Stratified analysis of age distribution (MCI onset) on each subphenotype
Gender
(d)
Region
Northeast
North central
West
South
Unknown
(e)
Panel 3: Stratified analysis of progressive time (in days) on the whole cohort
Gender
P-value=0.358
(f)
Region
P-value<0.001
(g)
Subphenotypes
P-value<0.001
(h)
Panel 4: Stratified analysis of progressive time (in days) on each subphenotype
Gender
(i)
Region
(j)

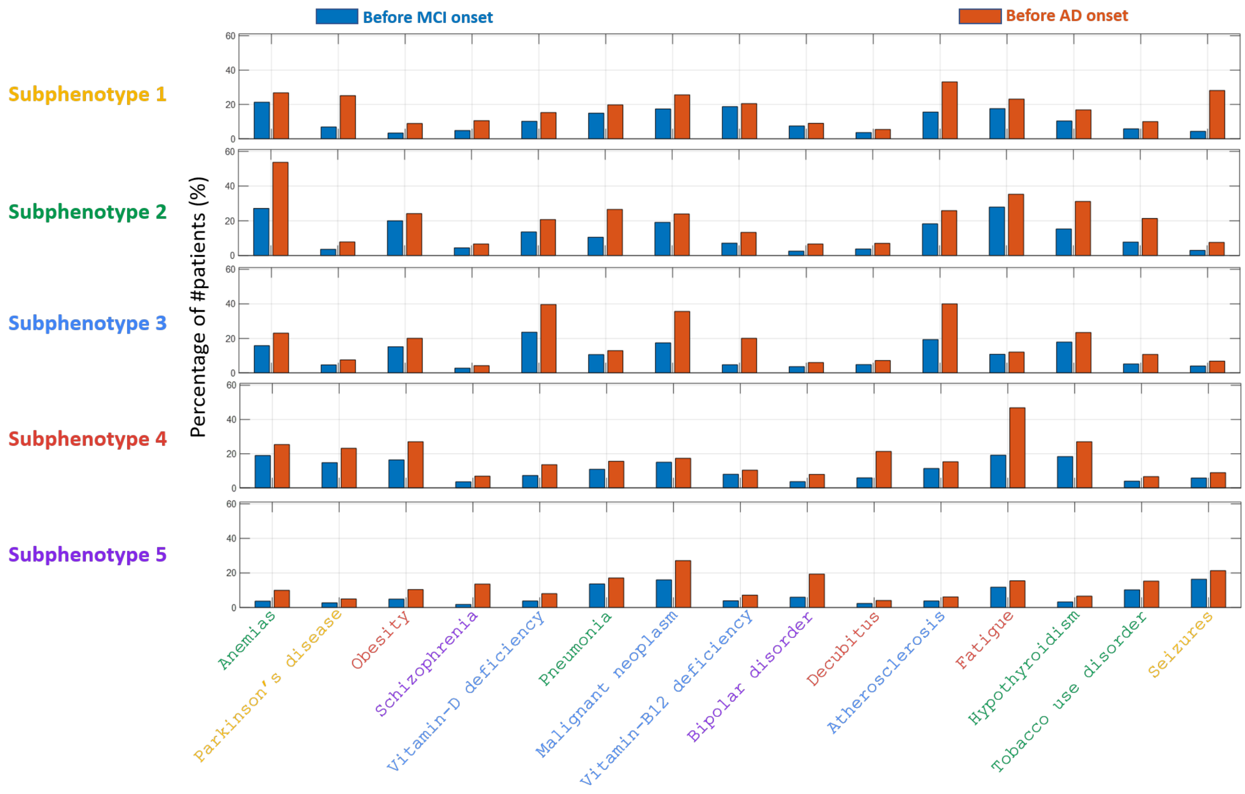

Before MCI onset
Before AD onset
Subphenotype 1
Subphenotype 2
Subphenotype 3
Subphenotype 4
Subphenotype 5
Percentage of #patients (%)
Anemias
Parkinson's disease
Obesity
Schizophrenia
Vitamin-D deficiency
Pneumonia
Malignant neoplasm
Vitamin-B12 deficiency
Bipolar disorder
Decubitus
Atherosclerosis
Fatigue
Hypothyroidism
Tobacco use disorder
Seizures

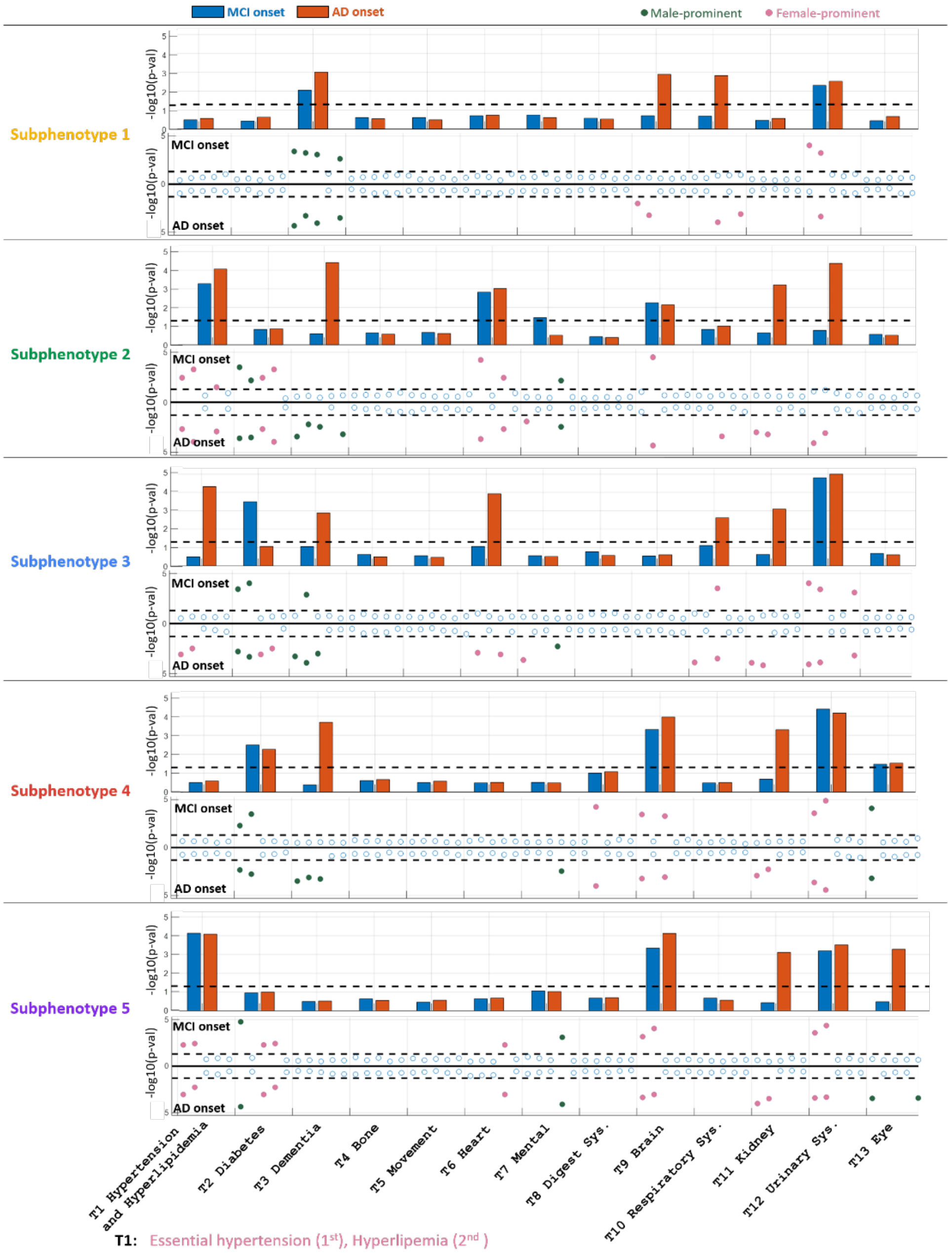

**T1:** Essential hypertension (1st), Hyperlipemia (2nd )
**T2:** Type 2 diabetes (1st), Type 1 diabetes (2nd),
**T3:** Dementia (1st), Senile dementia (2nd), Memory loss (3rd)
**T7:** Hearing loss (4th)
**T9:** Late effects of cerebrovascular disease (1st), Occlusion of cerebral arteries (2nd)
**T11:** Chronic kidney failure (1st), Hypertensive chronic kidney disease (2nd)
**T12:** Urinary tract infection (1st), Retention of urine (2nd)

**Results on the Mount Sinai dataset (validation cohort).**

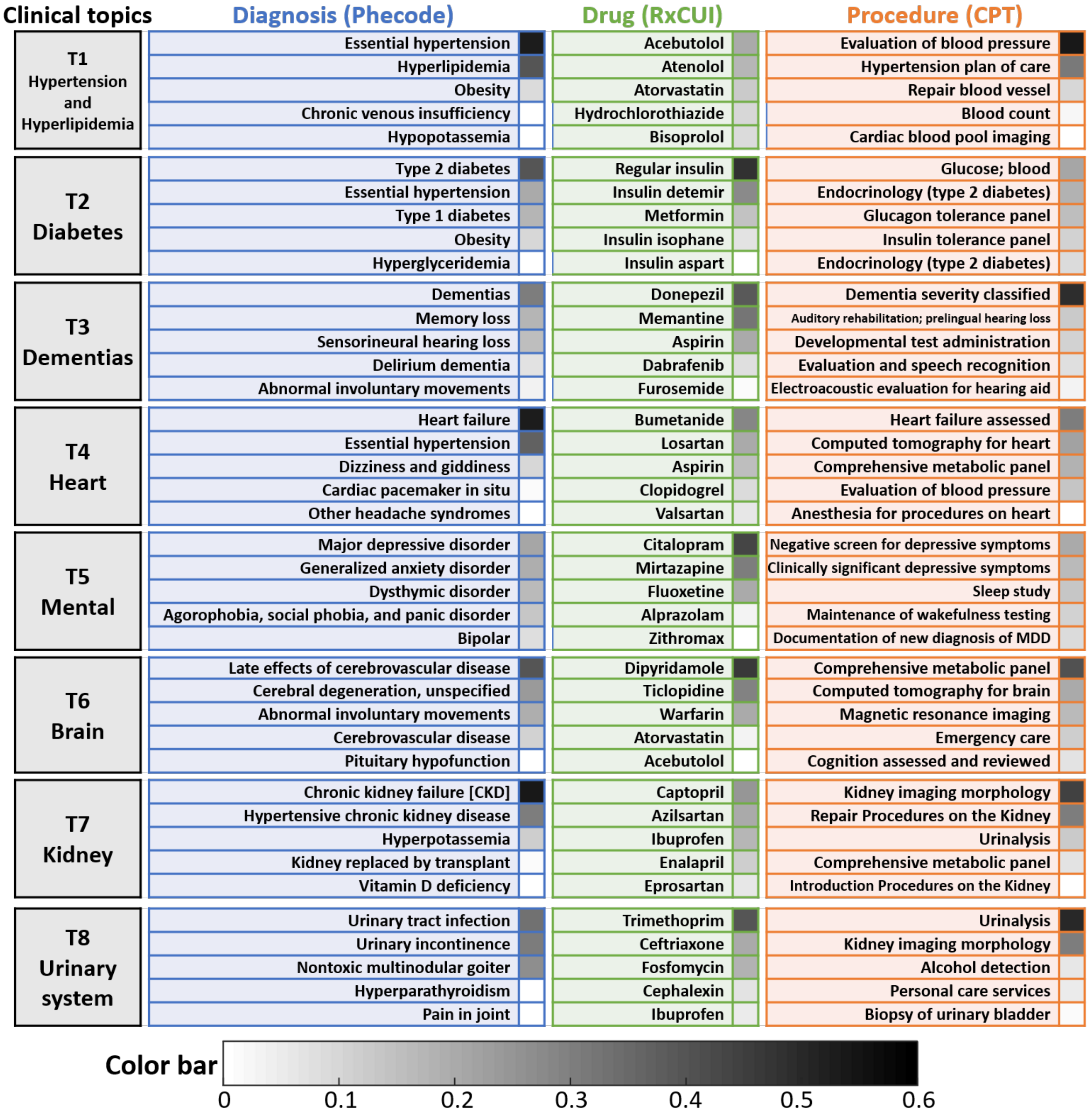

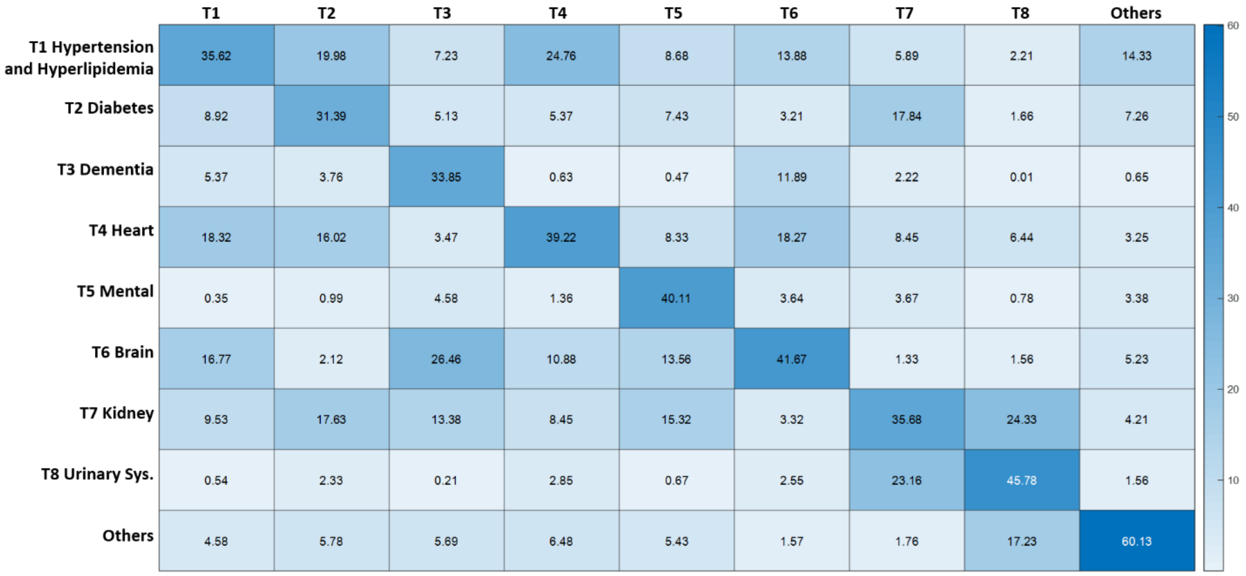
T1 T2 T3 T4 T5 T6 T7 T8 Others
T1 Hypertension and Hyperlipidemia 35.62 19.98 7.23 24.76 8.68 13.88 5.89 2.21 14.33
T2 Diabetes 8.92 31.39 5.13 5.37 7.43 3.21 17.84 1.66 7.26
T3 Dementia 5.37 3.76 33.85 0.63 0.47 11.89 2.22 0.01 0.65
T4 Heart 18.32 16.02 3.47 39.22 8.33 18.27 8.45 6.44 3.25
T5 Mental 0.35 0.99 4.58 1.36 40.11 3.64 3.67 0.78 3.38
T6 Brain 16.77 2.12 26.46 10.88 13.56 41.67 1.33 1.56 5.23
T7 Kidney 9.53 17.63 13.38 8.45 15.32 3.32 35.68 24.33 4.21
T8 Urinary Sys. 0.54 2.33 0.21 2.85 0.67 2.55 23.16 45.78 1.56
Others 4.58 5.78 5.69 6.48 5.43 1.57 1.76 17.23 60.13
60
50
40
30
20
10

**a. Change of topic compositions with time according to different subphenotypes**

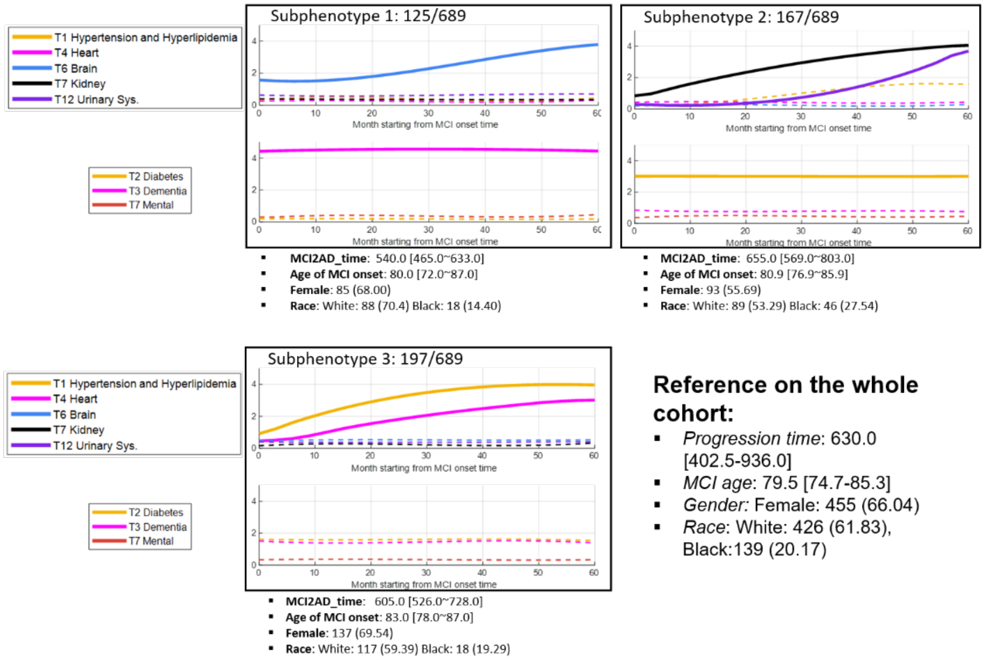

**b. Change of each topic with time within different subphenotypes**

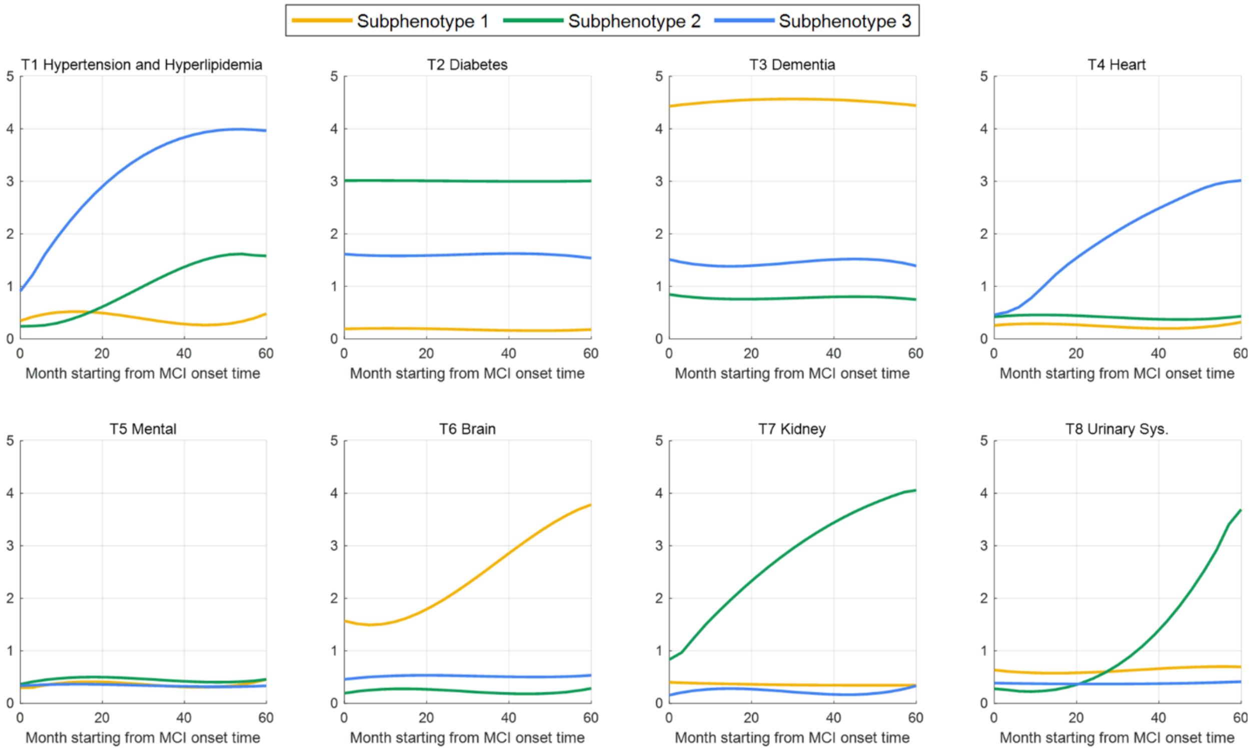

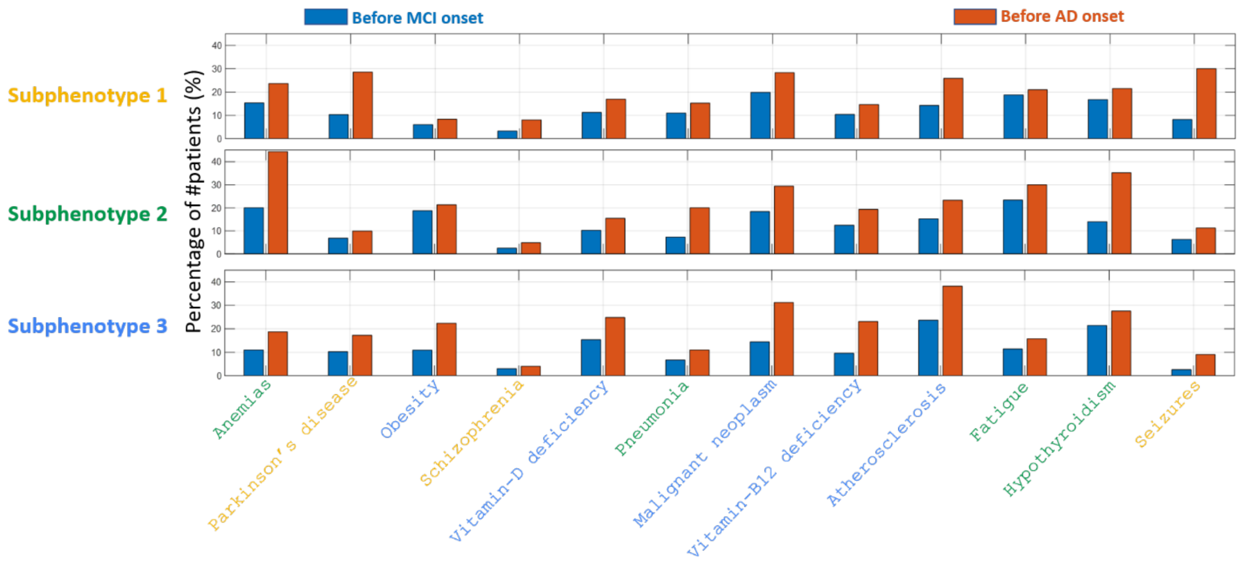

Before MCI onset
Before AD onset
Subphenotype 1
Subphenotype 2
Subphenotype 3
Percentage of #patients (%)
Anemias
Parkinson's disease
Obesity
Schizophrenia
Vitamin-D deficiency
Pneumonia
Malignant neoplasm
Vitamin-B12 deficiency
Atherosclerosis
Fatigue
Hypothyroidism
Seizures

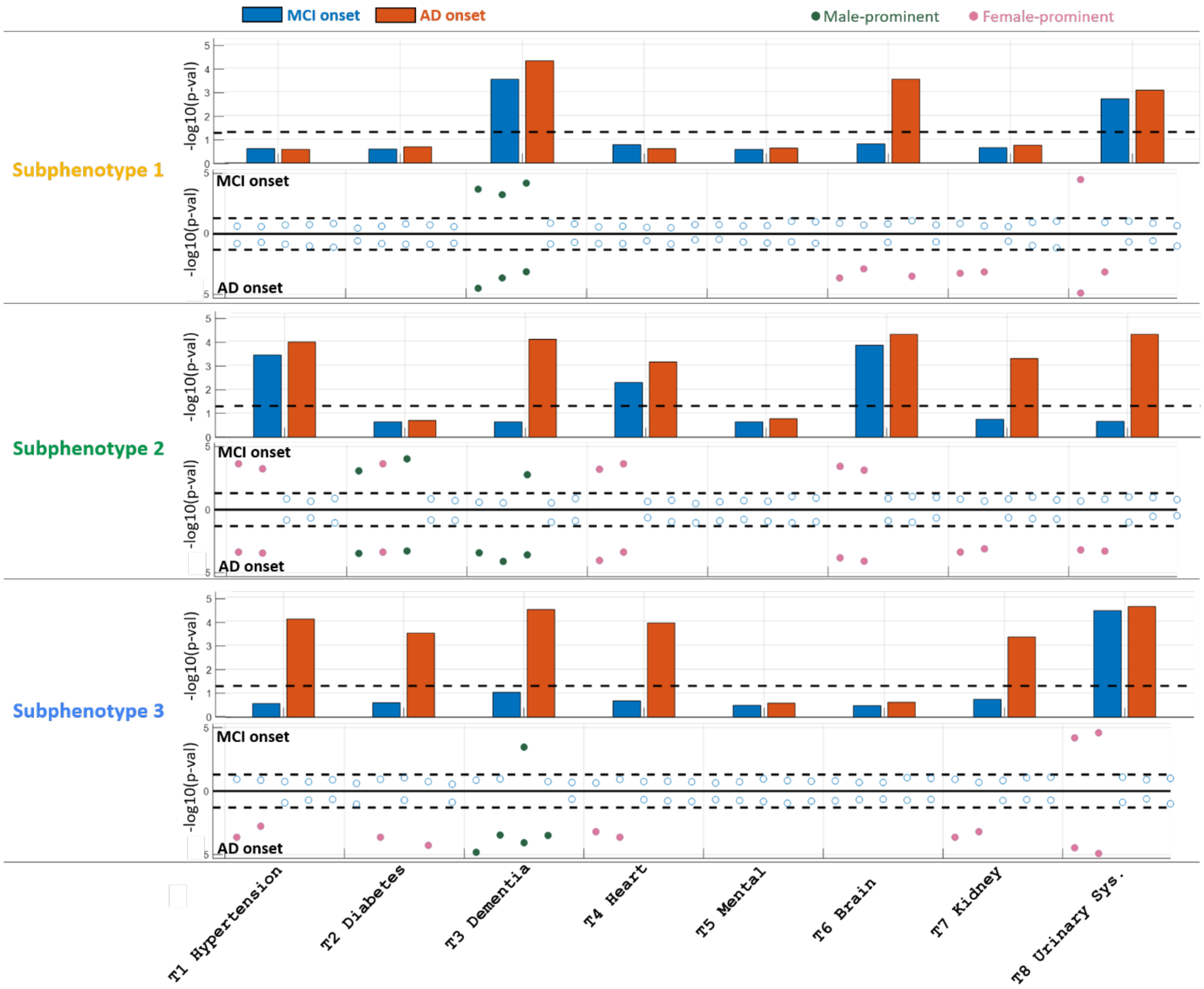

**T1:** Essential hypertension (1st), Hyperlipemia (2nd )
**T3:** Dementia (1st), Memory loss (2nd), Hearing loss (3rd)
**T4:** Heart failure (1st), Essential hypertension (2nd ),
**T6:** Late effects of cerebrovascular disease (1st), Cerebral degeneration, unspecified (2nd)
**T7:** Chronic kidney failure (1st), Hypertensive chronic kidney disease (2nd)
**T8:** Urinary tract infection (1st), Urinary incontinence(2nd)

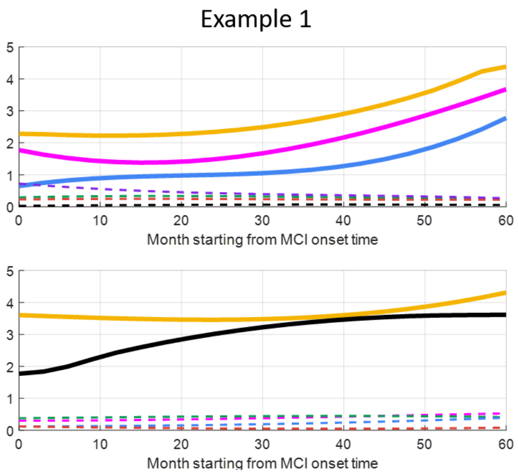

The probability of this sample belonging to each subphenotype:
[S1: 0.288, S2: 0.005, S3: 0.472, S4: 0.004, S5: 0.231]

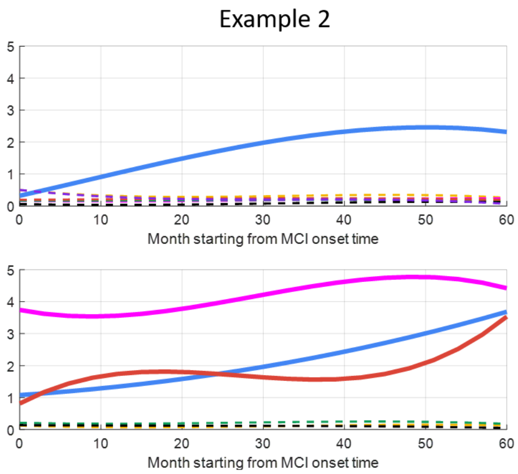

The probability of this sample belonging to each subphenotype:
[S1: 0.428, S2: 0.003, S3: 0.068, S4: 0.298, S5: 0.203]

### Tables

##### Clinical codes for diagnosis and medications used in the analysis

**Table 1. ICD codes of MCI and AD for cohort selection. MCI: mild cognitive impairment; AD: Alzheimer's disease.**

| | ICD-9 | ICD-10 |
|---|---|---|
| **MCI** | 331.83, 294.9 | G31.84, F09 |
| **AD** | 331.0 | G30, G30.0, G30.1, G30.8, G30.9 |

**Table 2. Phecode of key comorbidities.**

| Comorbidity | Phecode | Comorbidity | Phecode |
|---|---|---|---|
| Hypertension | 401.1 | Parkinson’s disease | 332 |
| Hyperlipidemia | 272.1, 272.13 | Obesity | 278.1 |
| Diabetes | 250.1, 250.2 | Schizophrenia | 295.1 |
| Dementias | 278, 278.1 | Vitamin-D deficiency | 261.4 |
| Memory loss | 292.3 | Pneumonia | 480 |
| Heart disease | 401.21, 411.8 | Malignant neoplasm | 195.1 |
| Sleep disorders | 327 | Vitamin-B12 deficiency | 281.12 |
| Anxiety | 300.1, 300.11 | Bipolar disorder | 296.1 |
| GERD | 530.11 | Decubitus | 707.1 |
| Cerebrovascular disease | 433.8, 433 | Atherosclerosis | 440 |
| Chronic airway obstruction | 496 | Fatigue | 798 |
| Chronic renal failure (CKD) | 585.3 | Hypothyroidism | 244.4 |
| Urinary tract infection | 591 | Tobacco use disorder | 318 |
| Glaucoma and Cataract | 365, 366 | Seizures | 345.3 |
| Anemias | 285 | | |

**Table 3. ATC code (third level) of key medications used in OneFlorida and Mount Sinai dataset**

| Medication | ATC code | Medication | ATC code |
|---|---|---|---|
| Antithrombotic agents | B01A | Anti-dementia drugs | N06D |
| Lipid modifying agents | C10A | Drug for obstructive airway disease | R03B |
| Drugs for GORD | A02B | Antiepileptics | N03A |
| Opioids | N02A | Hypnotics and sedatives | N05C |
| Antidepressants | N06A | Drug for joint and muscular pain | M02A |
| Antiinfectives | S01A | Quinolone antibacterials | G04B |
| Corticosteroids | D07A | Urologicals | A10B |
| Beta blocking agents | C07A | Insulins and analogues | J01M |
| Anxiolytics | N05B | | |

**Table 4. TCGPI code of key medications used in MarketScan**

| Medication | TCGPI code | Medication | TCGPI code |
|---|---|---|---|
| Antithrombotic agents | 83x | Drug for obstructive airway disease | 45x |
| Drugs for GORD | 46x, 52x | Hypnotics and sedatives | 60x |
| Opioids | 65x | Quinolone antibacterials | 09x |
| Antidepressants | 58x | Urologicals | 56x |
| Antiinfectives | 01x, 16x | Insulins and analogues | 2710x |
| Beta blocking agents | 33x | | |

**Pairwise P-value of demographics between any two subphenotypes on the development cohort**

**Table 5. Pairwise P-value (FDR correction) between two subphenotypes on *gender* (development cohort)**

| | S1 | S2 | S3 | S4 | S5 |
|---|---|---|---|---|---|
| S1 | | 0.004 | 0.811 | 0.083 | <0.001 |
| S2 | | | 0.001 | <0.001 | 0.003 |
| S3 | | | | 0.114 | <0.001 |
| S4 | | | | | <0.001 |

**Table 6. Pairwise P-value (FDR correction) between subphenotypes on *race* (development cohort)**

| | S1 | S2 | S3 | S4 | S5 |
|---|---|---|---|---|---|
| S1 | | <0.001 | <0.001 | 0.052 | <0.001 |
| S2 | | | <0.001 | <0.001 | 0.027 |
| S3 | | | | 0.001 | 0.058 |
| S4 | | | | | 0.001 |

**Table 7. Pairwise P-value (FDR correction) between subphenotypes on *age of MCI onset* (development cohort)**

| | S1 | S2 | S3 | S4 | S5 |
|---|---|---|---|---|---|
| S1 | | 0.045 | 0.514 | <0.001 | <0.001 |
| S2 | | | 0.004 | <0.001 | <0.001 |
| S3 | | | | <0.001 | <0.001 |
| S4 | | | | | <0.001 |

**Table 8. Pairwise P-value (FDR correction) between subphenotypes on *progressive time* (development cohort)**

| | S1 | S2 | S3 | S4 | S5 |
|---|---|---|---|---|---|
| S1 | | <0.001 | <0.001 | 0.013 | <0.001 |
| S2 | | | 0.062 | <0.001 | 0.078 |
| S3 | | | | 0.047 | 0.001 |
| S4 | | | | | <0.001 |

#### Statistics on the MarketScan dataset (validation cohort)

**Table 9 Characteristics (making statistics on MCI onset) of the identified subphenotypes (validation cohort from MarketScan dataset). The p-value for Sex, Region, key comorbidity, and key medicine are obtained by $\chi^2$ test (false discovery rate correction for post-hoc pairwise comparisons in Sex and Region are in Table 10~11 in Supplement). The p-value for Age and Progression time are obtained by Kruskal-Wallis test (with Dunn's test for post-hoc pairwise comparisons in Table 12~13 in Supplement).**

| Variable | | Total | Subphenotype I | Subphenotype II | Subphenotype III | Subphenotype IV | Subphenotype V | P-value |
|---|---|---|---|---|---|---|---|---|
| No. of Patient (%) | | 18805 (100) | 2308 (12.27) | 2627 (13.97) | 3499 (18.61) | 1839 (9.78) | 2882 (15.33) | |
| **Age (MCI onset),** yr, Median (IQR) | | 79.0 [73.0~84.0] | 79.0 [72.0~84.0] | 78.0 [73.0~83.0] | 81.0 [77.0~85.0] | 83.0 [78.0~87.0] | 74.0 [66.0~80.0] | <0.001 |
| **Sex** female, N (%) | | 10897 (57.95) | 1339 (58.02) | 1303 (49.60) | 2169 (61.99) | 1394 (75.80) | 1895 (65.76) | <0.001 |
| **Region**, N (%) | Northeast | 3910 (20.79) | 455 (19.71) | 526 (20.02) | 762 (21.78) | 365 (19.85) | 588 (20.40) | <0.001 |
| | North central | 5753 (30.59) | 743 (32.19) | 898 (34.18) | 1022 (29.21) | 598 (32.52) | 815 (28.28) | |
| | West | 3253 (17.30) | 372 (16.12) | 416 (15.84) | 673 (19.23) | 358 (19.47) | 552 (19.15) | |
| | South | 5583 (29.69) | 690 (29.90) | 742 (28.25) | 977 (27.92) | 474 (25.77) | 890 (30.88) | |
| | Unknown | 306 (1.63) | 48 (2.08) | 45 (1.71) | 65 (1.86) | 44 (2.39) | 37 (1.28) | |
| **Progression time**, day, Median (IQR) | | 770.0 [537.5~1152.0] | 689.5 [491.0, 1059.25] | 839.0 [568.0~1231.0] | 815.0 [567.0~1214.5] | 745.0 [530.0~1108.5] | 897.5 [688.0~1274.75] | <0.001 |
| 1~2 year, N (%) | | 8694 (46.23) | 1240 (53.73) | 1043 (39.70) | 1476 (42.18) | 896 (48.72) | 875 (30.36) | |
| 2~3 year, N (%) | | 4854 (25.81) | 545 (23.61) | 738 (28.09) | 915 (26.15) | 470 (25.56) | 1027 (35.63) | |
| 3~4 year, N (%) | | 2701 (14.36) | 291 (12.61) | 427 (16.25) | 542 (15.49) | 263 (14.30) | 493 (17.11) | |
| 4~5 year, N (%) | | 1382 (7.35) | 122 (5.29) | 225 (8.56) | 316 (9.03) | 110 (5.98) | 259 (8.99) | |
| >5 year, N (%) | | 1174 (6.24) | 110 (4.77) | 194 (7.38) | 250 (7.14) | 100 (5.44) | 228 (7.91) | |
| **Key comorbidity,** N (%) | | | | | | | | |
| Hypertension | | 6775 (36.03) | 241 (10.44) | 359 (13.67) | 1970 (56.30) | 346 (18.81) | 173 (6.00) | <0.001 |
| Hyperlipidemia | | 3512 (18.68) | 246 (10.66) | 295 (11.23) | 1325 (37.87) | 158 (8.59) | 381 (13.22) | <0.001 |
| Diabetes | | 2890 (15.37) | 194 (8.41) | 960 (36.54) | 536 (15.32) | 226 (12.29) | 766 (26.58) | <0.001 |

| | | | | | | | |
|---|---|---|---|---|---|---|---|
| Dementias | 3691 (19.63) | 913 (39.56) | 297 (11.31) | 186 (5.32) | 365 (19.85) | 310 (10.76) | <0.001 |
| Memory loss | 3050 (16.22) | 894 (38.73) | 169 (6.43) | 241 (6.89) | 269 (14.63) | 212 (7.36) | <0.001 |
| Heart disease | 2109 (11.22) | 241 (10.44) | 329 (12.52) | 624 (17.83) | 63 (3.43) | 138 (4.79) | <0.001 |
| Sleep disorder | 1551 (8.25) | 123 (5.33) | 110 (4.19) | 163 (4.66) | 182 (9.90) | 326 (11.31) | <0.001 |
| Anxiety | 1203 (6.40) | 59 (2.56) | 193 (7.35) | 167 (4.77) | 73 (3.97) | 627 (21.76) | <0.001 |
| Gastroesophageal reflux disease | 1637 (8.71) | 123 (5.33) | 272 (10.35) | 747 (21.35) | 155 (8.43) | 143 (4.96) | <0.001 |
| Cerebrovascular disease | 1711 (9.10) | 389 (16.85) | 198 (7.54) | 201 (5.74) | 187 (10.17) | 130 (4.51) | <0.001 |
| Chronic airway obstruction | 1053 (5.60) | 64 (2.77) | 114 (4.34) | 280 (8.00) | 195 (10.60) | 212 (7.36) | <0.001 |
| Chronic renal failure (CKD) | 1654 (8.80) | 98 (4.25) | 682 (25.96) | 188 (5.37) | 72 (3.92) | 152 (5.27) | <0.001 |
| Urinary tract infection | 1985 (10.56) | 172 (7.45) | 206 (7.84) | 361 (10.32) | 159 (8.65) | 403 (13.98) | <0.001 |
| Glaucoma and Cataract | 884 (4.70) | 64 (2.77) | 233 (8.87) | 82 (2.34) | 32 (1.74) | 421 (14.61) | <0.001 |
| **Key Medicine,** N (%) | | | | | | | |
| Antithrombotic agents | 5829 (31.00) | 456 (19.76) | 736 (28.02) | 1293 (36.95) | 199 (10.82) | 326 (11.31) | <0.001 |
| Gastrointestinal agents | 5455 (29.01) | 422 (18.28) | 695 (26.46) | 1188 (33.95) | 317 (17.24) | 308 (10.69) | <0.001 |
| Opioids | 5089 (27.06) | 221 (9.58) | 458 (17.43) | 893 (25.52) | 729 (39.64) | 333 (11.55) | <0.001 |
| Antidepressants | 2768 (14.72) | 243 (10.53) | 727 (27.67) | 370 (10.57) | 161 (8.75) | 940 (32.62) | <0.001 |
| Antiinfectives | 5064 (26.93) | 543 (23.53) | 782 (29.77) | 348 (9.95) | 550 (29.91) | 355 (12.32) | <0.001 |
| Beta blocking agents | 3691 (19.63) | 450 (19.50) | 362 (13.78) | 1175 (33.58) | 290 (15.77) | 585 (20.30) | <0.001 |
| Hypnotics and sedatives | 1767 (9.40) | 195 (8.45) | 230 (8.76) | 222 (6.34) | 188 (10.22) | 364 (12.63) | <0.001 |
| Urological | 2049 (10.90) | 168 (7.28) | 173 (6.59) | 230 (6.57) | 169 (9.19) | 286 (9.92) | <0.001 |
| Insulins and analogues | 2591 (13.78) | 155 (6.72) | 1207 (45.95) | 223 (6.37) | 129 (7.01) | 509 (17.66) | <0.001 |

**Table 10. Pairwise P-value (FDR correction) between two subphenotypes on *gender* (validation cohort of MarketScan)**

| | S1 | S2 | S3 | S4 | S5 |
|---|---|---|---|---|---|
| **S1** | | <0.001 | 0.002 | <0.001 | <0.001 |
| **S2** | | | <0.001 | <0.001 | <0.001 |
| **S3** | | | | <0.001 | 0.002 |
| **S4** | | | | | <0.001 |

**Table 11. Pairwise P-value (FDR correction) between subphenotypes on *region* (validation cohort of MarketScan)**

| | S1 | S2 | S3 | S4 | S5 |
|---|---|---|---|---|---|
| **S1** | | 0.460 | 0.004 | 0.014 | 0.002 |
| **S2** | | | <0.001 | 0.011 | <0.001 |
| **S3** | | | | 0.045 | 0.047 |
| **S4** | | | | | <0.001 |

**Table 12. Pairwise P-value (FDR correction) between subphenotypes on *age of MCI onset* (validation cohort of MarketScan)**

| | S1 | S2 | S3 | S4 | S5 |
|---|---|---|---|---|---|
| **S1** | | <0.001 | <0.001 | <0.001 | <0.001 |
| **S2** | | | <0.001 | <0.001 | <0.001 |
| **S3** | | | | <0.001 | <0.001 |
| **S4** | | | | | <0.001 |

**Table 13. Pairwise P-value (FDR correction) between subphenotypes on *progressive time***
**(validation cohort of MarketScan)**

| | S1 | S2 | S3 | S4 | S5 |
|---|---|---|---|---|---|
| **S1** | | <0.001 | <0.001 | <0.001 | <0.001 |
| **S2** | | | 0.207 | <0.001 | <0.001 |
| **S3** | | | | <0.001 | <0.001 |
| **S4** | | | | | <0.001 |

#### Statistics of subphenotypes on the Mount Sinai dataset (validation cohort)

#### Table 14. Characteristics of the identified subphenotypes (validation cohort of Mount Sinai)

| Variable | | Total | Subphenotype I | Subphenotype II | Subphenotype III | P-value |
|---|---|---|---|---|---|---|
| No. of Patient (%) | | 689 | 125 (18.14) | 167 (24.24) | 197 (28.59) | -- |
| **Age (MCI onset),** yr, Median (IQR) | | 79.5 [74.7-85.3] | 80.0 [72.0~87.0] | 80.9 [76.9~85.9] | 83.0 [78.0~87.0] | 0.018 |
| **Sex** female, N (%) | | 455 (66.04) | 85 (68.00) | 93 (55.69) | 137 (69.54) | 0.014 |
| **Race,** N (%) | Caucasian | 426 (61.83) | 88 (70.4) | 89 (53.29) | 117 (59.39) | <0.001 |
| | African American | 139 (20.17) | 18 (14.40) | 46 (27.54) | 18 (19.29) | |
| | Asian | 7 (1.02) | 0 (0) | 2 (1.20) | 1 (0.51) | |
| | Other/Unknown | 117 (16.98) | 19 (15.20) | 30 (17.96) | 61 (30.96) | |
| **Progression time**, day, Median (IQR) | | 630 [402.5-936.0] | 540.0 [465.0~633.0] | 655.0 [569.0~803.0] | 605.0 [526.0~728.0] | <0.001 |
| 1~2 year, N (%) | | 344 (49.93) | 75 (60.00) | 78 (46.71) | 103 (52.88) | |
| 2~3 year, N (%) | | 198 (28.74) | 41 (32.80) | 49 (29.34) | 57 (28.93) | |
| 3~4 year, N (%) | | 71 (10.30) | 6 (4.80) | 17 (10.18) | 23 (11.68) | |
| 4~5 year, N (%) | | 46 (6.68) | 2 (1.60) | 9 (5.39) | 12 (6.09) | |
| >5 year, N (%) | | 30 (4.30) | 1 (0.80) | 14 (8.38) | 2 (1.02) | |
| **Key comorbidity,** N (%) | | | | | | |
| Hypertension | | 234 (33.96) | 10 (8.00) | 29 (17.37) | 79 (40.10) | <0.001 |
| Hyperlipidemia | | 107 (15.53) | 5 (4.00) | 28 (16.77) | 59 (29.95) | <0.001 |
| Diabetes | | 167 (24.24) | 6 (4.80) | 34 (20.36) | 70 (35.53) | <0.001 |
| Dementias | | 159 (23.08) | 47 (37.60) | 17 (10.18) | 26 (13.20) | <0.001 |

| | | | | | |
|---|---|---|---|---|---|
| Memory loss | 67 (9.72) | 22 (17.60) | 17 (10.18) | 14 (7.11) | 0.012 |
| Heart disease | 112 (16.26) | 21 (16.80) | 14 (8.38) | 38 (19.29) | 0.011 |
| Sleep disorder | 38 (5.52) | 5 (4.00) | 13 (7.78) | 15 (7.61) | 0.364 |
| Anxiety | 53 (7.69) | 10 (8.00) | 17 (10.18) | 12 (6.09) | 0.357 |
| Gastroesophageal reflux disease | 70 (10.16) | 14 (11.20) | 18 (10.78) | 30 (15.23) | 0.377 |
| Cerebrovascular disease | 94 (13.64) | 22 (17.60) | 6 (3.59) | 22 (11.17) | <0.001 |
| Chronic airway obstruction | 85 (12.34) | 6 (4.80) | 14 (8.38) | 25 (12.69) | 0.052 |
| Chronic renal failure (CKD) | 71 (10.30) | 4 (3.20) | 34 (20.36) | 15 (7.61) | <0.001 |
| Urinary tract infection | 125 (18.14) | 10 (8.00) | 24 (14.37) | 15 (7.61) | 0.069 |
| Glaucoma and Cataract | 39 (5.66) | 1 (0.80) | 18 (10.78) | 9 (4.57) | <0.001 |
| **Key Medicine**, N (%) | | | | | |
| Antithrombotic agents | 144 (20.90) | 12 (9.60) | 41 (24.55) | 43 (21.83) | 0.003 |
| Gastrointestinal agents | 153 (22.21) | 14 (11.20) | 47 (28.14) | 39 (19.80) | 0.001 |
| Opioids | 78 (11.32) | 16 (12.80) | 28 (16.77) | 20 (10.15) | 0.174 |
| Antidepressants | 74 (10.74) | 10 (8.00) | 27 (16.17) | 16 (8.12) | 0.024 |
| Antiinfectives | 101 (14.76) | 16 (12.80) | 26 (15.57) | 28 (14.21) | 0.798 |
| Corticosteroids | 84 (12.19) | 18 (14.40) | 23 (13.77) | 22 (11.17) | 0.640 |
| Beta blocking agents | 120 (17.42) | 24 (19.20) | 22 (13.17) | 49 (24.87) | 0.019 |
| Anti-dementia drugs | 106 (15.38) | 39 (31.20) | 25 (14.97) | 21 (10.66) | <0.001 |
| Hypnotics and sedatives | 48 (6.97) | 10 (8.00) | 9 (5.39) | 21 (10.66) | 0.187 |
| Urological | 65 (9.43) | 9 (7.20) | 22 (13.17) | 19 (9.64) | 0.234 |
| Insulins and analogues | 84 (12.19) | 4 (3.20) | 56 (33.53) | 16 (8.12) | <0.001 |

**Table 15. Mathematical notations in DMTM.**

| Notations | Meaning |
| --- | --- |
| $\boldsymbol{x}_{n,t}^{(m)} \in \{0,1\}^{V_m}$ | The original binary feature vector ($m$-th modality) of $n$-th patient at $t$-th visit, where 1 means having this event, and 0 means having no this event |
| $\boldsymbol{m = 1, \cdots, M}$ | Index of modality, where $M$ is the total number of modalities |
| $\boldsymbol{V_m}$ | Total unique number of clinical events of $m$-th modality |
| $\boldsymbol{T_n}$ | Total number of visits for the $n$-th patient |
| $\boldsymbol{k = 1, \cdots, K}$ | Index of clinical topics, where $K$ is the total number of topics |
| $\boldsymbol{\Phi}^{(m)} \in \mathcal{R}_{+}^{V_m \times K}$ | The matrix of clinical topics of $m$-th modality, where each column correspondents to one topic |
| $\phi_{\mathrm{k}}^{(\mathrm{m})}$ | The $k$-th column of $\boldsymbol{\Phi}^{(m)}$, representing $k$-th topic of $m$-th modality. It is a distribution over all unique events in $m$-th modality |
| $\boldsymbol{\theta}_{n,t} \in \mathcal{R}_{+}^{K}$ | Topic weight vector of $n$-th patient at $t$-th visit |
| $\boldsymbol{\Pi} \in \mathcal{R}_{+}^{K \times K}$ | The transition matrix of clinical topics, where each element, $\pi_{ij}$, represents the probability of transition from $i$-th topic to the $j$-th topic |
| $\{\boldsymbol{v_k}\}_{\boldsymbol{k=1}}^{\boldsymbol{K}}$ | Parameters in the prior of $\boldsymbol{\Pi}$ |
| $\boldsymbol{u}_{n,t}^{(m)}$ | The latent count variables sampled by Bernoulli-Poisson link with the original binary features $\boldsymbol{x}_{n,t}^{(m)}$ |
| $\tau_0, \gamma_0, \eta_0, \epsilon_0$ | Hyperparameters |

**Table 16. To figure out the reason why five key topics are missed on the Mount Sinai dataset, we compare the percentage of occurrence of top diagnosis records in these topics on three cohorts (see details in Supplemental Method). If the percentage of occurrence of one diagnosis event is the most in one cohort, we highlight it by boldface. We found that these diagnosis events occur less frequently on the Mount Sinai cohort.**

| Clinical topic | PheCode (Name) | Percentage on the OneFlorida cohort (%) | Percentage on the MarketScan cohort (%) | Percentage on the Mount Sinai cohort (%) |
|---|---|---|---|---|
| Bone | 741.5 (Hemarthrosis) | 19.03 | 6.72 | 1.64 |
| | 760 (Back pain) | 20.92 | 23.17 | 8.46 |
| | 740.9 (Osteoarthrosis) | 15.81 | 6.32 | 3.29 |
| | 721 (Spondylosis and allied disorders) | 10.17 | 2.77 | 1.76 |
| | 729.1 (Rheumatism, unspecified and fibrositis) | 14.56 | 5.98 | 0.96 |
| | 722.6 (Degeneration of intervertebral disc) | 1.68 | 19.24 | 1.57 |
| | 721.1 (Spondylosis without myelopathy) | 2.55 | 15.39 | 4.68 |
| | 720 (Spinal stenosis of lumbar region) | 4.21 | 14.20 | 6.60 |
| | 761 (Cervicalgia) | 4.39 | 16.85 | 2.94 |
| Movement | 728 (Disorders of muscle, ligament, and fascia) | 22.58 | 3.26 | 2.45 |
| | 741.3 (Difficulty in walking) | 19.65 | 16.32 | 9.76 |
| | 350.2 (Abnormality of gait) | 15.76 | 24.79 | 3.29 |
| | 350.3 (Lack of coordination) | 10.44 | 7.32 | 1.01 |
| | 350.1 (Abnormal involuntary movements) | 9.02 | 4.21 | 2.29 |
| | 740.9 (Osteoarthrosis) | 15.81 | 6.32 | 3.29 |
| | 772.3 (Muscle weakness) | 4.38 | 11.67 | 3.71 |
| | 745 (Pain in joint) | 9.60 | 13.97 | 7.28 |
| Mental | 300.11(Generalized anxiety disorder) | 17.26 | 15.32 | 10.43 |
| | 296.22 (Major depressive disorder) | 20.03 | 18.46 | 6.14 |
| | 389 (Hearing loss) | 5.41 | 11.02 | 3.17 |
| | 296 (Mood disorders) | 4.36 | 2.10 | 0.98 |
| | 389.3 (Degenerative and vascular disorders of ear) | 3.62 | 0.96 | 1.34 |
| | 300.4 (Dysthymic disorder) | 1.67 | 3.14 | 0.76 |
| | 296.1 (Bipolar) | 10.04 | 12.31 | 5.62 |
| Digest system | 785 (Abdominal pain) | 20.40 | 19.36 | 12.31 |
| | 530.11 (GERD) | 21.36 | 22.10 | 10.02 |
| | 789 (Nausea and vomiting) | 8.36 | 4.21 | 2.10 |
| | 564 (Functional digestive disorders) | 8.15 | 3.03 | 0.67 |
| | 535 (Gastritis and duodenitis) | 3.15 | 0.17 | 1.34 |
| | 562.1 (Diverticulosis) | 4.52 | 13.66 | 2.01 |
| | 561.1 (Diarrhea) | 1.92 | 10.93 | 0.63 |
| Respiratory system | 496 (Chronic airway obstruction) | 19.57 | 15.22 | 9.97 |
| | 496.21 (Obstructive chronic bronchitis) | 7.72 | 8.54 | 4.20 |
| | 480 (Pneumonia) | 12.34 | 10.03 | 7.01 |
| | 513 (Respiratory abnormalities) | 14.23 | 4.10 | 2.20 |
| | 514 (Abnormal findings examination of lungs) | 6.51 | 6.42 | 1.07 |
| | 512.8 (Cough) | 10.27 | 16.39 | 8.75 |
| | 512.7 (Shortness of breath) | 4.74 | 7.72 | 0.97 |
